# Multimodal MRI Marker of Cognition Explains the Association Between Cognition and Mental Health in UK Biobank

**DOI:** 10.1101/2025.03.18.25324202

**Authors:** Irina Buianova, Mateus Silvestrin, Jeremiah Deng, Narun Pat

## Abstract

**Background:** Cognitive dysfunction often co-occurs with psychopathology. Advances in neuroimaging and machine learning have led to neural indicators that predict individual differences in cognition with reasonable performance. We examined whether these neural indicators explain the relationship between cognition and mental health in the UK Biobank cohort (*n* > 14000).

**Methods:** Using machine learning, we quantified the covariation between general cognition and 133 mental health indices and derived neural indicators of cognition from 72 neuroimaging phenotypes across diffusion-weighted MRI (dwMRI), resting-state functional MRI (rsMRI), and structural MRI (sMRI). With commonality analyses, we investigated how much of the cognition–mental health covariation is captured by each neural indicator and neural indicators combined within and across MRI modalities.

**Results:** The predictive association between mental health and cognition was at out-of-sample *r* = 0.3. Neuroimaging phenotypes captured 2.1% to 25.8% of the cognition–mental health covariation. The highest proportion of variance explained by dwMRI was attributed to the number of streamlines connecting cortical regions (19.3%), by rsMRI through functional connectivity between 55 large-scale networks (25.8%), and by sMRI via the volumetric characteristics of subcortical structures (21.8%). Combining neuroimaging phenotypes within modalities improved the explanation to 25.5% for dwMRI, 29.8% for rsMRI, and 31.6% for sMRI, and combining them across all MRI modalities enhanced the explanation to 48%.

**Conclusions:** We present an integrated approach to derive multimodal MRI markers of cognition that can be transdiagnostically linked to psychopathology. This demonstrates that the predictive ability of neural indicators extends beyond the prediction of cognition itself, enabling us to capture the cognition–mental health covariation.

## Introduction

Cognition and mental health are closely intertwined [1]. Cognitive dysfunction is present in various mental illnesses, including anxiety [2, 3], depression [4–6], and psychotic disorders [7–12]. National Institute of Mental Health’s Research Domain Criteria (RDoC) [13,14] treats cognition as one of the main basic functional domains that transdiagnostically underly mental health. According to RDoC, mental health should be studied in relation to cognition, alongside other domains such as negative and positive valence systems, arousal and regulatory systems, social processes, and sensorimotor functions. RDoC further emphasizes that each domain, including cognition, should be investigated not only at the behavioural level but also through its neurobiological correlates. In this study, we aim to examine how the covariation between cognition and mental health is reflected in neural markers of cognition, as measured through multimodal neuroimaging.

Recent efforts in brain Magnetic Resonance Imaging (MRI) and machine learning have led to predictive models that allow us to create MRI-based neural indicators of cognition with reasonable predictive performance [15–17]. These models are designed to predict cognition based on different cognitive tasks in unseen individuals who are not part of the modeling process [18, 19]. Yet, the extent to which MRI-based neural indicators designed to predict cognition capture the same variance that mental health shares with cognition remains unknown. Demonstrating that MRI-based neural indicators of cognition capture the covariation between cognition and mental health will thereby support the utility of such indicators for understanding the etiology of mental health [20].

Different MRI modalities measure different aspects of the brain, and MRI quantification techniques capture different brain features, resulting in distinct neuroimaging phenotypes. This means there are numerous approaches to derive neural indicators of cognition from MRI data. For example, diffusion-weighted MRI (dwMRI) measures the shape and amount of water diffusion in various directions and tissue compartments [21]. Different dwMRI metrics, such as fractional anisotropy (FA), which quantifies the degree of water diffusion directionality, and the streamline count, which indirectly reflects structural connectedness between the two regions (structural connectome), provide information about white matter orientation, density, and microstructural integrity [22–24]. Resting-state functional MRI (rsMRI) measures spontaneous low-frequency fluctuations in the Blood Oxygenation Level Dependent (BOLD) signal in the absence of a task, enabling the investigation of resting-state functional connectivity (RSFC) [25]. RSFC from rsMRI can be estimated between pairs of parcellated grey matter regions (functional connectome) or between widespread networks derived from the Independent Component Analysis (ICA). Structural MRI (sMRI) uses T1-weighted and T2-weighted imaging to quantify various aspects of brain anatomy and morphology. For example, the morphology of the cerebral cortex and white matter can be quantified by measuring grey or white matter thickness, volume, and area in regions defined by different atlases, whereas the characteristics of subcortical regions are conventionally quantified with volumes of subcortical nuclei and their subdivisions [26, 27]. Previous studies using machine learning have shown that both a) the choice of MRI modality and b) the quantification method within each modality affect the performance of MRI-based models in capturing cognition [15, 17, 28]. Dhamala and colleagues found that the predictive ability of structural and functional connectomes largely depends on the choice of atlases used to parcellate grey matter and how they were derived [28].

Given the heterogeneity of neuroimaging phenotypes from different MRI modalities, drawing information across them may boost the predictive ability of MRI-based neural indicators [29]. One way to integrate multiple neuroimaging phenotypes across MRI modalities is a stacking approach, which employs two levels of machine learning. First, researchers build a predictive model from each neuroimaging phenotype (e.g., cortical thickness from different grey matter parcellations) to predict a target variable (e.g., cognition). Next, in the stacking level, they use predicted values (i.e., cognition predicted from each neuroimaging phenotype) from the first level as features to predict the target variable [15]. Previous studies show that integrating multimodal neuroimaging phenotypes into “stacked models” enhances the prediction of cognition [15–17, 30]. Here, we aim to determine whether this improvement extends beyond the prediction of cognition itself, allowing us to capture more covariation between cognition and mental health.

Using the largest population-level neuroimaging dataset, the UK Biobank, we investigated (a) which neuroimaging phenotypes yield a neural indicator of cognition that explains the relationship between cognition and mental health the most, and (b) whether combining neuroimaging phenotypes within and across MRI modalities enhances the explanation of this relationship. We started by deriving a general cognition factor, or the *g*-factor, from twelve cognitive scores from different tasks. The *g*-factor underlies variability across cognitive domains and reflects the overall cognition [31, 32]. Next, we applied machine learning to predict the *g*-factor from 133 mental health indices and 72 neuroimaging phenotypes in unseen participants. For neuroimaging, we created predictive models from both individual neuroimaging phenotypes and phenotypes combined within and across three MRI modalities via stacking. Finally, we conducted commonality analyses [33] to quantify the contribution of neural indicators of cognition based on different neuroimaging phenotypes to explaining the relationship between cognition and mental health.

## Methods and Materials

### Study Design and Participants

We utilized cognition, mental health, and MRI data collected at the first imaging visit (2014-2019) and additional mental health data collected online from the UK Biobank (UK Biobank Resource Application 70132), a prospective epidemiological study involving individuals aged 40 to 69 years at recruitment. The analyses included participants who had all brain MRI scans, performed all cognitive tests, and completed mental health questionnaires (Table 1).

**Table 1.**
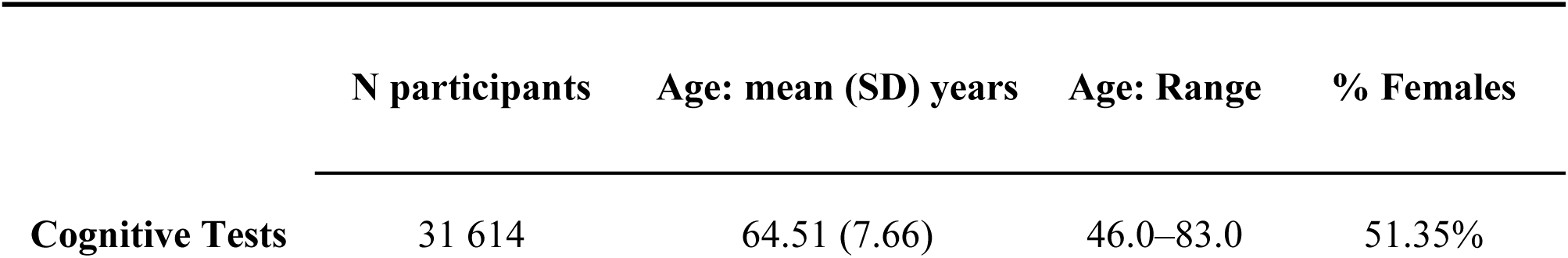
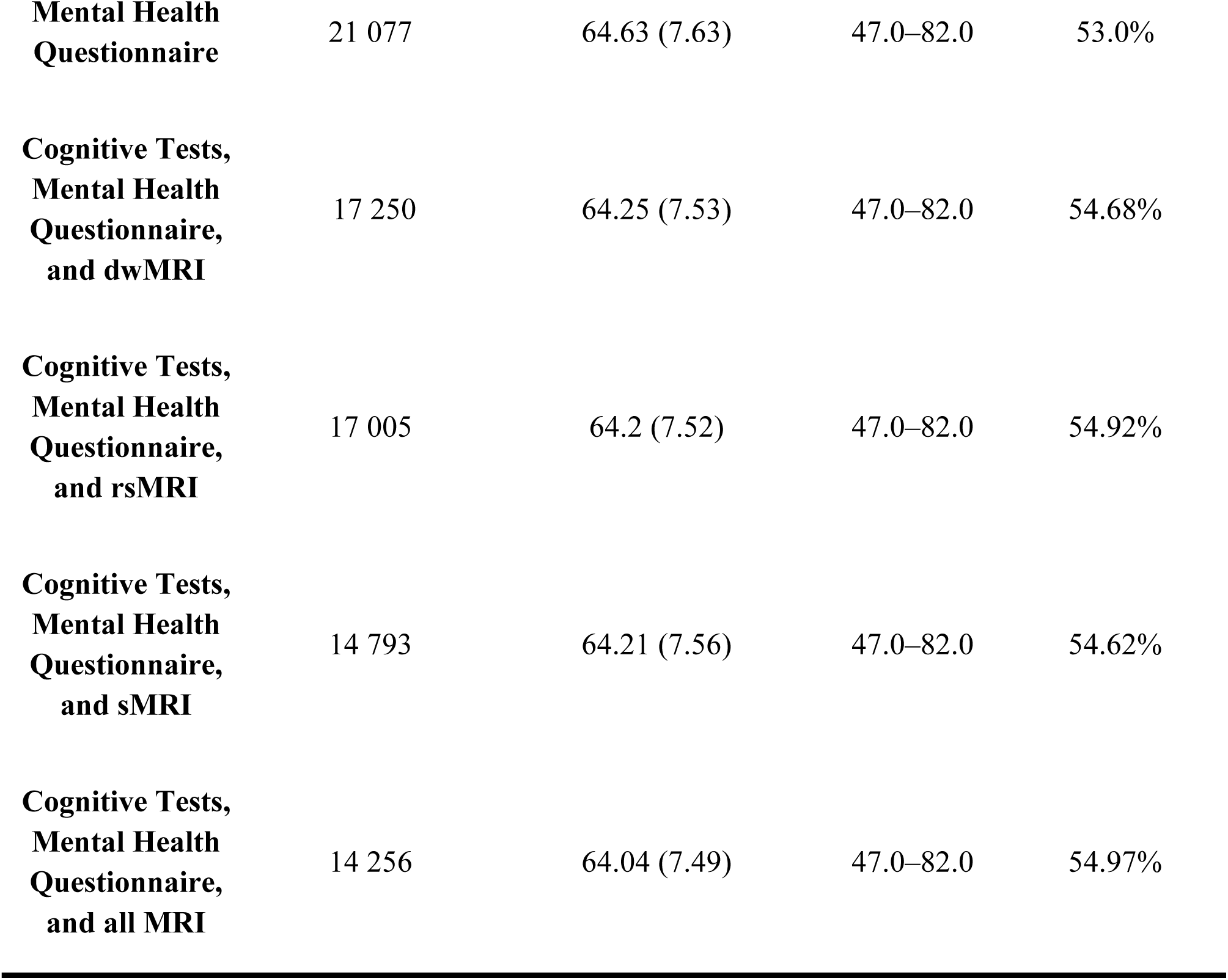
Demographics for each subsample analysed: number, age, and sex of participants who completed all cognitive tests, mental health questionnaires, and MRI scanning.

### Cognition

To derive the *g*-factor, we used twelve scores from eleven cognitive tests (Fig. 1 and Tables S1–S3) [34]. A detailed description of cognitive tasks is provided in the UK Biobank showcase (http://biobank.ndph.ox.ac.uk/showcase/label.cgi?id=100026) and discussed elsewhere [34]. To capture overall variability in cognition, we built a *g*-factor hierarchical model using exploratory structural equation modeling (ESEM). Here, the *g*-factor underlies a set of latent factors which, in turn, underlie 12 cognitive scores (Fig. 1, 2 and Supplementary Materials, S1).

**Figure 1.**
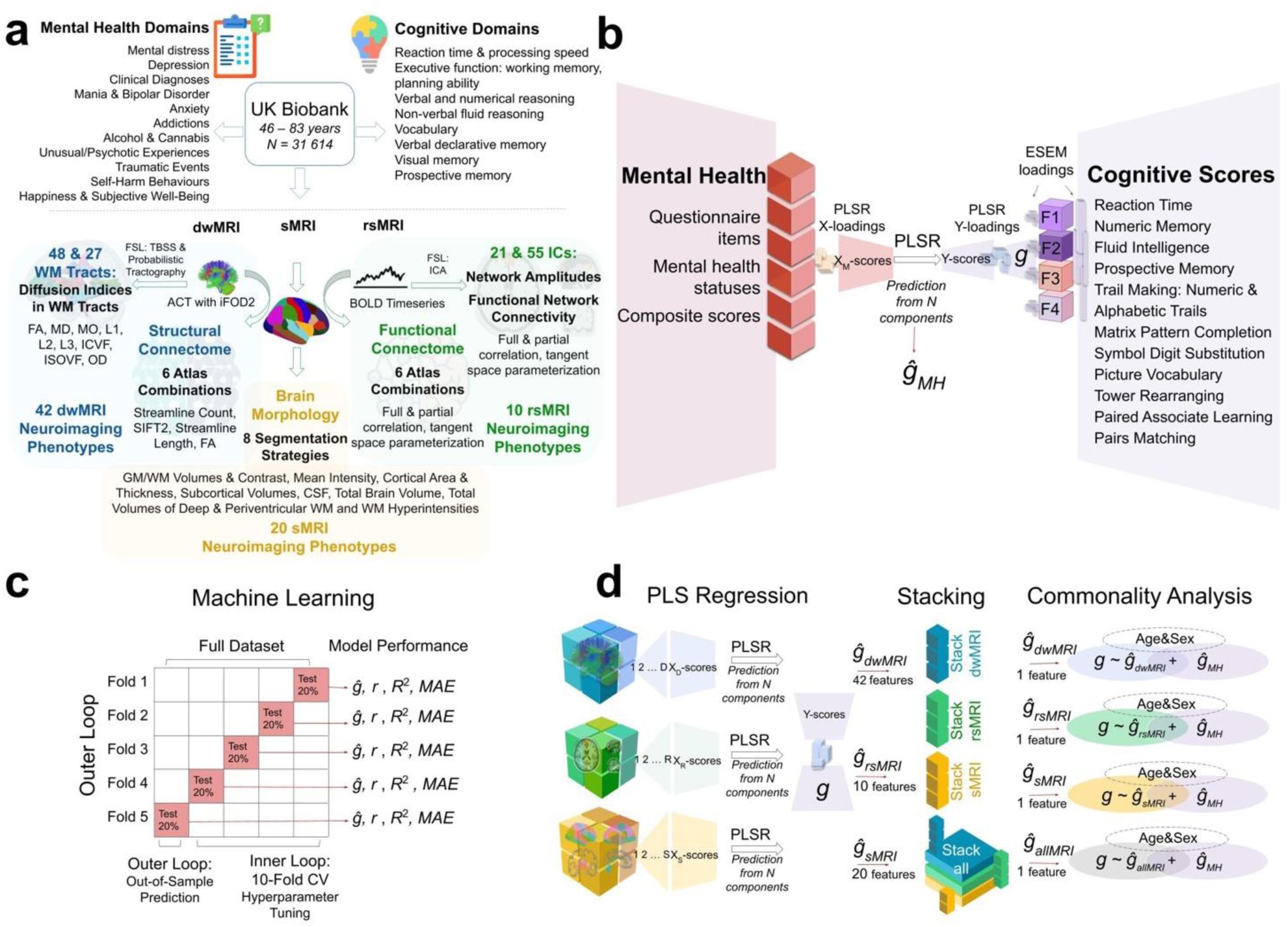
Experimental design. **a.** UK Biobank variables: cognitive tests, mental health, and neuroimaging phenotypes from three MRI modalities. **b** Derivation of the *g*-factor from cognitive performance scores with Exploratory Structural Equation Modeling (ESEM) and prediction of the *g*-factor from mental health indices using Partial Least Squares Regression (PLSR). **c** Scheme of the machine learning model (PLSR) with nested cross-validation. **d** Scheme of the two-level predictive modeling and commonality analyses. First, individual neuroimaging phenotypes from dwMRI (42 phenotypes), rsMRI (10 phenotypes), and sMRI (20 phenotypes) are used as features to predict the *g*-factor. Then, *g*-factor values predicted from distinct neuroimaging phenotypes are combined within each modality (“dwMRI Stacked”, “rsMRI Stacked”, and “Stacked sMRI”) as well as across all modalities (“All MRI Stacked”) and used as features, resulting in one predicted value per subject per stacked model (*ĝdwMRI*, *ĝrsMRI*, *ĝsMRI*, and *ĝallMRI*). Finally, values predicted from MRI data together with the values predicted from mental health indices (*ĝMH*) are used as independent explanatory variables in commonality analyses. *XD-scores*, *XR-scores*, *Xs-scores*, and *Y-scores*, weighted linear combinations of the original features (dwMRI, rsMRI, and sMRI neuroimaging phenotypes, respectively) in PLSR; *WM*, white matter; *TBSS*, tract-based spatial statistics; *ACT*, anatomically-constrained tractography; *iFOD2*, Fiber Orientation Distributions; *FA*, fractional anisotropy; *MD*, mean diffusivity; *MO*, diffusion tensor mode; *L1*, *L2*, *L3*, eigenvalues of the diffusion tensor; *ICVF*, intracellular volume fraction; *OD*, orientation dispersion index; *ISOVF*, isotropic volume fraction; *BOLD*, blood oxygenation level dependent signal; *ICA*, independent component analysis; *GM*, grey matter; *CSF*, cerebrospinal fluid; *F1*, *F2*, *F3*, and *F4*, latent factors from ESEM; *ESEM loadings*, loadings of the test scores onto the latent factors and loadings of the latent factors onto the *g*-factor; *X-loadings* and *Y-loadings*, loadings of the predictor (mental health measures; X) and target (*g*-factor; Y) variables; respectively, onto the PLSR components; *XM-scores* and *Y-scores*, the weighted linear combinations of the original predictor (mental health measures) and target (*g*-factor) variables, respectively; *ĝMH*, values of the *g*-factor predicted from mental health features; *ĝ*, predicted values of the *g*-factor; *r*, Pearson *r* (between original and predicted values of the *g*-factor); *R*^2^, coefficient of determination (between original and predicted values of the *g*-factor); *MAE*, mean absolute error; *CV*, cross-validation.

### Mental Health

Mental health measures encompassed 133 variables from twelve groups: mental distress, depression, clinical diagnoses related to the nervous system and mental health, mania (including bipolar disorder), neuroticism, anxiety, addictions, alcohol and cannabis use, unusual/psychotic experiences, traumatic events, self-harm behaviours, and happiness and subjective well-being (Fig. 1 and Tables S4 and S5). We included both self-report questionnaire items from all participants and composite diagnostic scores computed following Davis et al. and Dutt et al. [35,36] as features in our first-level (for explanation, see Data analysis section) Partial Least Squares Regression (PLSR) model. This approach leverages PLSR’s ability to handle multicollinearity through dimensionality reduction, enabling simultaneous use of granular symptom-level information and robust composite measures (for mental health scoring details, see Supplementary Materials, S2). We assess the contribution of each mental health index to general cognition by examining the direction and magnitude of its PLSR-derived loadings on the identified latent variables.

### Neuroimaging

For MRI-based models, we used dwMRI, rsMRI, and sMRI (Fig. 1 and Table S6). MRI acquisition protocols are available at http://biobank.ctsu.ox.ac.uk/crystal/refer.cgi?id=2367. MRI processing pipelines are described in the UK Biobank brain imaging documentation (https://biobank.ctsu.ox.ac.uk/crystal/crystal/docs/brain_mri.pdf) and discussed elsewhere (Supplementary Materials, S3) [37, 38].

MRI data processed by the UK Biobank are available as imaging-derived phenotypes (IDPs). In addition to the UK Biobank’s IDPs, we used structural connectomes and BOLD time series created by Mansour and colleagues [39]. We used structural connectomes for six combinations of cortical and subcortical atlases: the Desikan-Killiany (aparc) cortical atlas [40] + Melbourne Subcortical Atlas scale I (MSA-I) [41], the Destrieux (aparc.a2009s) cortical atlas [42] + MSA-I [41], the Glasser cortical atlas [43] + MSA-I [41], the Glasser cortical atlas [43] + MSA-IV [41], the Schaefer atlas for 200 cortical regions (7 networks) [44, 45] + MSA-I [42], and the Schaefer atlas for 500 cortical regions (7 networks) [44, 45] + MSA-IV [41]. Since the BOLD time series data were provided separately for cortical and subcortical parcellations, we combined them according to the combinations used for structural connectomes (Table S6).

### Diffusion-Weighted MRI (dwMRI)

To quantify dwMRI, we used 42 neuroimaging phenotypes. dwMRI IDPs included fractional anisotropy (FA), diffusion tensor mode (MO), mean diffusivity (MD), and three eigenvalues of the diffusion tensor (L1, L2, L3) derived from diffusion tensor fitting, as well as intracellular volume fraction (ICVF), isotropic or free water volume fraction (ISOVF), and the orientation dispersion index (OD) from NODDI (Neurite Orientation Dispersion and Density Imaging) for 48 and 27 white matter tracts reconstructed with tract-based spatial statistics (TBSS) and probabilistic tractography, respectively [46–51]. Structural connectome data included matrices with quantitative metrics for each node pair from six atlas combinations. The metrics included streamline count, fibre bundle capacity (from Spherical-Deconvolution Informed Filtering of Tractograms, SIFT2), mean streamline length, and mean FA (Table S6) [39].

### Resting-State Functional MRI (rsMRI)

To quantify rsMRI, we leveraged 10 neuroimaging phenotypes. rsMRI IDPs included functional connectivity metrics – normalized temporal and partial temporal correlations between nodes’ time series – and network amplitudes (the standard deviations of network time series [52]) for 21 and 55 networks (Independent Components, ICs) from ICA applied at 25 and 100 dimensions. We also applied tangent space parameterization to 21 and 55 networks to derive an additional metric for RSFC. Tangent space parameterization matrices were computed as deviations of subjects’ connectivities from the group connectivity matrix [53–55]. To obtain functional connectomes from the parcellated BOLD time series data, we calculated full and partial Pearson correlations for each node pair and applied tangent parameterization to six atlas combinations (Table S6).

### Structural MRI (sMRI)

To quantify sMRI, we used 20 neuroimaging phenotypes. sMRI IDPs from T1-weighted MRI included grey and white matter volumes and mean intensity, cortical area and thickness, the volume of subcortical structures and cerebrospinal fluid, grey/white matter contrast (fractional contrast between grey and white matter intensities), and total brain volume for eight segmentation strategies: regional grey matter volumes (FSL FAST) [56], subcortical volumes (FSL FIRST) [57], FreeSurfer automated subcortical volumetric segmentation (ASEG), FreeSurfer *ex-vivo* Brodmann Area Maps, FreeSurfer Destrieux (a2009s), FreeSurfer Desikan-Killiany-Tourville parcellation, FreeSurfer Desikan-Killiany parcellation (grey/white matter intensity, pial surface, and white matter surface), and FreeSurfer subcortical volumetric subsegmentation [58]. sMRI IDPs from T2-weighted MRI included the total volumes of deep white matter, periventricular white matter, and white matter hyperintensities (Table S6).

We corrected each MRI modality for common and specific sets of MRI-related confounds described in detail in Alfaro-Almagro et al. (Table S7) [59]. The procedure for confound modeling is discussed in Supplementary Materials (S4).

### Data analysis

We employed nested cross-validation to predict cognition from mental health indices and 72 neuroimaging phenotypes (Fig. 1). Nested cross-validation is a robust method for evaluating machine learning models while tuning their hyperparameters, ensuring that performance estimates are both accurate and unbiased. Here, we used a nested cross-validation scheme with five outer folds and ten inner folds.

We started by dividing the entire dataset into five outer folds. Each fold took a turn being held out as the outer-fold test set (20% of the data), while the remaining four folds (80% of the data) were used as an outer-fold training set. Within each outer-fold training set, we performed a second layer of cross-validation – this time splitting the data into ten inner folds. These inner folds were used exclusively for hyperparameter tuning: models were trained on nine of the inner folds and validated on the remaining one, cycling through all ten combinations.

We then selected the hyperparameter configuration that performed best across the inner-fold validation sets, as determined by the minimal mean squared error (*MSE*). The model was then retrained on the full outer-fold training set using this hyperparameter configuration and evaluated on the outer-fold test set, using four performance metrics: Pearson *r*, the coefficient of determination (*R*^2^), the mean absolute error (*MAE*), and the *MSE*. This entire process was repeated for each of the five outer folds, ensuring that every data point is used for both training and testing, but never at the same time. We opted for five outer folds instead of ten to reduce computational demands, particularly memory and processing time, given the substantial volume of neuroimaging data involved in model training. Five outer folds led to an outer-fold test set at least *n* = 4 000, which should be sufficient for model evaluation. In contrast, we retained ten inner folds to ensure robust and stable hyperparameter tuning, maximising the reliability of model selection.

To model the relationship between mental health and cognition, we employed Partial Least Squares Regression (PLSR) to predict the *g*-factor from 133 mental health variables. To model the relationship between neuroimaging data and cognition, we used a two-step stacking approach [15–17,61] to integrate information from 72 neuroimaging phenotypes across three MRI modalities. In the first step, we trained 72 base (first-level) PLSR models, each predicting the *g*-factor from a single neuroimaging phenotype. In the second step, we used the predicted values from these base models as input features for stacked models, which again predicted the *g*-factor. We constructed four stacked models based on the source of the base predictions: one each for dwMRI, rsMRI, sMRI, and a combined model incorporating all modalities (“dwMRI Stacked”, “rsMRI Stacked”, “sMRI Stacked”, and “All MRI Stacked”, respectively). Each stacked model was trained using one of four machine learning algorithms – ElasticNet, Random Forest, XGBoost, or Support Vector Regression – selected individually for each model (see Supplementary Materials, S6).

For rsMRI phenotypes, we treated the choice of functional connectivity quantification method – full correlation, partial correlation, or tangent space parametrization – as a hyperparameter. The method yielding the highest performance on the outer-fold training set was selected for predicting the *g*-factor (see Supplementary Materials, S5).

To prevent data leakage, we standardized the data using the mean and standard deviation derived from the training set and applied these parameters to the corresponding test set within each outer fold. This standardization was performed at three key stages: before *g*-factor derivation, before regressing out modality-specific confounds from the MRI data, and before stacking. Similarly, to maintain strict separation between training and testing data, both base and stacked models were trained exclusively on participants from the outer-fold training set and subsequently applied to the corresponding outer-fold test set.

To evaluate model performance and assess statistical significance, we aggregated the predicted and observed *g*-factor values from each outer-fold test set. We then computed a bootstrap distribution of Pearson’s correlation coefficient (*r*) by resampling with replacement 5 000 times, generating 95% confidence intervals (CIs) (Fig. 1). Model performance was considered statistically significant if the 95% CI did not include zero, indicating that the observed associations were unlikely to have occurred by chance.

Finally, we conducted a series of commonality analyses to quantify the proportion of the cognition–mental health relationship captured by MRI-based neural indicators [34]. We explain this analysis in detail in Supplementary Materials (S7). Briefly, for both observed (i.e., derived directly from cognitive scores, not mental health or MRI data) and predicted *g*-factors (i.e., predicted from either mental health indices or neuroimaging phenotypes using the top-performing algorithm), we separately pooled values across all five outer-fold test sets to reconstruct the complete dataset. We then applied a series of linear regression models, treating the observed *g*-factor as a response variable and the *g*-factor predicted from mental health (mental health-*g*) and neuroimaging phenotypes (neuroimaging-*g*) as two separate explanatory variables. We then decomposed the variance of the observed *g*-factor into the variance explained uniquely or commonly by mental health-*g* and neuroimaging-*g*. We quantified the contribution of each neuroimaging phenotype to explaining the cognition–mental health relationship by a percentage ratio between (a) the common effects between mental health-*g* and neuroimaging-*g* and (b) the total effects of mental health-*g*. That is, the nominator is the common variance that neuroimaging-*g* shares with mental health-*g* in explaining the observed *g*-factor, and the denominator is the variance of mental health-*g* that explains the observed *g*-factor regardless of neuroimaging-*g*.

To determine which neuroimaging features contribute most to the predictive performance of top-performing phenotypes within each modality, while accounting for the potential latent components derived from neuroimaging, we assessed feature importance using the Haufe transformation [62]. Specifically, we calculated Pearson correlations between the predicted *g*-factor and scaled and centred neuroimaging features across five outer-fold test sets. We also examined whether the performance of neuroimaging phenotypes in predicting cognition *per se is* related to their ability to explain the link between cognition and mental health. Here, we computed the correlation between the predictive performance of each neuroimaging phenotype and the proportion of the cognition–mental health relationship it captures. To understand how demographic factors, including age and sex, contribute to this relationship, we also conducted a separate set of commonality analyses treating age, sex, age^2^, age×sex, and age^2^×sex as an additional set of explanatory variables (Fig. 1).

## Results

### Derivation of the *g*-factor

In each fold, model performance estimates suggested good data factorability. According to parallel factor analysis, four factors were sufficient to explain the latent structure of the dataset (Fig. 2a). ESEM further supported the construct validity of factor structure (Fig. 2a). The *g*-factor captured around 39% of the variance in cognitive scores (Supplementary Materials, S8, Tables S8–S11).

**Figure 2.**
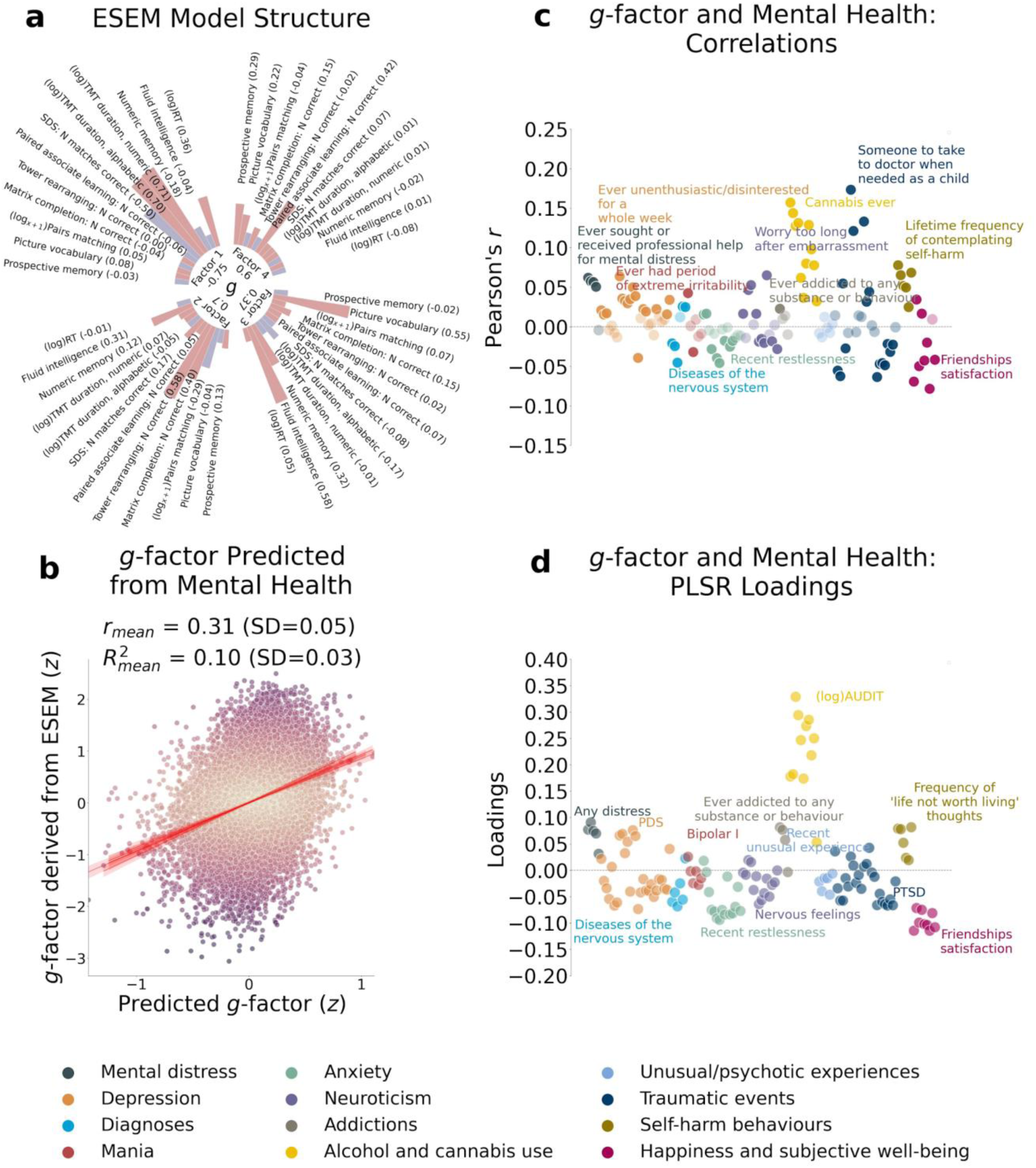
*g*-factor modeling and the relationship between the *g*-factor and mental health features. In our analysis, we derived the *g*-factor and built machine learning models in each outer fold separately. For visualization purposes, we display the results of the (a) Exploratory Structural Equation Modeling (ESEM) performed on a sample of 31 614 participants and (b) Partial Least Squares Regression (PLSR) model for the *g*-factor and mental health built on a sample of 21 077 participants with a single train/test split (80%/20%) as representations of ESEM and PLSR model structures across five folds. **a** Loadings of the cognitive test scores onto four latent factors and loadings of the latent factors onto the *g*-factor based on the ESEM. **b** Scatterplot of the observed *g*-factor and *g*-factor predicted from mental health indices with PLSR. Out-of-sample predictive performance of the PLSR model is evaluated with Pearson *r* and *R*^2^ averaged across five folds. **c** Pearson correlations between the *g*-factor and mental health indices. Mental health features are grouped into categories. Within each category, a feature with the highest absolute value of Pearson *r* is annotated. Pale and bright dots represent non-significant and significant correlations, respectively. **d** Loadings of mental health indices in the PLSR model showing the relationships between the features (mental health) and the target variable (*g*-factor). The loadings are averaged across all PLSR components and weighted by *R*^2^ in the training set. Mental health features are grouped into categories. Within each category, a feature with the highest absolute value of the loading is annotated. *SD*, standard deviation (mean across five folds).

### Predictive Modeling

#### Mental health

On average, information about mental health predicted the *g*-factor at *R*^2^_mean_ = 0.10 and *r*_mean_ = 0.31 (95% CI [0.291, 0.315]; Fig. 2b and 2c and Supplementary Materials, S9, Table S12).

The magnitude and direction of factor loadings for mental health in the PLSR model allowed us to quantify the contribution of individual mental health indices to cognition. Overall, the scores for mental distress, alcohol and cannabis use, and self-harm behaviours relate positively, and the scores for anxiety, neurological and mental health diagnoses, unusual or psychotic experiences, happiness and subjective well-being, and negative traumatic events relate negatively to cognition.

#### MRI

The predictive performance of neuroimaging phenotypes varied from low to moderate. At the modality level, rsMRI and dwMRI showed the highest and lowest performance, respectively (Fig. 3). On average, neuroimaging phenotypes stacked within dwMRI, rsMRI, and sMRI predicted the *g*-factor at *R*^2^_mean_ = 0.073, 0.105, and 0.095, and *r*_mean_ = 0.27 (95% CI [0.252, 0.273]), 0.33 (95% CI [0.308, 0.331]), and 0.3 (95% CI [0.284, 0.307]), respectively. Stacking all 72 neuroimaging phenotypes boosted the predictive performance of the MRI-based model for cognition, yielding *R*^2^_mean_ = 0.159 and *r*_mean_ = 0.398 (95% CI [0.379, 0.402]; Fig. 4). The best algorithm for the stacked model was XGBoost. We outline results for each neuroimaging phenotype, each MRI modality, and each stacking algorithm in Supplementary Materials (S10).

**Figure 3.**
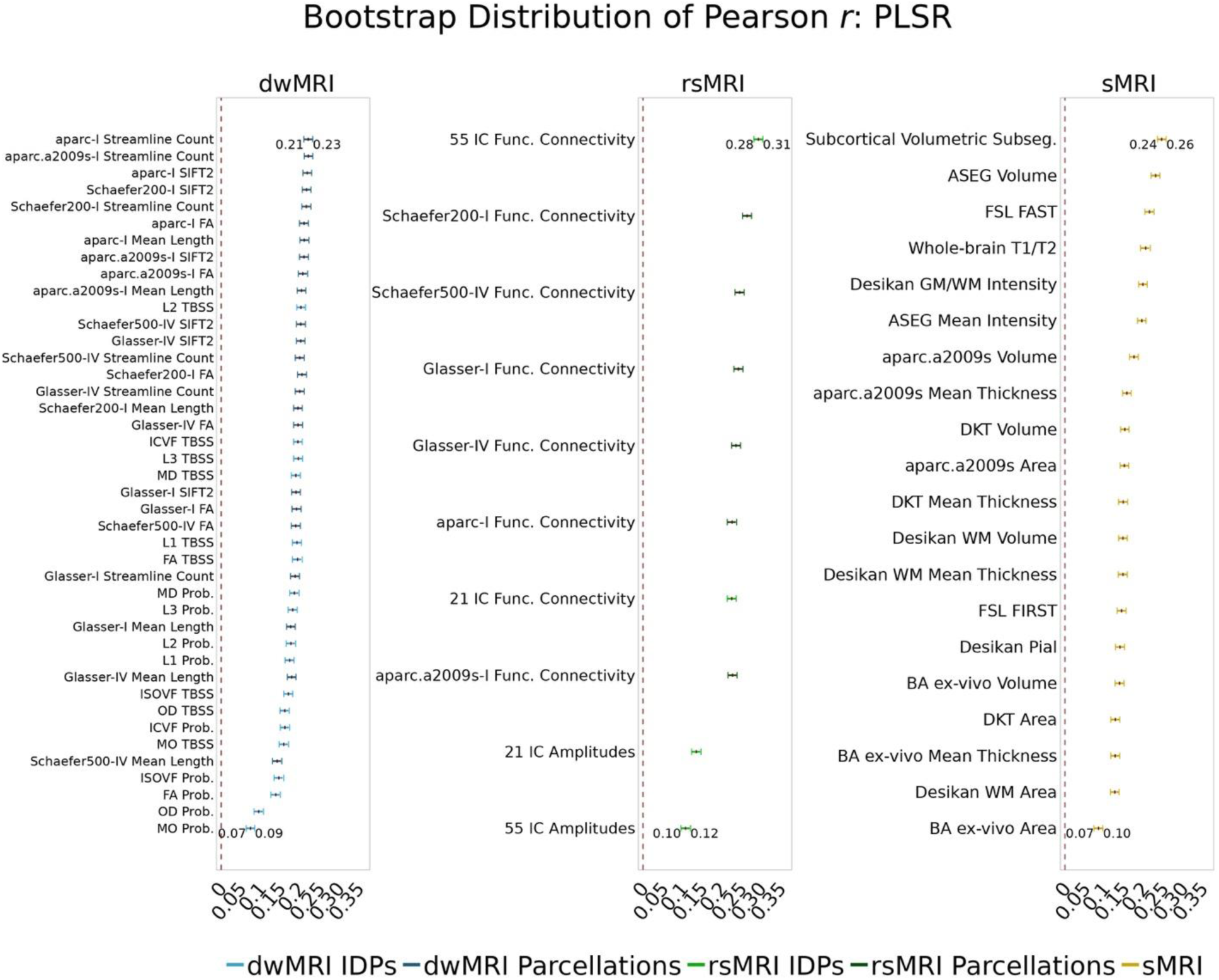
Predictive performance of machine learning models based on 72 individual neuroimaging phenotypes. Bootstrap distribution of Pearson *r* for the *g*-factor derived from ESEM and *g*-factor predicted from each neuroimaging phenotype, and corresponding 95% confidence intervals (95% CI). Values at the top and bottom of the plots indicate the lower and upper 95% CI for the bootstrap Pearson *r*.

**Figure 4.**
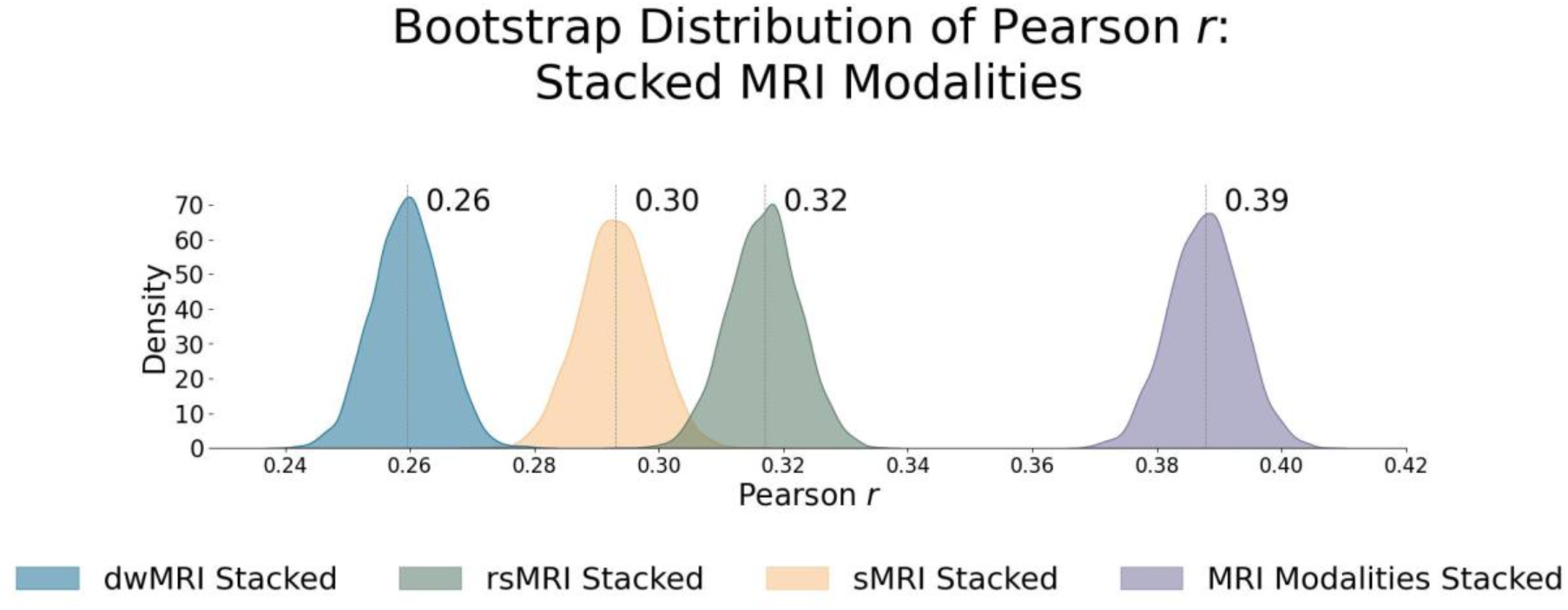
Predictive performance of machine learning models based on neuroimaging phenotypes stacked within and across three MRI modalities. Bootstrap distribution of Pearson *r* between the *g*-factor derived from ESEM and the *g*-factor predicted from stacked dwMRI, rsMRI, sMRI, and all MRI modalities stacked. Values at the top of each plot mark the median Pearson *r*.

#### dwMRI

Overall, models based on structural connectivity metrics performed better than TBSS and probabilistic tractography (Fig. 3). TBSS, in turn, performed better than probabilistic tractography (Fig. 3 and Table S13). The number of streamlines connecting brain areas parcellated with aparc MSA-I had the best predictive performance among all dwMRI neuroimaging phenotypes (*R*^2^_mean_ = 0.052, *r*_mean_ = 0.227, 95% CI [0.212, 0.235]). To identify features driving predictions, we correlated streamline counts in the aparc MSA-I parcellation with the predicted *g*-factor values from the PLSR model. Positive associations with the predicted *g*-factor were strongest for left superior parietal-left caudal anterior cingulate, left caudate-right amygdala, and left putamen-left hippocampus connections. The most marked negative correlations involved left putamen-right posterior thalamus and right pars opercularis-right caudal anterior cingulate pathways (Fig. 5 and Supplementary Fig. S2).

**Figure 5.**
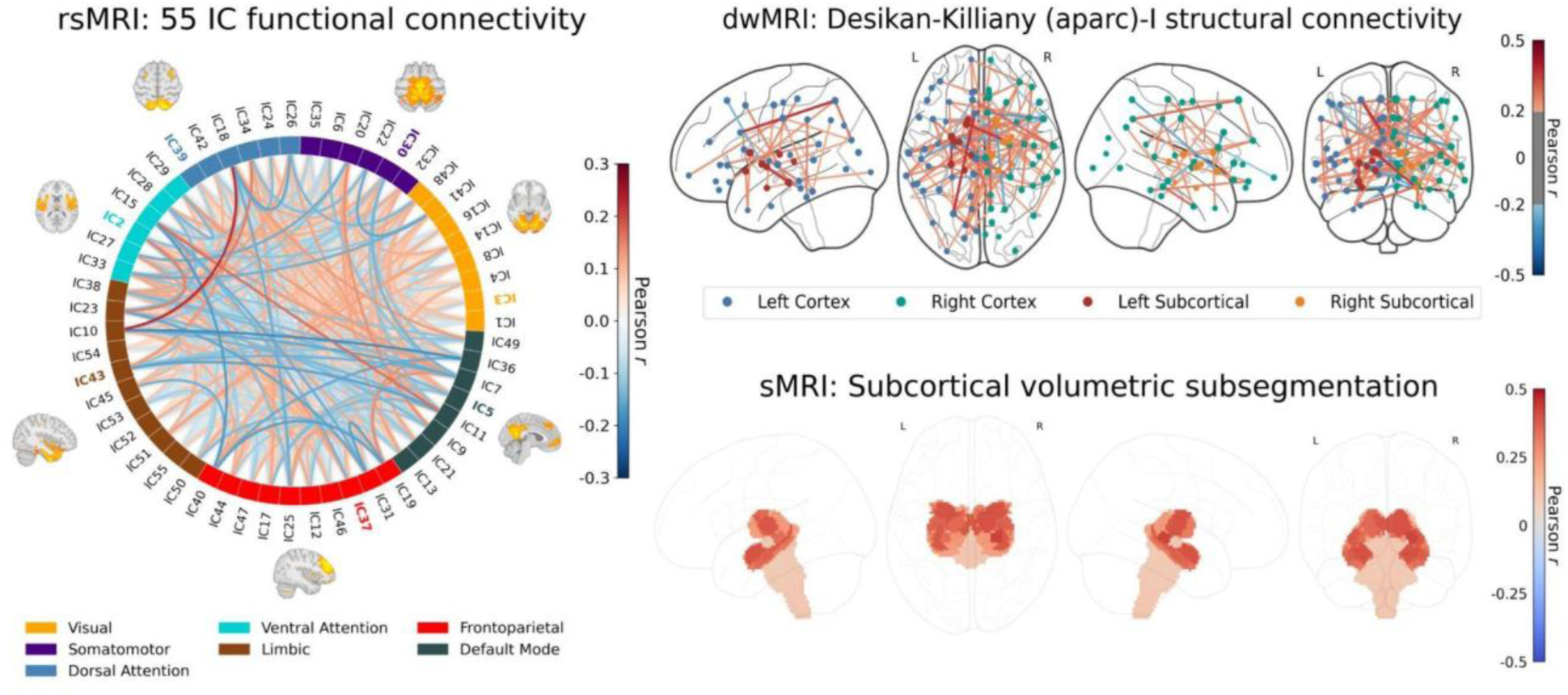
Feature importance maps for neuroimaging features with the highest predictive performance for cognition derived via the Haufe transformation [62]. The colour of the lines (rsMRI and dwMRI) and subcortical structures (sMRI) indicates the magnitude and direction of Pearson correlations between the predicted *g*-factor and features from the top-performing neuroimaging phenotype. Correlations were computed in test sets pooled across 5 outer folds. **rsMRI**: A connectogram displays network-level feature importance for functional connectivity between 55 neuronally driven independent components (IC) grouped into seven networks (Yeo 2011 parcellation). Full correlation matrices rather than tangent space parameterization were used for interpretability. The IC with the highest activation within each network is highlighted in colour, and its corresponding functional connectivity map is overlaid. Values of Pearson *r* for top correlations are given in Fig. S3. **dwMRI**: The importance of structural connections (streamline count) between brain regions parcellated using the aparc (Desikan-Killiany) MSA-I atlases for predicting cognition is shown as a glass brain plot, with cortical/subcortical nodes (circles) and their connecting edges (lines) coloured by correlation direction and strength. Values of Pearson *r* for top correlations are given in Fig. S2. **sMRI**: Regional volumes of subcortical structures derived from FreeSurfer subcortical volumetric subsegmentation are overlaid on a glass brain. Values of Pearson *r* for top correlations are given in Fig. S5.

The mean length of the streamlines connecting nodes from the Schaefer atlas for 500 cortical areas combined with MSA-IV had the lowest performance among all structural connectivity metrics (*R*^2^_mean_ = 0.018, *r*_mean_ = 0.145, 95% CI [0.132, 0.156]; Fig. 3). Among dwMRI IDPs, eigenvalue L2 from TBSS had the best predictive performance (*R*^2^_mean_ = 0.045, *r*_mean_ = 0.207, 95% CI [0.194, 0.216]), and MO and OD derived with probabilistic tractography were least predictive of the *g*-factor (*R*^2^_mean_ = 0.006, *r*_mean_ = 0.076, 95% CI [0.065, 0.087] and *R*^2^_mean_ = 0.01, *r*_mean_ = 0.099, 95% CI [0.085, 0.109], respectively).

Stacking *g*-factor values predicted from all dwMRI neuroimaging phenotypes improved the predictive performance of dwMRI to *R*^2^_mean_ = 0.073 and *r*_mean_ = 0.265 (95% CI [0.252, 0.273]; Fig. 4). The best algorithm for the stacked model was Random Forest.

#### rsMRI

Among RSFC metrics for 55 and 21 ICs, tangent parameterization matrices yielded the highest performance in the training set compared to full and partial correlation, as indicated by the cross-validation score. Functional connections between the limbic (IC10) and dorsal attention (IC18) networks, as well as between the ventral attention (IC15) and default mode (IC11) networks, displayed the highest positive association with cognition. In contrast, functional connectivity between the limbic (IC43, the highest activation within network) and default mode (IC11) and limbic (IC45) and frontoparietal (IC40) networks, between the dorsal attention (IC18) and frontoparietal (IC25) networks, and between the ventral attention (IC15) and frontoparietal (IC40) networks, showed the highest negative association with cognition (Fig. 5 and Supplementary Fig. S3 and S4).

Among RSFC metrics for parcellated time series data, full correlation matrices performed best in the training set. Overall, RSFC between 55 ICs quantified with tangent space parameterization had the highest predictive performance (*R*^2^_mean_ = 0.088, *r*_mean_ = 0.3, 95% CI [0.284, 0.307]), followed by RSFC between 200 cortical and 16 subcortical regions (Schaefer cortical atlas + MSA-I) measured with full correlation (*R*^2^_mean_ = 0.07, *r*_mean_ = 0.27, 95% CI [0.255, 0.278]). The predictive performance of 21 and 55 ICs amplitudes was the lowest (*R*^2^_mean_ = 0.013, *r*_mean_ = 0.14, 95% CI [0.098, 0.122] and *R*^2^_mean_ = 0.019, *r*_mean_ = 0.11, 95% CI [0.125, 0.149], respectively; Fig. 3 and Table S14).

Stacking *g*-factor values predicted from rsMRI neuroimaging phenotypes considerably improved the predictive performance of rsMRI to *R*^2^_mean_ = 0.105 and *r*_mean_ = 0.325 (95% CI [0.308, 0.331]; Fig. 4 and Table S14). Similar to dwMRI, the best algorithm in the stacked model was Random Forest.

#### sMRI

FreeSurfer subcortical volumetric subsegmentation and ASEG had the highest performance among all sMRI neuroimaging phenotypes (*R*^2^_mean_ = 0.068, *r*_mean_ = 0.244, 95% CI [0.237, 0.259] and *R*^2^_mean_ = 0.059, *r*_mean_ = 0.235, 95% CI [0.221, 0.243], respectively). In FreeSurfer subcortical volumetric subsegmentation, volumes of all subcortical structures, except for left and right hippocampal fissures, showed positive associations with cognition. The strongest relations were observed for the volumes of bilateral whole hippocampal head and whole hippocampus (Fig. 5 and Supplementary Fig. S5 for feature importance maps). Grey matter morphological characteristics from *ex vivo* Brodmann Area Maps showed the lowest predictive performance (*R*^2^_mean_ = 0.008, *r*_mean_ = 0.089, 95% CI [0.075, 0.098]; Fig. 3 and Table S15).

Stacking *g*-factor values predicted from all sMRI neuroimaging phenotypes improved the model’s predictive performance to *R*^2^_mean_ = 0.095 and *r*_mean_ = 0.298 (95% CI [0.284, 0.307]). The best algorithm in the stacked model was Support Vector Regression (Fig. 4 and Table S16).

### Commonality Analysis

Different neuroimaging phenotypes captured the relationship between cognition and mental health at varying degrees, as indicated by a percentage ratio between the common effect of mental health-*g* and neuroimaging-*g* and the total effect of mental health-*g*. Neuroimaging phenotypes from dwMRI, rsMRI, and sMRI accounted for 2.1%–19.3%, 4%–25.8%, and 4.8%–21.8% of the cognition–mental health relationship, respectively (Fig. 6 and Tables S18 and S19). Among dwMRI neuroimaging phenotypes, the number of streamlines connecting grey matter regions from Destrieux (aparc.a2009s) cortical + MSA-I subcortical parcellations shared the highest proportion of variance with mental health-*g* (19.3%). For rsMRI, the largest proportion of the common effect of mental health-*g* was shared with RSFC between 55 ICs (25.8%). For sMRI, subcortical volumetric subsegmentation contributed most to the link between cognition and mental health (21.8%). The correlation between the performance of each neuroimaging phenotype in predicting cognition and the proportion of the relationship between cognition and mental health captured by the phenotype was *r* = 0.97 (95% CI [0.958, 0.982]; Fig. 7).

When we stacked neuroimaging phenotypes within dwMRI, rsMRI, and sMRI, we captured 25.5%, 29.8%, and 31.6% of the predictive relationship between cognition and mental health, respectively. By stacking all 72 neuroimaging phenotypes across three MRI modalities, we enhanced the explanation to 48% (Fig. 8e–h and Table S20).

**Figure 6.**
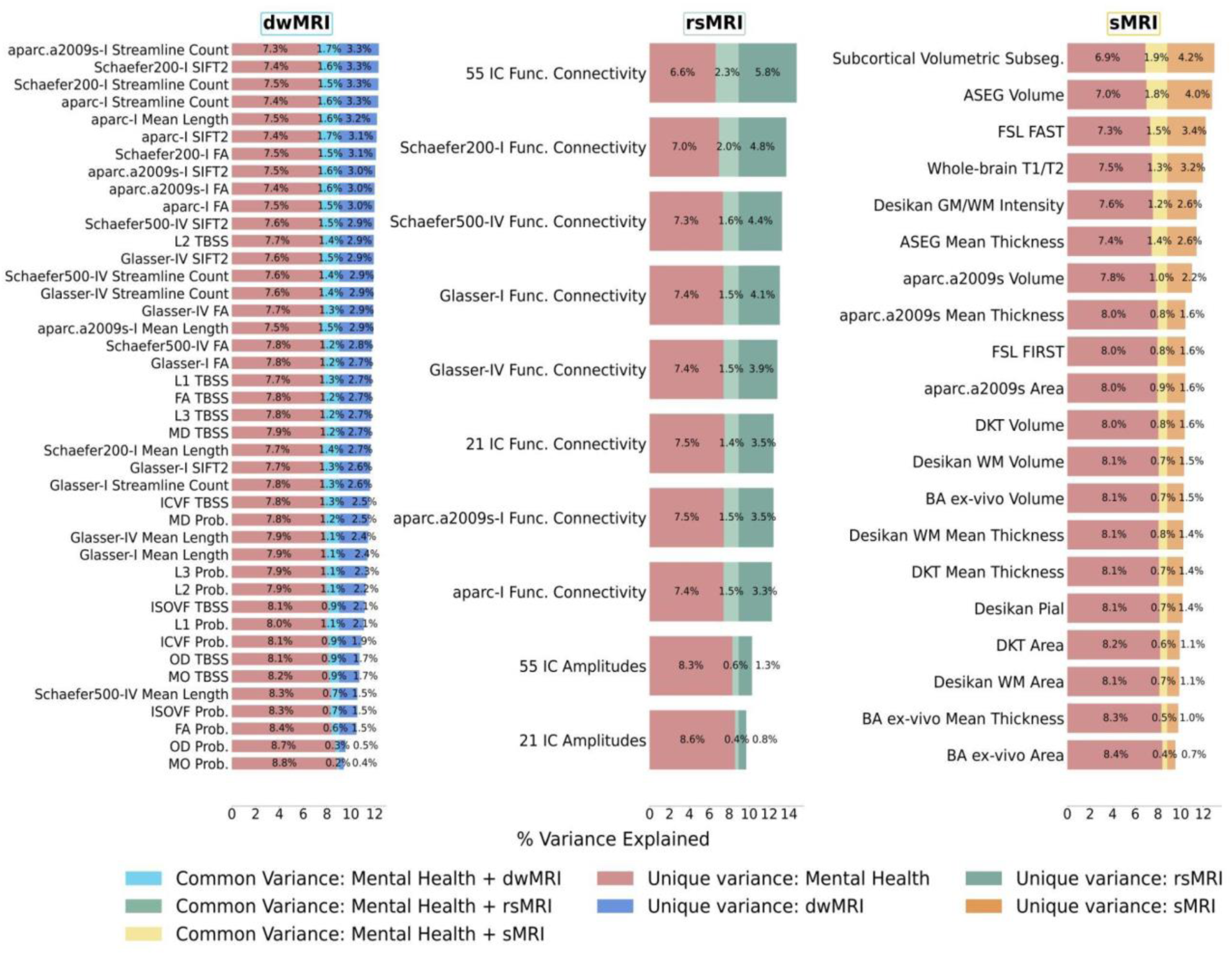
Results of commonality analyses: the contribution of neuroimaging phenotypes to the relationship between cognition and mental health. Stacked bar plot diagrams of the results of commonality analyses for each neuroimaging phenotype. *Unique variance* proportion (%) of variance in the *g*-factor explained uniquely by mental health and neuroimaging phenotypes, *Common variance* proportion (%) of variance in the *g*-factor shared between mental health and neuroimaging phenotypes. *aparc.a2009s*, Destrieux cortical atlas; *Schaefer7n200p*, Schaefer Atlas for 7 networks and 200 parcels; *Schaefer7n500p*, Schaefer Atlas for 7 networks and 500 parcels; *I*, Melbourne Subcortical Atlas I; *IV*, Melbourne Subcortical Atlas IV; *SIFT2*, Spherical-Deconvolution Informed Filtering of Tractograms 2; *FA*, fractional anisotropy; *MD*, mean diffusivity; *MO*, diffusion tensor mode; *L1, L2, and L3*, eigenvalues of the diffusion tensor; *OD*, orientation dispersion index; *ICVF*, intracellular volume fraction; *ISOVF*, isotropic volume fraction; *TBSS*, Tract-Based Spatial Statistics; *Func. Connectivity*, functional connectivity; *Subseg*., Freesurfer subsegmentation; *ASEG*, Freesurfer automated subcortical volumetric segmentation; *FSL FAST*, FSL FMRIB’s Automated Segmentation Tool; *WM*, white matter; *GM*, grey matter; *FSL*, *FIRST* FMRIB’s Integrated Registration and Segmentation Tool; *DKT*, Desikan-Killiany-Tourville; *BA*, FreeSurfer *ex-vivo* Brodmann Area Maps.

**Figure 7.**
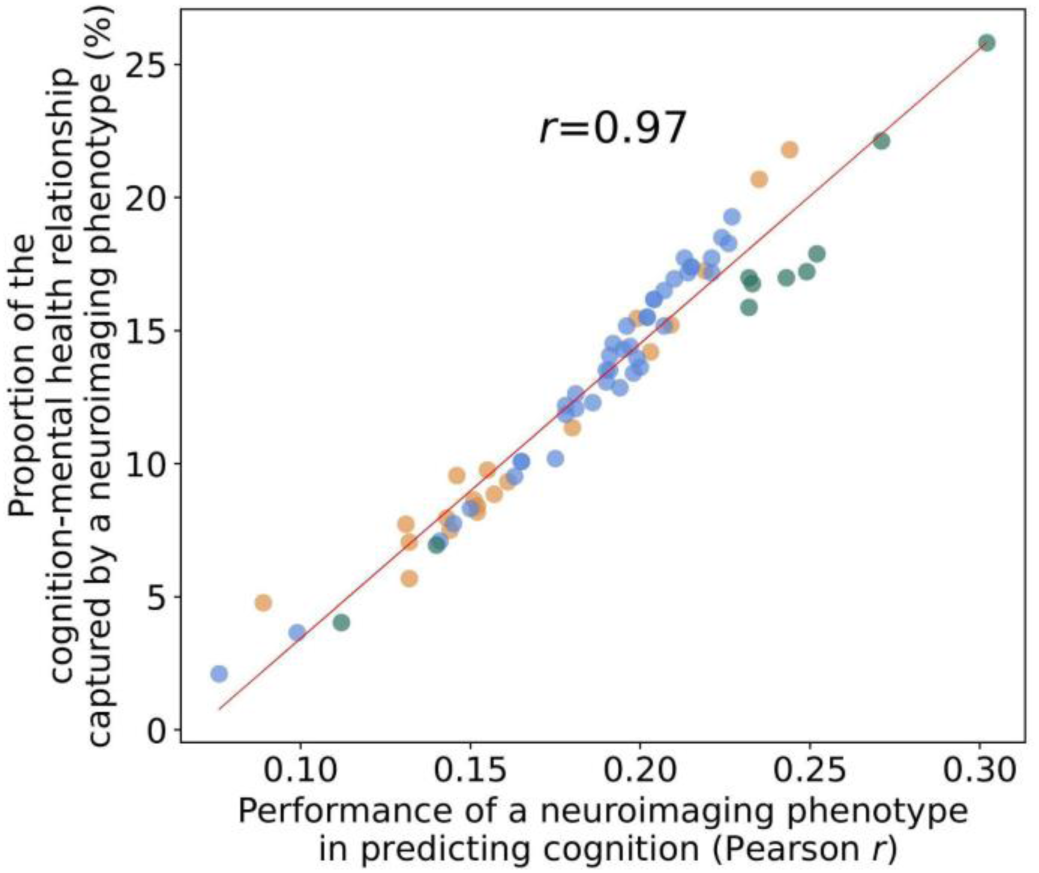
Scatterplot of the relationship between the PLSR performance of individual neuroimaging phenotypes and the proportion of the cognition–mental health relationship they capture.

Age and sex shared substantial overlapping variance with both mental health and neuroimaging in explaining cognition, accounting for 43% of the variance in the cognition–mental health relationship. Multimodal neural marker of cognition based on three MRI modalities (“All MRI Stacked”) explained 72% of this age and sex-related variance (Fig. 8i–l and Table S21).

**Figure 8.**
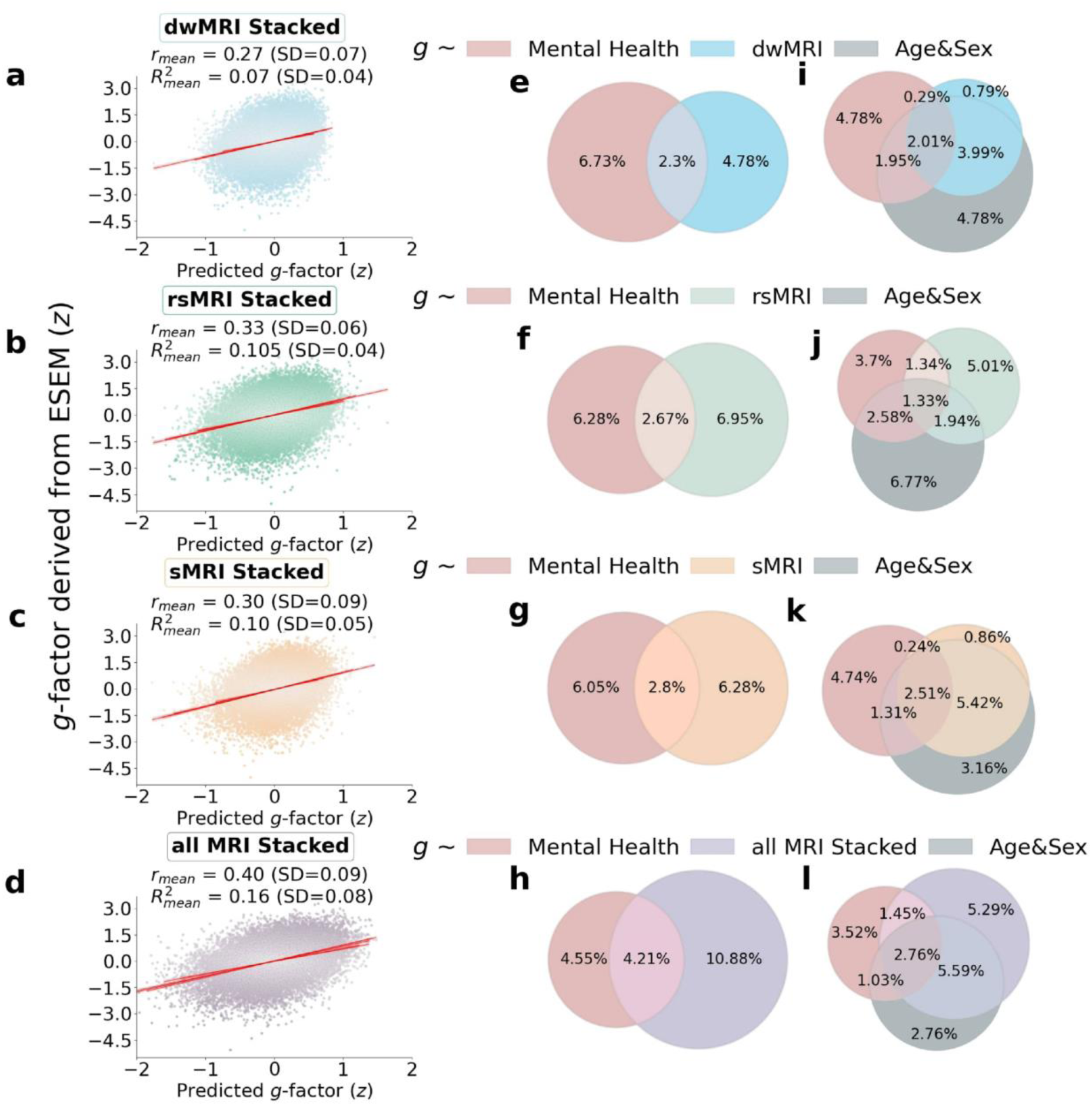
The contribution of neuroimaging phenotypes stacked within each and across all MRI modalities to the relationship between cognition and mental health: Results of predictive modeling and commonality analyses. **a-d** Distributions of the *g*-factor derived from cognitive tests via ESEM and predicted from stacked dwMRI **(a)**, rsMRI **(b)**, sMRI **(c)**, and all MRI modalities **(d)**. **e-l** Venn diagrams of the results of commonality analyses. **e-h** The proportion (%) of variance in the *g*-factor explained uniquely by mental health and neuroimaging phenotypes: dwMRI **(e)**, rsMRI **(f)**, sMRI **(g)**, and all MRI modalities stacked **(h)**, as well as the common effects between mental health and neuroimaging phenotypes. **i-l** The proportion (%) of variance in the *g*-factor explained uniquely by mental health, neuroimaging phenotypes stacked within dwMRI **(i),** rsMRI **(j)**, sMRI **(k)**, and all MRI modalities **(l)**, and age and sex, as well as the common effects among all explanatory variables.

## Discussion

Our study is the first to quantify the contribution of the neural indicators of cognition, as reflected by the *g*-factor, to its relationship with mental health in the largest population-level cohort. We show that the performance of each neural indicator in predicting cognition *per se* is strongly related to its ability to explain the link between cognition and mental health. In other words, the “robustness” of the neural indicator of cognition is associated with its capacity to capture the covariation between cognition and mental health. This means that the neural indicators that capture more of the individual differences in cognition also capture more of the cognition–mental health covariation. We further demonstrate that information from 72 neuroimaging phenotypes from three MRI modalities accounts for almost half of the variance in the relationship between cognition and mental health. Aspects of the brain that underpin this relationship are best reflected in neuroimaging phenotypes derived from rsMRI, followed by sMRI and dwMRI. For rsMRI, RSFC between 55 large-scale networks measured with tangent space parameterization was the best-performing neuroimaging phenotype. Among sMRI neuroimaging phenotypes, subcortical grey matter characteristics quantified using FreeSurfer’s subcortical volumetric subsegmentation, and ASEG shared the highest proportion of variance that mental health captures. For dwMRI, microstructural properties of the brain’s white matter that underlie the link between cognition and mental health were best reflected in the number of streamlines connecting grey matter regions from Destrieux cortical + MSA I subcortical parcellations. Integrating information from neuroimaging phenotypes within and across MRI modalities enhanced both the prediction of cognition and the explanation of the relationship between cognition and mental health. We discuss the results in the context of current knowledge in the field, as follows.

### The cognition and mental health relationship

Our analysis confirmed the validity of the *g*-factor [31] as a quantitative measure of cognition [31], demonstrating that it captures nearly half (39%) of the variance across twelve cognitive performance scores, consistent with prior studies [63–68]. Furthermore, we were able to predict cognition from 133 mental health indices, showing a medium-sized relationship that aligns with existing literature [69,70]. Although the observed mental health-cognition association is lower than within-sample estimates in conventional regression models, it aligns with our prior mega-analysis in children [69]. Notably, this effect size is not considered small in gerontology. In fact, it falls around the 70^th^ percentile of reported effects and approaches the threshold for a large effect at *r* = 0.32 [71]. While we focused specifically on cognition as an RDoC core domain, the strength of its relationship with mental health may be bounded by the influence of other functional domains, particularly in normative, non-clinical samples – a promising direction for future research.

The directions of PLSR loadings were broadly consistent with univariate correlations. PLSR extends beyond univariate approaches by modelling multivariate relationships across features and outcomes. Consistently, both univariate correlations and factor loadings derived from the PLSR model indicated that scores for mental distress, alcohol and cannabis use, and self-harm behaviours related positively, and the scores for anxiety, neurological and mental health diagnoses, unusual or psychotic experiences, happiness and subjective well-being, and negative traumatic events related negatively to the *g*-factor. Positive PLSR loadings of features related to mental distress may indicate greater susceptibility to or exaggerated perception of stressful events, psychological overexcitability, and predisposition to rumination in people with higher cognition [72]. On the other hand, these findings may be specific to the UK Biobank cohort and the way the questions for this mental health category were constructed. In particular, to evaluate mental distress, the UK Biobank questionnaire asked whether an individual sought or received medical help for or suffered from mental distress. In this regard, the estimate for mental distress may be more indicative of whether an individual experiencing mental distress had an opportunity or aspiration to visit a doctor and seek professional help [73]. Thus, people with better cognitive abilities and also with a higher socioeconomic status may indeed be more likely to seek professional help.

Limited evidence supports a positive association between self-harm behaviours and cognitive abilities, with some studies indicating higher cognitive performance as a risk factor for non-suicidal self-harm. Research shows an inverse relationship between cognitive control of emotion and suicidal behaviours that weakens over the life course [73,74]. Some studies have found a positive correlation between cognitive abilities and the risk of non-suicidal self-harm, suicidal thoughts, and suicidal plans that may be independent of or, conversely, affected by socioeconomic status [75,76]. In our study, the magnitude of the association between self-harm behaviours and cognition was low (Fig. 2), indicating a weak relationship.

Positive PLSR loadings of features related to alcohol and cannabis may also indicate the influence of other factors. Overall, this relationship is believed to be largely affected by age, income, education, social status, social equality, social norms, and quality of life [79–80]. For example, education level and income correlate with cognitive ability and alcohol consumption [79,81–83]. Research also links a higher probability of having tried alcohol or recreational drugs, including cannabis, to a tendency of more intelligent individuals to approach evolutionary novel stimuli [84,85]. This hypothesis is supported by studies showing that cannabis users perform better on some cognitive tasks [86]. Alternatively, frequent drinking can indicate higher social engagement, which is positively associated with cognition [87]. Young adults often drink alcohol as a social ritual in university settings to build connections with peers [88]. In older adults, drinking may accompany friends or family visits [89,90]. Mixed evidence on the link between alcohol and drug use and cognition makes it difficult to draw definite conclusions, leaving an open question about the nature of this relationship.

Consistent with previous studies, we showed that anxiety and negative traumatic experiences were inversely associated with cognitive abilities [90–93]. Anxiety may be linked to poorer cognitive performance via reduced working memory capacity, increased focus on negative thoughts, and attentional bias to threatening stimuli that hinder the allocation of cognitive resources to a current task [94–96]. Individuals with PTSD consistently showed impaired verbal and working memory, visual attention, inhibitory function, task switching, cognitive flexibility, and cognitive control [97–100]. Exposure to traumatic events that did not reach the PTSD threshold was also linked to impaired cognition. For example, childhood trauma is associated with worse performance in processing speed, attention, and executive function tasks in adulthood, and age at a first traumatic event is predictive of the rate of executive function decline in midlife [101,102]. In the UK Biobank cohort, adverse life events have been linked to lower cognitive flexibility, partially via depression level [103].

In agreement with our findings, cognitive deficits are often found in psychotic disorders [104,105]. We treated neurological and mental health symptoms as predictor variables and did not stratify or exclude people based on psychiatric status or symptom severity. Since no prior studies have examined isolated psychotic symptoms (e.g., recent unusual experiences, hearing unreal voices, or seeing unreal visions), we avoid speculating on how these symptoms relate to cognition in our sample.

Finally, both negative PLSR loadings and corresponding univariate correlations for features related to happiness and subjective well-being may be specific to the study cohort, as these findings do not agree with some previous research [107–109]. On the other hand, our results agree with the study linking excessive optimism or optimistic thinking to lower cognitive performance in memory, verbal fluency, fluid intelligence, and numerical reasoning tasks, and suggesting that pessimism or realism indicates better cognition [110]. The concept of realism/optimism as indicators of cognition is a plausible explanation for a negative association between the *g*-factor and friendship satisfaction, as well as a negative PLSR loading of feelings that life is meaningful, especially in older adults who tend to reflect more on the meaning of life [111]. The latter is supported by the study showing a negative association between cognitive function and the search for the meaning of life and a change in the pattern of this relationship after the age of 60 [112]. Finally, a UK Biobank study found a positive association of happiness with speed and visuospatial memory but a negative relationship with reasoning ability [113].

### How well does MRI capture the predictive relationship between cognition and mental health?

Consistent with previous studies, we show that MRI data predict individual differences in cognition with a medium-sized performance (*r* ≈ 0.4) [15–17, 28, 65, 114, 115]. This provides us confidence in using MRI to derive quantitative neuromarkers of cognition. Neural indicators of cognition derived from rsMRI and sMRI neuroimaging phenotypes explain approximately a third of this link (29.8% and 31.6%, respectively), whereas multimodal neural indicators of cognition derived from all neuroimaging phenotypes account for almost half (48%) of the variance in the cognition–mental health relationship. Yet, combining all neuroimaging phenotypes from three MRI modalities allowed us to explain the highest proportion of the variance in cognition that mental health captures.

Among dwMRI-derived neuroimaging phenotypes, models based on structural connectivity between brain areas parcellated with aparc MSA-I (streamline count), particularly connections with bilateral caudal anterior cingulate (left superior parietal-left caudal anterior cingulate, right pars opercularis-right caudal anterior cingulate), left putamen (left putamen-left hippocampus, left putamen-right posterior thalamus), and amygdala (left caudate-right amygdala), result in a neural indicator that best reflects microstructural resources associated with cognition, as indicated by predictive modeling, and more importantly, shares the highest proportion of the variance with mental health-*g*, as indicated by commonality analysis. One of the mechanisms that can be reflected in the link between streamline count and individual variations in cognition is strengthening local connections within a hemisphere and enhancing local (i.e., the connectivity between neighbouring regions) and nodal efficiency (i.e., how well a given brain region is connected to other regions) [116]. The somewhat limited utility of diffusion metrics derived specifically from probabilistic tractography in serving as robust quantitative neuromarkers of cognition and its shared variance with mental health may stem from their greater sensitivity and specificity to neuronal integrity and white matter microstructure rather than to dynamic cognitive processes. Critically, probabilistic tractography may be less effective at capturing relationships between white matter microstructure and behavioural scores cross-sectionally, as this method is more sensitive to pathological changes or dynamic microstructural alterations like those occurring during maturation. While these indices can capture abnormal white matter microstructure in clinical populations such as Alzheimer’s disease, schizophrenia, or attention deficit hyperactivity disorder (ADHD) [117–119], the empirical evidence on their associations with cognitive performance is controversial [114, 120–126].

We extend findings on the superior performance of rsMRI in predicting cognition, which aligns with the literature [15, 28], by showing that it also explains almost a third of the variance in cognition that mental health captures. At the rsMRI neuroimaging phenotype level, this performance is mostly driven by RSFC patterns among 55 ICA-derived networks quantified using tangent space parameterization. At a feature level, these associations are best captured by the strength of functional connections among limbic, dorsal attention and ventral attention, frontoparietal and default mode networks. These functional networks have been consistently linked to cognitive processes in prior research [127–130].

ICA is a data-driven technique that does not rely on predefined anatomical boundaries. It captures intrinsic large-scale functional networks accounting for individual variability in RSFC. Thus, by providing more “personalized” connectivity representations, ICA-derived networks may be more robust neuronal indicators of cognitive performance than node-to-node estimates [131, 132]. Furthermore, using tangent space parametrization to quantify RSFC not only improves the predictive performance of rsMRI for cognition, as shown previously [55, 133–136], but also allows us to capture more variance that cognition shares with mental health. Although tangent parametrization matrices do not reflect individual functional brain connections and cannot be interpreted directly, the linearization and projection to Euclidean space make functional connectivity estimates more suitable for statistical analysis and machine learning [54]. Resting-state fluctuation amplitudes from rsMRI, which are the least predictive of cognition and explain the smallest proportion of the variance in cognition that mental health captures, are believed to reflect cardiovascular and cerebrovascular factors distinct from neural effects [137]. Research indicates that network amplitudes are significantly influenced by age, cardiovascular and lung function, and other physical measures, and are therefore subject to high interindividual variability. Consequently, interindividual variability in vascular health and aging dynamics may confront the capacity of network amplitudes to serve as a robust neural marker of cognition [52].

Integrating information about brain anatomy by stacking sMRI neuroimaging phenotypes allowed us to explain a third of the link between cognition and mental health. Among all sMRI neuroimaging phenotypes, those that quantified the morphology of subcortical structures, particularly volumes of bilateral hippocampus and hippocampal head, explain the highest portion of the variance in cognition captured by mental health. Our findings show that, at least in older adults, volumetric properties of subcortical structures are not only more predictive of individual variations in cognition but also explain a greater portion of cognitive variance shared with mental health than structural characteristics of more distributed cortical grey and white matter. This aligns with the Scaffolding Theory that proposes stronger compensatory engagement of subcortical structures in cognitive processing in older adults [138–140].

The study has several limitations. First, the UK Biobank MRI data include only one task for task functional MRI (tfMRI), the Hariri hammer task [141], which is not designed to be cognitively demanding. Compared to other MRI modalities, tfMRI from certain tasks, such as the *n*-back working memory task [142], has been shown to provide the most robust neuromarker of cognition [17]. Thus, by not including cognitively demanding tfMRI tasks in predictive models, we may have missed condition-specific variance that may account for a substantial portion of the variance specific to particular cognitive domains. Second, for generalizability purposes, we did not stratify the cohort nor exclude individuals with neurological, cardiovascular, or any other clinically established disorders, assuming that, in older adults, these effects can be tightly intertwined with neuronal mechanisms that sustain cognitive processing. Finally, the UK Biobank’s mental health questionnaire and cognitive test battery may miss important information as they cover a limited and specific set of neuropsychiatric conditions and cognitive domains, respectively. For example, the mental health questionnaire does not include questions evaluating autism or ADHD symptoms. The UK Biobank’s cognitive test battery was designed specifically for the study and is different from commonly used cognitive test batteries such as the NIH Toolbox for Assessment of Neurological and Behavioral Function [143] used in the Human Connectome Project [144] and Adolescent Brain Cognitive Development Study [145] or Wechsler Adult Intelligence Scale [146] used in the Dunedin Study [147], which hinders the cross-study comparison of the cognitive performance score.

Overall, our findings suggest that RSFC is a more fine-tuned system that exhibits a degree of flexibility and variability that is not entirely constrained by anatomical pathways, as cortical regions can use alternative or parallel pathways to strengthen or weaken functional connections underlying cognitive processing [148]. Although RSFC is believed to reflect anatomical connectivity [148, 149], alterations in structural connectivity do not always compromise RSFC [149]. From this point, a pattern of RSFC maintained for an extended period may eventually cause concurrent alterations in cognition and mental health, especially if it involves brain areas that have overlapping effects on both. Still, the physical integrity and morphology of the structures providing a “physical” relay for functionally connected brain areas to communicate play a pivotal role in the brain-behaviour relationship. Although more rigid and less flexible in an adult brain, they determine the amount of neural resources available for cognitive processing. Nevertheless, none of the neuroimaging phenotypes or MRI modalities alone is sufficient to provide a complete picture of the complex relationship between cognition and mental health, and combining all sources of information about brain structure and function reflected in three MRI modalities allows us to derive robust quantitative neuromarkers of cognition and boost the explanation of neural correlates that underlie this link.

In line with the National Institute of Mental Health’s RDoC framework, we shed light on the relationship between cognition and mental health as one of the six transdiagnostic spectrums of neuropsychiatric symptoms at one of the seven units of analysis, i.e., the neural level [150]. This may serve as a methodological “bridge” linking other units of analysis, such as genes and behaviour. By elucidating the role of different neuroimaging phenotypes as neural correlates of cognition that overlap with mental health, we provide potential targets for behavioural and physiological interventions that may affect cognition.

Although recent debates [18] have challenged the predictive utility of MRI for cognition, our multimodal marker integrating 72 neuroimaging phenotypes captures nearly half of the mental health-explained variance in cognition. We demonstrate that neural markers with greater predictive accuracy for cognition also better explain cognition–mental health covariation, showing that multimodal MRI can capture both a substantial cognitive variance and nearly half of its shared variance with mental health. Finally, we show that our neuromarkers explain a substantial portion of the age- and sex-related variance in the cognition–mental health relationship, highlighting their relevance in modeling cognition across demographic strata.

The remaining unexplained variance in the relationship between cognition and mental health likely stems from multiple sources. One possibility is the absence of certain neuroimaging modalities in the UK Biobank dataset, such as task-based fMRI contrasts, positron emission tomography, arterial spin labeling, and magnetoencephalography/electroencephalography. Prior research has consistently demonstrated strong predictive performance from specific task-based fMRI contrasts, particularly those derived from tasks like the *n*-Back working memory task and the face-name episodic memory task, none of which is available in the UK Biobank [15,17,61,69,114,142,151].

Moreover, there are inherent limitations in using MRI as a proxy for brain structure and function. Measurement error and intra-individual variability, such as differences in a cognitive state between cognitive assessments and MRI acquisition, may also contribute to the unexplained variance. According to the RDoC framework, brain circuits represent only one level of neurobiological analysis relevant to cognition [14]. Other levels, including genes, molecules, cells, and physiological processes, may also play a role in the cognition–mental health relationship.

Neuroimaging offers a unique window into the biological mechanisms underlying cognition–mental health overlap – insights unattainable from behavioural data alone. Our findings validate brain-based neural markers as a core unit of analysis for cognitive functioning, advancing mental health research through the lens of cognition. Beyond this conceptual contribution, the study has clinical implications. First, by demonstrating a transdiagnostic link between cognition and mental health, we support interventions that enhance cognition as a pathway to improving mental health. Second, we show neuroimaging as an effective tool for assessing the neurobiological basis of this link. Quantifying neuroimaging’s capacity to capture this relationship is essential for future research integrating imaging with cognitive testing to monitor treatment-related neural changes. Such work could enable personalised interventions, using neuroimaging to track cognitive changes and treatment efficacy (e.g., stimulant medications for ADHD) aimed at boosting cognitive functioning.

## Data availability

The data used in this study are subject to restrictions. Access requests should be directed to the UK Biobank (https://www.ukbiobank.ac.uk/).

## Code availability

The codes for analysing the data are available at https://github.com/HAM-lab-Otago-University/UKBiobank/.

## Data Availability

https://github.com/HAM-lab-Otago-University/UKBiobank/

## Acknowledgments

This research has been conducted using the UK Biobank Resource under Application Number 70132. We are thankful to Professor Bruce Russell for his assistance with access to the UK Biobank dataset.

N.P. and J.D. were supported by Health Research Council of New Zealand (grant numbers 21/618 and 24/838) and by Neurological Foundation of New Zealand (grant number 2350 PRG). N.P. was supported by the Ministry of Business, Innovation and Employment (grant numbers UOA2421 and RTVU2403). I.B. was supported by the University of Otago.

