## Supplementary Information for "Multimodal MRI Marker of Cognition Explains the Association Between Cognition and Mental Health in UK Biobank"

### Supplementary Methods

#### S1. Cognition

##### Description of core measures

We used twelve scores from the eleven cognitive tests that represented the following cognitive domains: reaction time and processing speed (Reaction Time test), working memory (Numeric Memory test), verbal and numerical reasoning (Fluid Intelligence test), executive function (Trail Making Test), non-verbal fluid reasoning (Matrix Pattern Completion test), processing speed (Symbol Digit Substitution test), vocabulary (Picture Vocabulary test), planning abilities (Tower Rearranging test), verbal declarative memory (Paired Associate Learning test), prospective memory (Prospective Memory test), and visual memory (Pairs Matching test) [1].

**Table S1.** Cognitive tests and core measures of the UK Biobank cognitive test battery used in the study

| Test | Cognitive domain | Core measures | Field ID |
|---|---|---|---|
| **Reaction Time** | Reaction time and processing speed | Mean time to correctly identify matches | 20023 |
| **Numeric Memory** | Working memory | Maximum digits remembered correctly | 4282 |
| **Fluid Intelligence** | Verbal and numerical reasoning | Fluid intelligence score | 20016 |
| **Prospective Memory** | Prospective memory | Initial answer | 4292 |
| **Trail Making** | Executive function | Duration to complete numeric path (trail 1)<br>Duration to complete alphabetic path (trail 2) | 6348<br>6350 |

| | | | |
|---|---|---|---|
| **Matrix Pattern Completion** | Non-verbal fluid reasoning | Number of puzzles correctly solved | 6373 |
| **Symbol Digit Substitution** | Processing speed | Number of symbol digit matches made correctly | 23324 |
| **Picture Vocabulary** | Vocabulary (crystallized cognitive ability) | Specific cognitive ability | 26302 |
| **Tower Rearranging** | Planning abilities (a component of executive function) | Number of puzzles correct | 21004 |
| **Paired Associate Learning** | Verbal declarative memory | Number of word pairs correctly associated | 20197 |
| **Pairs Matching** | Visual memory | Number of incorrect matches in round | 399 |

#### Transformations of cognitive scores

To eliminate skewness, we $\log_x$-transformed the mean time to correctly identify matches in the Reaction Time test and duration to complete numeric and alphabetic trails in the Trail Making test [1–3]. For the Pairs Matching task, as the outcome measure, we selected and $\log_{x+1}$-transformed the number of incorrect matches made in the six-pair version of the test because of greater variability of the values and a very low rate of incorrect matches in the three-pair version of the test [1]. In the Prospective Memory test, we scored the participants as zero if they did not complete the task on the first attempt, i.e., touched a shape different from the target shape (orange circle), and as one if they correctly selected the target shape at the initial attempt. Correspondingly, we replaced values that encode all shapes except the target shape (0 – blue square, 1 – pink star, 2 – grey cross) with 0 and the value that encoded the target shape (3 – orange circle) with 1. We also replaced negative values with NA as they encoded invalid trials or trials abandoned by a participant, and zeroes with NA in the tests where a zero score would indicate an invalid trial (e.g., Fluid Intelligence score, specific cognitive ability in Picture Vocabulary test, and duration to complete trail in Trail Making test).

**Table S2.** Whole-sample distributions of cognitive performance scores used to derive the *g*-factor (N = 31 614)

| No | Variable | Statistics / Values | Frequencies | Distribution Plot |
|---|---|---|---|---|

| | | | | |
|---|---|---|---|---|
| **1** | **(log$_x$)Reaction time** | Mean (SD): 6.4 (0.2)<br>min < med < max:<br>5.8 < 6.4 < 7.4<br>IQR (CV): 0.2 (0) | 550 distinct values | 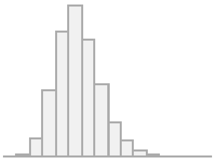 |
| **2** | **Fluid intelligence score** | Mean (SD): 6.6 (2)<br>min < med < max:<br>1 < 7 < 13<br>IQR (CV): 3 (0.3) | 13 distinct values | 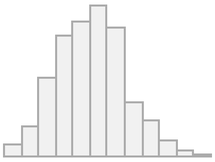 |
| **3** | **Numeric memory: Maximum digits remembered correctly** | Mean (SD): 6.8 (1.3)<br>min < med < max:<br>2 < 7 < 12<br>IQR (CV): 2 (0.2) | 11 distinct values | 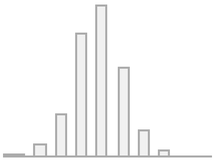 |
| **4** | **(log$_x$)Trail making: Duration to complete numeric path** | Mean (SD): 5.4 (0.3)<br>min < med < max:<br>4.5 < 5.3 < 7.5<br>IQR (CV): 0.3 (0.1) | 638 distinct values | 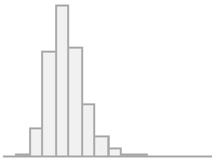 |
| **5** | **(log$_x$)Trail making: Duration to complete alphabetic path** | Mean (SD): 6.3 (0.4)<br>min < med < max:<br>5.1 < 6.2 < 8.7<br>IQR (CV): 0.5 (0.1) | 1542 distinct values | 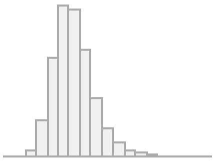 |
| **6** | **Symbol digit substitution: Number of correct matches** | Mean (SD): 18.9 (5.2)<br>min < med < max:<br>0 < 19 < 37<br>IQR (CV): 7 (0.3) | 38 distinct values | 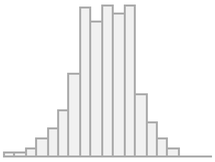 |
| **7** | **Paired associate learning: Number of correct pairs** | Mean (SD): 7 (2.5)<br>min < med < max:<br>0 < 7 < 10<br>IQR (CV): 4 (0.4) | 11 distinct values | 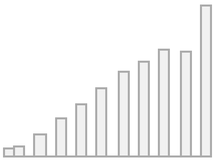 |
| **8** | **Tower rearranging: Number of puzzles correct** | Mean (SD): 9.9 (3.2)<br>min < med < max:<br>0 < 10 < 18<br>IQR (CV): 4 (0.3) | 19 distinct values | 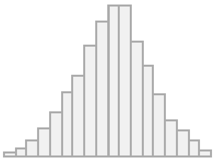 |
| **9** | **Matrix pattern completion: Number of puzzles correct** | Mean (SD): 8 (2.1)<br>min < med < max:<br>0 < 8 < 15<br>IQR (CV): 2 (0.3) | 16 distinct values | 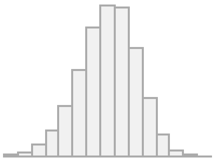 |
| **10** | **(log$_{x+1}$)Pairs matching: Number of incorrect matches** | Mean (SD): 1.4 (0.6)<br>min < med < max:<br>0 < 1.4 < 3.8<br>IQR (CV): 0.7 (0.4) | 35 distinct values | 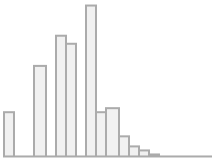 |
| **11** | **Picture vocabulary: Specific cognitive ability** | Mean (SD): 0.4 (0.1)<br>min < med < max:<br>0 < 0.4 < 0.6<br>IQR (CV): 0.1 (0.2) | 3834 distinct values | 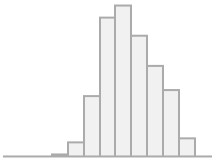 |
| **12** | **Prospective memory: Initial answer** | Min: 0<br>Mean: 0.8<br>Max: 1 | 0: 5502 (17.4%)<br>1: 26112 (82.6%) | 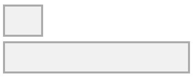 |

*SD* standard deviation, *IQR* interquartile range, *CV* coefficient of variation.

#### Derivation of the *g*-factor with Exploratory Structural Equation (ESEM) Modeling

To examine data factorability, i.e., suitability of the data for factor analysis, we derived the Kaiser-Meyer-Olkin (KMO) statistic for factor adequacy and ran Bartlett's test of sphericity [4, 5]. We then applied exploratory factor analysis (EFA) that compares the eigenvalues of the actual correlation matrix with those from a random matrix of the same size as the original from the random data simulations to determine the number of factors and the loadings of the cognitive test scores onto the latent factors (*fa.parallel* function of *psych* package in R) [6]. We assessed the output of EFA with ESEM (*esem_efa* function of *esemComp* package) and used anchor variables obtained in the previous step to build and fit a hierarchical model. To retain factor loadings, we used confirmatory factor analysis (CFA) with the ML (maximum likelihood) estimator and NLMINB (Non-Linear Minimization using the Broyden–Fletcher–Goldfarb–Shanno algorithm) optimization method. Finally, we evaluated the goodness of model fit using the Comparative Fit Index (CFI), the Tucker-Lewis Index (TLI), the Root Mean Squared Error of Approximation (RMSEA), the Bayesian Information Criteria (BIC), and the Standardized Root Mean Square Residual (SRMR), and obtained *g*-factor values using *lavaan* package [7–11].

**Table S3.** Characteristics of the train and test sets used to build *g*-factor in each of the five folds

| **Fold** | **Number** | | **Age, Mean (SD)** | | **Females, %** | |
|---|---|---|---|---|---|---|
| | **Train** | **Test** | **Train** | **Test** | **Train** | **Test** |
| **1** | 25290 | 6323 | 64.52±7.64 | 64.48 (7.74) | 51.39 | 51.16 |
| **2** | 25291 | 6322 | 64.53±7.66 | 64.44 (7.62) | 51.19 | 51.96 |
| **3** | 25290 | 6323 | 64.53±7.66 | 64.45 (7.65) | 51.38 | 51.19 |
| **4** | 25290 | 6323 | 64.5±7.66 | 64.56 (7.65) | 51.27 | 51.64 |
| **5** | 25291 | 6322 | 64.48±7.67 | 64.63 (7.61) | 51.49 | 50.78 |

*SD* standard deviation.

### S2. Mental Health

#### Description of core measures

Composite mental health scores included the Generalized Anxiety Disorder (GAD-7), the Posttraumatic Stress Disorder (PTSD) Checklist (PCL-6), the Alcohol Use Disorders Identification Test (AUDIT), the Patient Health Questionnaire (PHQ-9) [12], the Eysenck Neuroticism (N-12), Probable Depression Status (PDS), and the Recent Depressive Symptoms (RDS-4) scores [13, 14]. To calculate the GAD-7, PCL-6, AUDIT, and PHQ-9, we used questions introduced at the online follow-up [12]. To obtain the N-12, PDS, and RDS-4 scores [14], we used data collected during the baseline assessment [13, 14].

We subcategorized depression and GAD based on frequency, current status (ever had depression or anxiety and current status of depression or anxiety), severity, and clinical diagnosis (depression or anxiety confirmed by a healthcare practitioner). Additionally, we differentiated between different depression statuses, such as recurrent depression, depression triggered by loss, etc. Variables related to self-harm were subdivided based on whether a person has ever self-harmed with the intent to die.

To make response scales more intuitive, we recorded responses within the well-being domain such that the lower score corresponded to a lesser extent of satisfaction (“Extremely unhappy”) and the higher score indicated a higher level of happiness (“Extremely happy”). For all questions, we assigned the median values to “Prefer not to answer” (-818 for in-person assessment and -3 for online questionnaire) and “Do not know” (-121 for in-person assessment and -1 for online questionnaire) responses. We excluded the “Work/job satisfaction” question from the mental health derivatives list because it included a “Not employed” response option, which could not be reasonably coded.

To calculate the risk of PTSD, we used questions from the PCL-6 questionnaire. Following Davis and colleagues [12], PCL-6 scores ranged from 6 to 29. A PCL-6 score of 12 or below corresponds to a low risk of meeting the Clinician-Administered PTSD Scale diagnostic criteria. PCL-6 scores between 13 and 16 and between 17 and 25 are indicative of an increased risk and high risk of PTSD, respectively. A score of above 26 is interpreted as a very high risk of PTSD [12, 15]. PTSD status was set to positive if the PCL-6 score exceeded or was equal to 14 and encompassed stressful events instead of catastrophic trauma alone [12].

To assess alcohol consumption, alcohol dependence, and harm associated with drinking, we calculated the sum of the ten questions from the AUDIT questionnaire [16]. We additionally subdivided the AUDIT score into the alcohol consumption score (questions 1-3, AUDIT-C) and the score reflecting problems caused by alcohol (questions 4-10, AUDIT-P) [17]. In questions 2-10 that followed the first trigger question ("Frequency of drinking alcohol"), we replaced missing values with 0 as they would correspond to a "Never" response to the first question. An AUDIT score cut-off of 8 suggests moderate or low-risk alcohol consumption, and scores of 8 to 15 and above 15 indicate severe/harmful and hazardous (alcohol dependence or moderate-severe alcohol use disorder) drinking, respectively [16, 18]. Subsequently, hazardous alcohol use and alcohol dependence status correspond to AUDIT scores of ≥ 8 and ≥ 15, respectively. The "Alcohol dependence ever" status was set to positive if a participant had ever been physically dependent on alcohol. To reduce skewness, we $\log_{x+1}$-transformed the AUDIT, AUDIT-C, and AUDIT-P scores [17].

**Table S4.** Derivation of mental health scores

| Disorder / Exposure | Definition | Fields | Resources |
|---|---|---|---|
| **PHQ-9** | The sum of the nine depressive symptoms scored 0 to 4:<br>• Little interest or pleasure in doing things<br>• Feeling down depressed, or hopeless<br>• Trouble sleeping<br>• Feeling tired<br>• Poor appetite or overeating<br>• Feeling bad about yourself<br>• Trouble concentrating<br>• Moving or speaking slowly or fidgety or restless<br>• Thoughts that you would be better off dead<br>Answers "Prefer not to answer" were | 20507<br>20508<br>20510<br>20511<br>20513<br>20514<br>20517<br>20518<br>20519 | **Davis et al., 2020**<br>Kroenke K, Spitzer RL, Williams JB, Löwe B. The patient health questionnaire somatic, anxiety, and depressive symptom scales: a systematic review. Gen Hosp Psychiatry. 2010;32(4):345–59 |

| | | | |
|---|---|---|---|
| | assigned the lowest score (0). | | |
| **Depression ever** | At least one core symptom of depression (Persistent sadness or Loss of interest) that lasted most or all of the day on most or all days within two weeks with some or a lot of impact on normal activity, plus additional depressive symptoms that represent a change in the mental and/or physical state from usual state and occur over the same period with thoughts about death.<br><br>A total of ≥ 5 symptoms, including core symptoms. The score is obtained based on the DSM definition of major depressive disorder. | 20435<br>20436<br>20437<br>20439<br>20440<br>20441<br>20446<br>20449<br>20450<br>20532<br>20536 | **Davis et al., 2020**<br><br>CIDI-SF (Composite International Diagnostic Interview – Short Form), depression module, lifetime version<br><br>Kessler RC, Andrews G, Mroczek D, Ustun B, Wittchen HU. The World Health Organization composite international diagnostic interview short-form (CIDI-SF). Int J Methods Psychiatr Res. 1998;7(4):171–85 |
| **Bipolar affective disorder type I** | Ever had depression and ever manic/hyper or irritable, plus at least three manifestations of mania or irritability (more talkative, more restless, thoughts racing, needed less sleep, more creative or had more ideas, easily distracted, more confident, more active) or four manifestations if never manic/hyper, plus duration of symptoms for a week or more and symptoms caused significant problems. | 20435<br>20436<br>20437<br>20439<br>20440<br>20441<br>20446<br>20449<br>20450<br>20492<br>20493<br>20501<br>20502<br>20532<br>20536<br>20548 | **Davis et al., 2020**<br><br>Cerimele JM, Chwastiak LA, Dodson S, Katon WJ. The prevalence of bipolar disorder in general primary care samples: a systematic review. Gen Hosp Psychiatry. 2014;36(1):19–25<br><br>Carvalho AF, Takwoingi Y, Sales PM, et al. Screening for bipolar spectrum disorders: A comprehensive meta-analysis of accuracy studies. J Affect Disord. 2015;172:337–346 |
| **Bipolar affective disorder type II** | Ever had depression and ever manic/hyper or irritable, plus at least three manifestations of mania or irritability (more talkative, more restless, thoughts racing, needed less sleep, more creative or had more ideas, easily distracted, more confident, more active) or four manifestations if never manic/hyper, plus duration of symptoms for a week or more and symptoms did not cause significant problems. | 20435<br>20436<br>20437<br>20439<br>20440<br>20441<br>20446<br>20449<br>20450 | **Davis et al., 2020** |

| | | | |
|---|---|---|---|
| | | 20492<br>20501<br>20502<br>20532<br>20536<br>20548 | |
| **Subthreshold depressive symptoms ever** | Does not meet Composite International Diagnostic Interview (CIDI) diagnostic criteria for depression, but has at least one of the following symptoms:<br>● Persistent depression or anhedonia based on CIDI<br>● PHQ9 score for current depressive symptoms exceeds the threshold for mild depression<br>● The presence of a clinician diagnosis of depression. | 20002<br>20435<br>20436<br>20437<br>20439<br>20440<br>20441<br>20446<br>20449<br>20450<br>20507<br>20508<br>20510<br>20511<br>20513<br>20514<br>20517<br>20518<br>20519<br>20532<br>20536<br>20544 | **Davis et al., 2020**<br>National Institute for Health and Clinical Excellence. Depression in adults: recognition and management. NICE Clinical Guideline CG90 |
| **Depression single episode** | A single episode of depression without bipolar disorder type I. | 20435<br>20436<br>20437<br>20439<br>20440<br>20441<br>20442<br>20446<br>20449<br>20450 | **Davis et al., 2020** |

| | | | |
|---|---|---|---|
| | | 20492<br>20493<br>20501<br>20502<br>20532<br>20536 | |
| **Recurrent depression** | More than one episode of depression throughout a lifetime without bipolar disorder type I. | 20435<br>20436<br>20437<br>20439<br>20440<br>20441<br>20442<br>20446<br>20449<br>20450<br>20492<br>20493<br>20501<br>20502<br>20532<br>20536 | **Davis et al., 2020** |
| **Depression single episode triggered by a loss** | A single episode of depression that started within two months after a traumatic event. | 20435<br>20436<br>20437<br>20439<br>20440<br>20441<br>20442<br>20446<br>20447<br>20449<br>20450<br>20492<br>20493<br>20501<br>20502<br>20532 | **Davis et al., 2020** |

| | | | |
|---|---|---|---|
| | | 20536 | |
| **Current depression** | At least a single episode of depression ("Depression ever") with a minimum of 5 depression symptoms from the PHQ-9 occurring more than half days or for several days for suicidal thoughts. | 20435<br>20436<br>20437<br>20439<br>20440<br>20441<br>20446<br>20449<br>20450<br>20507<br>20508<br>20510<br>20511<br>20513<br>20514<br>20517<br>20518<br>20519<br>20532<br>20536 | **Davis et al., 2020**<br>Manea L, Gilbody S, McMillan D. Optimal cut-off score for diagnosing depression with the Patient Health Questionnaire (PHQ-9): a meta-analysis. CMAJ. 2012;184(3):E191–E6 |
| **Current severe depression** | At least a single episode of depression ("Depression ever") As current depression (above) with PHQ score > 15. | 20435<br>20436<br>20437<br>20439<br>20440<br>20441<br>20446<br>20449<br>20450<br>20507<br>20508<br>20510<br>20511<br>20513<br>20514<br>20517<br>20518 | **Davis et al., 2020**<br>Manea L, Gilbody S, McMillan D. Optimal cut-off score for diagnosing depression with the Patient Health Questionnaire (PHQ-9): a meta-analysis. CMAJ. 2012;184(3):E191–E6 |

| | | | |
|---|---|---|---|
| | | 20519<br>20532<br>20536 | |
| **GAD-7** | The sum of the recent symptoms of anxiety scored 0 to 3:<br>● Feelings of nervousness or anxiety<br>● Inability to stop or control worrying<br>● Worrying too much about different things<br>● Trouble relaxing<br>● Restlessness<br>● Easy annoyance or irritability<br>● Feelings of foreboding. | 20505<br>20506<br>20509<br>20512<br>20515<br>20516<br>20520 | **Davis et al., 2020**<br>Kroenke K, Spitzer RL, Williams JB, Löwe B. The patient health questionnaire somatic, anxiety, and depressive symptom scales: a systematic review. Gen Hosp Psychiatry. 2010;32(4):345–59 |
| **Lifetime anxiety disorder (GAD ever)** | Ever felt worried, tense, or anxious for most of the day for at least six months with difficulties in controlling symptoms. The symptoms were often difficult to control, they interfered with daily activity and were accompanied by at least three somatic symptoms (restless, keyed up or on edge., easily tired, difficulty keeping the mind on current activity, more irritable than usual, tense muscles, trouble falling or staying asleep). | 20417<br>20418<br>20419<br>20420<br>20421<br>20422<br>20423<br>20425<br>20426<br>20427<br>20429<br>20537<br>20538<br>20539<br>20540<br>20541<br>20542<br>20543 | **Davis et al., 2020**<br>CIDI-SF (Composite International Diagnostic Interview – Short Form), GAD module, lifetime version. Scored based on the DSM definition of GAD<br>Gigantesco A, Morosini P. Development, reliability and factor analysis of a self-administered questionnaire which originates from the World Health Organization's Composite International Diagnostic Interview - Short Form (CIDI-SF) for assessing mental disorders. Clin Pract Epidemiol Ment Health. 2008;4:8<br>Kessler RC, Andrews G, Mroczek D, Ustun B, Wittchen HU. The World Health Organization composite international diagnostic interview short-form (CIDI-SF). Int J Methods Psychiatr Res. 1998;7(4):171–85<br>National Institute for Health and Clinical Excellence. Generalised anxiety disorder |

| | | | |
|---|---|---|---|
| | | | and panic disorder in adults: management. NICE Clinical Guideline CG113 |
| **Current anxiety** | Ever had GAD ("GAD Ever") and GAD-7 score ≥ 10. Subdivided into mild, moderate, and severe with cut-offs at 5, 10, and 15. | 20417<br>20418<br>20419<br>20420<br>20421<br>20422<br>20423<br>20425<br>20426<br>20427<br>20429<br>20505<br>20506<br>20509<br>20512<br>20515<br>20516<br>20520<br>20537<br>20538<br>20539<br>20540<br>20541<br>20542<br>20543 | **Davis et al., 2020**<br>Kroenke K, Spitzer RL, Williams JB, Löwe B. The patient health questionnaire somatic, anxiety, and depressive symptom scales: a systematic review. Gen Hosp Psychiatry. 2010;32(4):345–59 |
| **N-12** | The sum of the following scores:<br>● Mood swings<br>● Miserableness<br>● Irritability<br>● Sensitivity / hurt feelings<br>● Fed-up feelings<br>● Nervous feeling<br>● Worrier / anxious feelings<br>● Tense / "highly strung" | 1920<br>1930<br>1940<br>1950<br>1960<br>1970<br>1980<br>1990<br>2000 | **Dutt et al., 2021**<br>**Smith et al., 2013** |

| | | | |
|---|---|---|---|
| | • Worry too long after embarrassment<br>• Suffer from “nerves”<br>• Loneliness, isolation<br>• Guilty feelings | 2010<br>2020<br>2030 | |
| **PDS** | Ever been depressed or unenthusiastic for at least one week and seen either a GP or psychiatrist for nerves, anxiety, tension, or depression. | 2090<br>2100<br>4598<br>4609<br>4631<br>5375 | **Dutt et al., 2021** |
| **RDS-4** | Frequency of depressed mood, disinterest, restlessness, and tiredness during the past two weeks scored 1 to 4. | 2050<br>2060<br>2070<br>2080 | **Dutt et al., 2021** |
| **PCL-6** | The sum of scores on the core symptoms of PTSD in the past month:<br>• Repeated disturbing thoughts of a stressful experience<br>• Felt very upset when reminded of a stressful experience<br>• Avoided activities or situations because of a previous stressful experience<br>• Felt distant from other people<br>• Felt irritable or had angry outbursts in the past month<br>• Recent trouble concentrating on things<br>The symptoms are grouped into three clusters:<br>• Memories, thoughts, or images, upset when reminded<br>• Avoid activities or situations, feeling distance | 20494<br>20495<br>20496<br>20497<br>20498<br>20508 | **Davis et al., 2020**<br>Lang AJ, Stein MB. An abbreviated PTSD checklist for use as a screening instrument in primary care. Behaviour research and therapy. 2005;43(5):585–94 |

| | | | |
|---|---|---|---|
| | or cut-off<br>• Irritable or angry, difficulty concentrating. | | |
| **PTSD** | PCL-6 score ≥ 14. | 20494<br>20495<br>20496<br>20497<br>20498<br>20508 | **Davis et al., 2020** |
| **Unusual experience** | Experience of hallucinations or delusions, such as:<br>• Unreal voice<br>• Unreal vision<br>• Believed in an unreal conspiracy against self<br>• Believed in unreal communications or signs. | 20463<br>20468<br>20471<br>20474 | **Davis et al., 2020**<br>Nuevo R, Chatterji S, Verdes E, Naidoo N, Arango C, Ayuso-Mateos JL. The Continuum of Psychotic Symptoms in the General Population: A Cross-national Study. Schizophrenia Bulletin. 2012;38(3):475–85 |
| **Recent unusual experience** | Reports at least one or two hallucination or delusion episodes within the last year. | 20467 | **Davis et al., 2020** |
| **Life not worth living** | Ever felt that life was not worth living. | 20479 | **Davis et al., 2020** |
| **Self-harm** | Ever harmed self, whether or not meant to die. | 20480 | **Davis et al., 2020** |
| **Non-suicidal self-harm** | Ever self-harmed without intention to end life, i.e., never attempted suicide. | 20480<br>20483 | **Davis et al., 2020** |
| **Suicide attempt** | Ever harmed self with intent to end life. | 20480<br>20483 | **Davis et al., 2020** |
| **AUDIT** | The sum of scores (0 to 4) on questions about alcohol consumption comprising three domains:<br>• **Consumption**: Frequency, amount of typical drinks, frequency of having six or more | 20403<br>20405<br>20407<br>20408<br>20409 | **Davis et al., 2020**<br>Saunders JB, Aasland OG, Babor TF, de la Fuente JR, Grant M. Development of the Alcohol Use Disorders Identification Test (AUDIT): WHO Collaborative Project on Early Detection of |

| | | | |
|---|---|---|---|
| | drinks<br>● **Dependence**: Unable to stop, failed to do what expected due to drinking, needed to drink first thing<br>● **Harm**: Guilt due to drinking, unable to remember due to drink<br>Plus had injuries due to drinking or advice to cut down on drinking. | 20411<br>20412<br>20413<br>20414<br>20416 | Persons with Harmful Alcohol Consumption–II. Addiction. 1993;88(6):791–804<br>Reinert DF, Allen JP. The alcohol use disorders identification test: an update of research findings. Alcohol Clin Exp Res. 2007;31(2):185–199 |
| **Alcohol consumption (AUDIT-C)** | Sum of questions 1-3 of the Alcohol Consumption domain. | 20403<br>20414<br>20416 | **Sanchez-Roige et al., 2019** |
| **Problems caused by alcohol (AUDIT-P)** | Sum of questions 4-10 of the Alcohol Dependence and Alcohol Harm domains. | 20405<br>20407<br>20408<br>20409<br>20411<br>20412<br>20413 | **Sanchez-Roige et al., 2019** |
| **Hazardous / Harmful alcohol use** | AUDIT score ≥ 8. | 20403<br>20405<br>20407<br>20408<br>20409<br>20411<br>20412<br>20413<br>20414<br>20416 | **Davis et al., 2020**<br>Babor TF, Higgins-Biddle JC, Saunders JB, Monteiro MG. AUDIT: The alcohol use disorders identification test: Guidelines for use in primary health care. 2nd ed. World Health Organization. 2001<br>Stansfeld S, Clark C, Bebbington P, King M, Jenkins R, Hinchliffe S. Chapter 2: Common mental disorders. In McManus S, Bebbington P, Jenkins R, Brugha T. (eds) (2016) Mental health and wellbeing in England: Adult Psychiatric. Morbidity Survey 2014. Leeds: NHS Digital |

| | | | |
|---|---|---|---|
| **Current alcohol dependence** | AUDIT score ≥15. | 20403<br>20405<br>20407<br>20408<br>20409<br>20411<br>20412<br>20413<br>20414<br>20416 | **Davis et al., 2020**<br>Babor TF, Higgins-Biddle JC, Saunders JB, Monteiro MG. AUDIT: The alcohol use disorders identification test: Guidelines for use in primary health care. 2nd ed. World Health Organization. 2001<br>Drummond C, McBride O, Fear N, Fuller E. (2016) 'Chapter 10: Alcohol dependence' in McManus S, Bebbington P, Jenkins R, Brugha T. (eds) Mental health and wellbeing in England: Adult Psychiatric Morbidity Survey 2014. Leeds: NHS Digital |
| **Alcohol dependence ever** | Ever physically dependent on alcohol. | 20404 | **Davis et al., 2020** |
| **Addiction ever** | Ever addicted to any substance or behaviour. | 20401 | **Davis et al., 2020** |
| **Substance addiction** | Ever been addicted to alcohol, illicit/recreational drugs, or medication. | 20406<br>20456<br>20503 | **Davis et al., 2020** |
| **Current addiction** | Ongoing addiction or dependence. | 20415<br>20432<br>20457<br>20504 | **Davis et al., 2020** |
| **Cannabis ever** | Taking cannabis at least once in life. | 20453 | **Davis et al., 2020** |
| **Cannabis daily** | Maximum frequency of taking cannabis when using it every day. | 20454 | **Davis et al., 2020** |
| **Childhood adverse events** | A positive score if any of the five questions of the Childhood Trauma Screen (CTS) reach the threshold:<br>• Felt loved as a child ≤ 3 | 20487<br>20488<br>20489<br>20490 | **Davis et al., 2020**<br>Walker EA, Gelfand A, Katon WJ, et al. Adult health status of women with histories of childhood abuse |

| | | | |
|---|---|---|---|
| | (never, rarely, or sometimes)<br>• Physically abused by family as a child ≥ 2 (often or very often)<br>• Felt hated by a family member as a child ≥ 2 (often or very often)<br>• Sexually molested as a child ≥ 2 (often or very often)<br>• Someone to take to the doctor when needed as a child ≤ 4 (never, rarely, sometimes, or often). | 20491 | and neglect. Am J Med. 1999;107(4):332–339 |
| **Adult adverse events** | A positive score if any of the five questions of the Adult Trauma Screen reach the threshold:<br>• Been in a confiding relationship as an adult ≤ 3 (never, rarely, or sometimes)<br>• Physical violence by partner or ex-partner as an adult ≥ 2 (often or very often)<br>• Belittlement by partner or ex-partner as an adult ≥ 2 (often or very often)<br>• Sexual interference by partner or ex-partner without consent as an adult ≥ 2 (often or very often)<br>• Able to pay rent/mortgage ≤ 4 (never, rarely, sometimes, or often). | 20521<br>20522<br>20523<br>20524<br>20525 | **Davis et al., 2020** |
| **Catastrophic trauma** | At least one catastrophic event:<br>• Victim of sexual assault<br>• Victim of physically violent crime<br>• Been in a serious accident believed to be life-threatening<br>• Witnessed sudden violent death<br>• Diagnosed with a life-threatening illness | 20526<br>20527<br>20528<br>20529<br>20530<br>20531 | **Davis et al., 2020** |

| | | | |
|---|---|---|---|
| | • Been involved in combat or exposed to war zones. | | |
| **Wellbeing** | The sum of the following scores:<br>• General happiness<br>• Happiness with own health<br>• Belief that life is meaningful. | 20458<br>20459<br>20460 | **Davis et al., 2020** |
| **Any distress** | Reported functional impairment due to mental distress:<br>• Ever sought/received help for mental distress<br>• Mental distress prevented usual activities<br>• Mental health problems diagnosed by a healthcare professional<br>plus, a positive diagnosis for a specific condition (Depression ever, GAD ever, Addiction ever, Bipolar ever, Psychotic experiences, PTSD, Self-harm ever). | 20499<br>20500<br>20544 | **Davis et al., 2020** |

*N-12* the Eysenck Neuroticism score, *PTSD* posttraumatic stress disorder, *PCL-6* PTSD Checklist, *PHQ-9* Patient Health Questionnaire score, *PDS* Probable Depression Status, *RDS-4* Recent Depressive Symptoms, *AUDIT* Alcohol Use Disorders Identification Test, *GAD-7* Generalized Anxiety Disorder score, *GP* general practitioner.

**Table S5.** Whole-sample distributions of mental health measures used as features in machine learning models (N = 21 077)

| No | Variable | Mental Health Domain | Statistics / Values | Frequencies | Distribution Plot |
|---|---|---|---|---|---|
| 1 | **Diagnoses: Anxiety/panic attacks** | Clinical diagnoses (Non-cancer illness code, self-reported) | Min: 0<br>Mean: 0<br>Max: 1 | 0: 20324 (96.4%)<br>1: 753 (3.6%) | 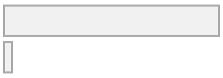 |
| 2 | **N-12** | Neuroticism | Mean (SD): 3.3 (3)<br>min < med < max:<br>0 < 3 < 12<br>IQR (CV): 4 (0.9) | 13 distinct values | 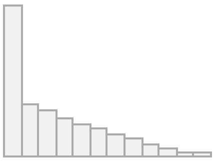 |
| 3 | **PCL-6** | Traumatic events | Mean (SD): 7.8 (2.9)<br>min < med < max:<br>6 < 6 < 29<br>IQR (CV): 3 (0.4) | 24 distinct values | 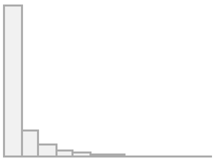 |

| | | | | | |
|---|---|---|---|---|---|
| 4 | **PHQ-9** | Depression | Mean (SD): 2.6 (3.5)<br>min < med < max:<br>0 < 1 < 27<br>IQR (CV): 4 (1.4) | 28 distinct values | 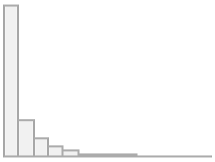 |
| 5 | **PDS** | Depression | Min: 0<br>Mean: 0.2<br>Max: 1 | 0: 16367 (77.7%)<br>1: 4710 (22.3%) | 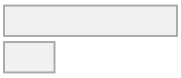 |
| 6 | **RDS-4** | Depression | Mean (SD): 5.1 (1.7)<br>min < med < max:<br>4 < 5 < 16<br>IQR (CV): 2 (0.3) | 13 distinct values | 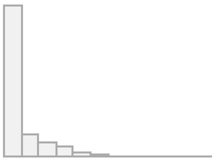 |
| 7 | **Diagnoses: Depression** | Clinical diagnoses (Non-cancer illness code, self-reported) | Min: 0<br>Mean: 0.1<br>Max: 1 | 0: 19144 (90.8%)<br>1: 1933 (9.2%) | 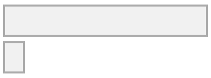 |
| 8 | **Self-harm: Ever thought life not worth living** | Self-harm behaviours | Min: 0<br>Mean: 0.3<br>Max: 1 | 0: 14661 (69.6%)<br>1: 6416 (30.4%) | 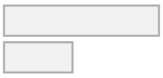 |
| 9 | **Ever attempted suicide** | Self-harm behaviours | Min: 0<br>Mean: 0<br>Max: 1 | 0: 20675 (98.1%)<br>1: 402 (1.9%) | 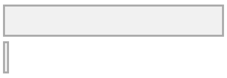 |
| 10 | **Ever self-harmed** | Self-harm behaviours | Min: 0<br>Mean: 0<br>Max: 1 | 0: 20238 (96.0%)<br>1: 839 (4.0%) | 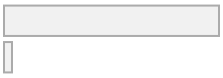 |
| 11 | **Ever self-harmed (non-suicidal)** | Self-harm behaviours | Min: 0<br>Mean: 0<br>Max: 1 | 0: 20665 (98.0%)<br>1: 412 (2.0%) | 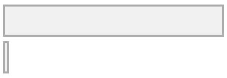 |
| 12 | **Unusual experience** | Unusual/psychotic experiences | Min: 0<br>Mean: 0<br>Max: 1 | 0: 20083 (95.3%)<br>1: 994 (4.7%) | 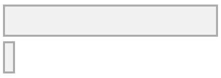 |
| 13 | **Recent unusual experience** | Unusual/psychotic experiences | Min: 0<br>Mean: 0<br>Max: 1 | 0: 20820 (98.8%)<br>1: 257 (1.2%) | 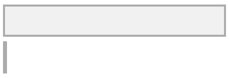 |
| 14 | **Repeated disturbing thoughts of stressful experience in past month** | Traumatic events | Mean (SD): 0.4 (0.7)<br>min < med < max:<br>0 < 0 < 4<br>IQR (CV): 1 (2) | 0: 15611 (74.1%)<br>1: 4151 (19.7%)<br>2: 691 (3.3%)<br>3: 520 (2.5%)<br>4: 104 (0.5%) | 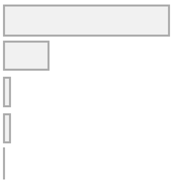 |
| 15 | **Felt very upset when reminded of stressful experience in past month** | Traumatic events | Mean (SD): 0.5 (0.8)<br>min < med < max:<br>0 < 0 < 4<br>IQR (CV): 1 (1.6) | 0: 13692 (65.0%)<br>1: 5718 (27.1%)<br>2: 941 (4.5%)<br>3: 559 (2.7%)<br>4: 167 (0.8%) | 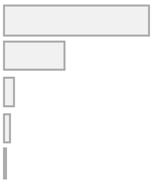 |
| 16 | **Avoided activities or situations because of previous stressful experience in past month** | Traumatic events | Mean (SD): 0.3 (0.6)<br>min < med < max:<br>0 < 0 < 4<br>IQR (CV): 0 (2.4) | 0: 17051 (80.9%)<br>1: 2981 (14.1%)<br>2: 547 (2.6%)<br>3: 401 (1.9%)<br>4: 97 (0.5%) | 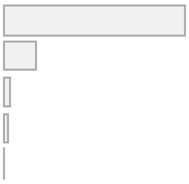 |
| 17 | **Frequency of "life not worth living" thoughts** | Self-harm behaviours | Mean (SD): 0.5 (0.8)<br>min < med < max:<br>0 < 0 < 2<br>IQR (CV): 1 (1.6) | 0: 14661 (69.6%)<br>1: 2812 (13.3%)<br>2: 3604 (17.1%) | 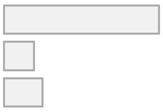 |
| 18 | **Lifetime frequency of contemplating self-harm** | Self-harm behaviours | Mean (SD): 0.2 (0.6)<br>min < med < max:<br>0 < 0 < 2<br>IQR (CV): 0 (2.6) | 0: 17956 (85.2%)<br>1: 1573 (7.5%)<br>2: 1548 (7.3%) | 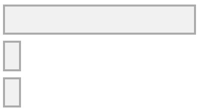 |

| | | | | |
|---|---|---|---|---|
| **19** | **Ever had prolonged feelings of sadness or depression** | Depression | Min: 0<br>Mean: 0.5<br>Max: 1 | 0: 9708 (46.1%)<br>1: 11369 (53.9%) |
| **20** | **Ever had prolonged loss of interest in normal activities** | Depression | Min: 0<br>Mean: 0.4<br>Max: 1 | 0: 13043 (61.9%)<br>1: 8034 (38.1%) |
| **21** | **Ever felt worried, tense, or anxious for most of a month or longer** | Anxiety | Min: 0<br>Mean: 0.2<br>Max: 1 | 0: 15908 (75.5%)<br>1: 5169 (24.5%) |
| **22** | **Ever worried more than most people would in a similar situation** | Anxiety | Min: 0<br>Mean: 0.2<br>Max: 1 | 0: 16689 (79.2%)<br>1: 4388 (20.8%) |
| **23** | **Ever addicted to any substance or behaviour** | Addictions | Min: 0<br>Mean: 0.1<br>Max: 1 | 0: 19896 (94.4%)<br>1: 1181 (5.6%) |
| **24** | **Ever believed in an unreal conspiracy against self** | Unusual/psychotic experiences | Min: 0<br>Mean: 0<br>Max: 1 | 0: 20937 (99.3%)<br>1: 140 (0.7%) |
| **25** | **Ever believed in unreal communications or signs** | Unusual/psychotic experiences | Min: 0<br>Mean: 0<br>Max: 1 | 0: 20944 (99.4%)<br>1: 133 (0.6%) |
| **26** | **Ever heard an unreal voice** | Unusual/psychotic experiences | Min: 0<br>Mean: 0<br>Max: 1 | 0: 20727 (98.3%)<br>1: 350 (1.7%) |
| **27** | **Ever seen an unreal vision** | Unusual/psychotic experiences | Min: 0<br>Mean: 0<br>Max: 1 | 0: 20432 (96.9%)<br>1: 645 (3.1%) |
| **28** | **Mood swings** | Neuroticism | Min: 0<br>Mean: 0.3<br>Max: 1 | 0: 14186 (67.3%)<br>1: 6891 (32.7%) |
| **29** | **Miserableness** | Neuroticism | Min: 0<br>Mean: 0.3<br>Max: 1 | 0: 14338 (68.0%)<br>1: 6739 (32.0%) |
| **30** | **Irritability** | Neuroticism | Min: 0<br>Mean: 0.2<br>Max: 1 | 0: 16056 (76.2%)<br>1: 5021 (23.8%) |
| **31** | **Sensitivity / hurt feelings** | Neuroticism | Min: 0<br>Mean: 0.4<br>Max: 1 | 0: 12287 (58.3%)<br>1: 8790 (41.7%) |
| **32** | **Fed-up feelings** | Neuroticism | Min: 0<br>Mean: 0.3<br>Max: 1 | 0: 15147 (71.9%)<br>1: 5930 (28.1%) |
| **33** | **Nervous feelings** | Neuroticism | Min: 0<br>Mean: 0.2<br>Max: 1 | 0: 17631 (83.7%)<br>1: 3446 (16.3%) |
| **34** | **Worrier / anxious feelings** | Neuroticism | Min: 0<br>Mean: 0.5<br>Max: 1 | 0: 11178 (53.0%)<br>1: 9899 (47.0%) |
| **35** | **Tense / “highly strung”** | Neuroticism | Min: 0<br>Mean: 0.1<br>Max: 1 | 0: 18864 (89.5%)<br>1: 2213 (10.5%) |
| **36** | **Worry too long after embarrassment** | Neuroticism | Min: 0<br>Mean: 0.4<br>Max: 1 | 0: 11600 (55.0%)<br>1: 9477 (45.0%) |
| **37** | **Suffer from “nerves”** | Neuroticism | Min: 0<br>Mean: 0.1<br>Max: 1 | 0: 18348 (87.1%)<br>1: 2729 (12.9%) |
| **38** | **Loneliness, isolation** | Neuroticism | Min: 0<br>Mean: 0.1<br>Max: 1 | 0: 18362 (87.1%)<br>1: 2715 (12.9%) |

| | | | | |
|---|---|---|---|---|
| 39 | **Guilty feelings** | Neuroticism | Min: 0<br>Mean: 0.2<br>Max: 1 | 0: 15978 (75.8%)<br>1: 5099 (24.2%) |
| 40 | **Risk taking** | Neuroticism | Min: 0<br>Mean: 0.2<br>Max: 1 | 0: 15911 (75.5%)<br>1: 5166 (24.5%) |
| 41 | **Frequency of depressed mood in last 2 weeks** | Depression | Mean (SD): 1.2 (0.5)<br>min < med < max:<br>1 < 1 < 4<br>IQR (CV): 0 (0.4) | 1: 17559 (83.3%)<br>2: 3020 (14.3%)<br>3: 308 (1.5%)<br>4: 190 (0.9%) |
| 42 | **Frequency of unenthusiasm/disinterest in the last 2 weeks** | Depression | Mean (SD): 1.2 (0.5)<br>min < med < max:<br>1 < 1 < 4<br>IQR (CV): 0 (0.4) | 1: 18031 (85.5%)<br>2: 2553 (12.1%)<br>3: 308 (1.5%)<br>4: 185 (0.9%) |
| 43 | **Frequency of tenseness/restlessness in last 2 weeks** | Depression | Mean (SD): 1.2 (0.5)<br>min < med < max:<br>1 < 1 < 4<br>IQR (CV): 0 (0.4) | 1: 17150 (81.4%)<br>2: 3504 (16.6%)<br>3: 266 (1.3%)<br>4: 157 (0.7%) |
| 44 | **Frequency of tiredness/lethargy in last 2 weeks** | Depression | Mean (SD): 1.5 (0.7)<br>min < med < max:<br>1 < 1 < 4<br>IQR (CV): 1 (0.5) | 1: 11831 (56.1%)<br>2: 7698 (36.5%)<br>3: 824 (3.9%)<br>4: 724 (3.4%) |
| 45 | **Seen a psychiatrist for nerves, anxiety, tension, or depression** | Depression | Min: 0<br>Mean: 0.1<br>Max: 1 | 0: 19200 (91.1%)<br>1: 1877 (8.9%) |
| 46 | **Seen a doctor (GP) for nerves, anxiety, tension, or depression** | Depression | Min: 0<br>Mean: 0.3<br>Max: 1 | 0: 14774 (70.1%)<br>1: 6303 (29.9%) |
| 47 | **Ever depressed for a whole week** | Depression | Mean (SD): 0.6 (0.6)<br>min < med < max:<br>0 < 1 < 3<br>IQR (CV): 1 (1.1) | 0: 9985 (47.4%)<br>1: 10727 (50.9%)<br>3: 365 (1.7%) |
| 48 | **Ever unenthusiastic/ disinterested for a whole week** | Depression | Min: 0<br>Mean: 0.3<br>Max: 1 | 0: 14705 (69.8%)<br>1: 6372 (30.2%) |
| 49 | **Ever highly irritable/ argumentative for 2 days** | Mania | Min: 0<br>Mean: 0.1<br>Max: 1 | 0: 18868 (89.5%)<br>1: 2209 (10.5%) |
| 50 | **Ever manic/hyper for 2 days** | Mania | Min: 0<br>Mean: 0<br>Max: 1 | 0: 20508 (97.3%)<br>1: 569 (2.7%) |
| 51 | **Mental and behavioural disorders** | Clinical diagnoses (ICD-10 code: F) | Mean (sd): 0 (0.1)<br>min < med < max:<br>0 < 0 < 3<br>IQR (CV): 0 (12.7) | 0: 20934 (99.3%)<br>1: 129 (0.6%)<br>2: 12 (0.1%)<br>3: 2 (0.0%) |
| 52 | **Diseases of the nervous system** | Clinical diagnoses (ICD-10 code: G) | Mean (sd): 0.1 (0.2)<br>min < med < max:<br>0 < 0 < 2<br>IQR (CV): 0 (4.2) | 0: 19879 (94.3%)<br>1: 1144 (5.4%)<br>2: 54 (0.3%) |
| 53 | **Diagnoses: Neurological problems, nervous system injury, epilepsy** | Clinical diagnoses (Non-cancer illness code, self-reported) | Min: 0<br>Mean: 0<br>Max: 1 | 0: 20659 (98.0%)<br>1: 418 (2.0%) |
| 54 | **Diagnoses: Stress, insomnia, migraine, nervous/mental problems** | Clinical diagnoses (Non-cancer illness code, self-reported) | Min: 0<br>Mean: 0.1<br>Max: 1 | 0: 19349 (91.8%)<br>1: 1728 (8.2%) |

| | | | | |
|---|---|---|---|---|
| 55 | **Ever had a period of mania/excitability** | Mania | Min: 0<br>Mean: 0<br>Max: 1 | 0: 20232 (96.0%)<br>1: 845 (4.0%) |
| 56 | **Ever had a period of extreme irritability** | Mania | Min: 0<br>Mean: 0.3<br>Max: 1 | 0: 15705 (74.5%)<br>1: 5372 (25.5%) |
| 57 | **Recent feelings of inadequacy** | Depression | Mean (SD): 1.2 (0.6)<br>min < med < max:<br>1 < 1 < 4<br>IQR (CV): 0 (0.5) | 1: 17073 (81.0%)<br>2: 3245 (15.4%)<br>3: 393 (1.9%)<br>4: 366 (1.7%) |
| 58 | **Recent trouble concentrating on things** | Depression | Mean (SD): 1.2 (0.5)<br>min < med < max:<br>1 < 1 < 4<br>IQR (CV): 0 (0.4) | 1: 17405 (82.6%)<br>2: 3021 (14.3%)<br>3: 378 (1.8%)<br>4: 273 (1.3%) |
| 59 | **Recent feelings of depression** | Depression | Mean (SD): 1.2 (0.5)<br>min < med < max:<br>1 < 1 < 4<br>IQR (CV): 0 (0.4) | 1: 16787 (79.6%)<br>2: 3675 (17.4%)<br>3: 400 (1.9%)<br>4: 215 (1.0%) |
| 60 | **Recent poor appetite or overeating** | Depression | Mean (SD): 1.2 (0.6)<br>min < med < max:<br>1 < 1 < 4<br>IQR (CV): 0 (0.5) | 1: 17549 (83.3%)<br>2: 2582 (12.3%)<br>3: 491 (2.3%)<br>4: 455 (2.2%) |
| 61 | **Recent thoughts of suicide or self-harm** | Depression | Mean (SD): 1 (0.3)<br>min < med < max:<br>1 < 1 < 4<br>IQR (CV): 0 (0.2) | 1: 20282 (96.2%)<br>2: 681 (3.2%)<br>3: 64 (0.3%)<br>4: 50 (0.2%) |
| 62 | **Recent lack of interest or pleasure in doing things** | Depression | Mean (SD): 1.2 (0.5)<br>min < med < max:<br>1 < 1 < 4<br>IQR (CV): 0 (0.4) | 1: 17456 (82.8%)<br>2: 2928 (13.9%)<br>3: 429 (2.0%)<br>4: 264 (1.3%) |
| 63 | **Trouble falling or staying asleep, or sleeping too much** | Depression | Mean (SD): 1.7 (0.9)<br>min < med < max:<br>1 < 1 < 4<br>IQR (CV): 1 (0.5) | 1: 11002 (52.2%)<br>2: 7210 (34.2%)<br>3: 1366 (6.5%)<br>4: 1499 (7.1%) |
| 64 | **Recent changes in speed/amount of moving or speaking** | Depression | Mean (SD): 1.1 (0.3)<br>min < med < max:<br>1 < 1 < 4<br>IQR (CV): 0 (0.3) | 1: 20085 (95.3%)<br>2: 798 (3.8%)<br>3: 128 (0.6%)<br>4: 66 (0.3%) |
| 65 | **Recent feelings of tiredness or low energy** | Depression | Mean (SD): 1.6 (0.8)<br>min < med < max:<br>1 < 1 < 4<br>IQR (CV): 1 (0.5) | 1: 11169 (53.0%)<br>2: 7964 (37.8%)<br>3: 1029 (4.9%)<br>4: 915 (4.3%) |
| 66 | **Depression ever** | Depression | Min: 0<br>Mean: 0.2<br>Max: 1 | 0: 16248 (77.1%)<br>1: 4829 (22.9%) |
| 67 | **Subthreshold depression** | Depression | Min: 0<br>Mean: 0.4<br>Max: 1 | 0: 13473 (63.9%)<br>1: 7604 (36.1%) |
| 68 | **Bipolar I** | Mania | Min: 0<br>Mean: 0<br>Max: 1 | 0: 20979 (99.5%)<br>1: 98 (0.5%) |

| | | | | |
|---|---|---|---|---|
| 69 | **Bipolar II** | Mania | Min: 0<br>Mean: 0<br>Max: 1 | 0: 20903 (99.2%)<br>1: 174 (0.8%) |
| 70 | **Depression single episode** | Depression | Min: 0<br>Mean: 0.1<br>Max: 1 | 0: 19221 (91.2%)<br>1: 1856 (8.8%) |
| 71 | **Recurrent depression** | Depression | Min: 0<br>Mean: 0.1<br>Max: 1 | 0: 18334 (87.0%)<br>1: 2743 (13.0%) |
| 72 | **Depression triggered by loss** | Depression | Min: 0<br>Mean: 0.1<br>Max: 1 | 0: 19622 (93.1%)<br>1: 1455 (6.9%) |
| 73 | **Current depression** | Depression | Min: 0<br>Mean: 0<br>Max: 1 | 0: 20764 (98.5%)<br>1: 313 (1.5%) |
| 74 | **Current severe depression** | Depression | Min: 0<br>Mean: 0<br>Max: 1 | 0: 20873 (99.0%)<br>1: 204 (1.0%) |
| 75 | **Recent easy annoyance or irritability** | Anxiety | Mean (SD): 1.3 (0.6)<br>min < med < max:<br>1 < 1 < 4<br>IQR (CV): 1 (0.4) | 1: 15570 (73.9%)<br>2: 4879 (23.1%)<br>3: 369 (1.8%)<br>4: 259 (1.2%) |
| 76 | **Recent feelings of nervousness or anxiety** | Anxiety | Mean (SD): 1.3 (0.6)<br>min < med < max:<br>1 < 1 < 4<br>IQR (CV): 1 (0.5) | 1: 15515 (73.6%)<br>2: 4745 (22.5%)<br>3: 436 (2.1%)<br>4: 381 (1.8%) |
| 77 | **Recent inability to stop or control worrying** | Anxiety | Mean (SD): 1.3 (0.6)<br>min < med < max:<br>1 < 1 < 4<br>IQR (CV): 0 (0.5) | 1: 16568 (78.6%)<br>2: 3694 (17.5%)<br>3: 407 (1.9%)<br>4: 408 (1.9%) |
| 78 | **Recent feelings of foreboding** | Anxiety | Mean (SD): 1.2 (0.5)<br>min < med < max:<br>1 < 1 < 4<br>IQR (CV): 0 (0.4) | 1: 17842 (84.7%)<br>2: 2616 (12.4%)<br>3: 330 (1.6%)<br>4: 289 (1.4%) |
| 79 | **Recent trouble relaxing** | Anxiety | Mean (SD): 1.3 (0.7)<br>min < med < max:<br>1 < 1 < 4<br>IQR (CV): 1 (0.5) | 1: 15371 (72.9%)<br>2: 4620 (21.9%)<br>3: 529 (2.5%)<br>4: 557 (2.6%) |
| 80 | **Recent restlessness** | Anxiety | Mean (SD): 1.1 (0.4)<br>min < med < max:<br>1 < 1 < 4<br>IQR (CV): 0 (0.4) | 1: 18739 (88.9%)<br>2: 1898 (9.0%)<br>3: 266 (1.3%)<br>4: 174 (0.8%) |
| 81 | **Recent worrying too much about different things** | Anxiety | Mean (SD): 1.4 (0.6)<br>min < med < max:<br>1 < 1 < 4<br>IQR (CV): 1 (0.5) | 1: 14840 (70.4%)<br>2: 5259 (25.0%)<br>3: 499 (2.4%)<br>4: 479 (2.3%) |
| 82 | **GAD-7** | Anxiety | Mean (SD): 1.9 (3.2)<br>min < med < max:<br>0 < 0 < 21<br>IQR (CV): 3 (1.6) | 22 distinct values |
| 83 | **GAD ever** | Anxiety | Min: 0<br>Mean: 0.1<br>Max: 1 | 0: 19690 (93.4%)<br>1: 1387 (6.6%) |

| | | | | |
|---|---|---|---|---|
| **84** | **Current GAD** | Anxiety | Min: 0<br>Mean: 0<br>Max: 1 | 0: 20779 (98.6%)<br>1: 298 (1.4%) |
| **85** | **Current GAD mild** | Anxiety | Min: 0<br>Mean: 0<br>Max: 1 | 0: 20667 (98.1%)<br>1: 410 (1.9%) |
| **86** | **Current GAD moderate** | Anxiety | Min: 0<br>Mean: 0<br>Max: 1 | 0: 20909 (99.2%)<br>1: 168 (0.8%) |
| **87** | **Current GAD severe** | Anxiety | Min: 0<br>Mean: 0<br>Max: 1 | 0: 20947 (99.4%)<br>1: 130 (0.6%) |
| **88** | **PTSD** | Traumatic events | Min: 0<br>Mean: 0.1<br>Max: 1 | 0: 19926 (94.5%)<br>1: 1151 (5.5%) |
| **89** | **Amount of alcohol drunk on a typical drinking day** | Alcohol and cannabis use | Mean (SD): 0.8 (1.1)<br>min < med < max:<br>0 < 0 < 4<br>IQR (CV): 1 (1.3) | 0: 10819 (51.3%)<br>1: 5709 (27.1%)<br>2: 2560 (12.1%)<br>3: 1402 (6.7%)<br>4: 587 (2.8%) |
| **90** | **Frequency of drinking alcohol** | Alcohol and cannabis use | Mean (SD): 2.7 (1.2)<br>min < med < max:<br>0 < 3 < 4<br>IQR (CV): 2 (0.4) | 0: 1350 (6.4%)<br>1: 2450 (11.6%)<br>2: 3957 (18.8%)<br>3: 6694 (31.8%)<br>4: 6626 (31.4%) |
| **91** | **Frequency of consuming six or more units of alcohol** | Alcohol and cannabis use | Mean (SD): 1 (1.2)<br>min < med < max:<br>0 < 1 < 4<br>IQR (CV): 2 (1.2) | 0: 10321 (49.0%)<br>1: 5279 (25.0%)<br>2: 1969 (9.3%)<br>3: 2830 (13.4%)<br>4: 678 (3.2%) |
| **92** | **(log)AUDIT** | Alcohol and cannabis use | Mean (SD): 1.6 (0.7)<br>min < med < max:<br>0 < 1.6 < 3.6<br>IQR (CV): 1 (0.4) | 35 distinct values |
| **93** | **(log)AUDIT-C** | Alcohol and cannabis use | Mean (SD): 1.5 (0.6)<br>min < med < max:<br>0 < 1.6 < 2.6<br>IQR (CV): 0.8 (0.4) | 13 distinct values |
| **94** | **(log)AUDIT-P** | Alcohol and cannabis use | Mean (SD): 0.3 (0.7)<br>min < med < max:<br>0 < 0 < 3.1<br>IQR (CV): 0 (2) | 23 distinct values |
| **95** | **Hazardous alcohol use (AUDIT ≥ 8)** | Alcohol and cannabis use | Min: 0<br>Mean: 0.2<br>Max: 1 | 0: 15994 (75.9%)<br>1: 5083 (24.1%) |
| **96** | **Alcohol dependence (AUDIT ≥ 15)** | Alcohol and cannabis use | Min: 0<br>Mean: 0<br>Max: 1 | 0: 20038 (95.1%)<br>1: 1039 (4.9%) |
| **97** | **Felt hated by family member as a child** | Traumatic events | Mean (SD): 1.3 (0.7)<br>min < med < max:<br>1 < 1 < 5<br>IQR (CV): 0 (0.6) | 1: 17886 (84.9%)<br>2: 1377 (6.5%)<br>3: 1305 (6.2%)<br>4: 284 (1.3%)<br>5: 225 (1.1%) |

| | | | | |
|---|---|---|---|---|
| **98** | **Physically abused by family as a child** | Traumatic events | Mean (SD): 1.3 (0.7)<br>min < med < max:<br>1 < 1 < 5<br>IQR (CV): 0 (0.5) | 1: 17061 (80.9%)<br>2: 2416 (11.5%)<br>3: 1333 (6.3%)<br>4: 162 (0.8%)<br>5: 105 (0.5%) |
| **99** | **Felt loved as a child** | Traumatic events | Mean (SD): 4.2 (1)<br>min < med < max:<br>1 < 5 < 5<br>IQR (CV): 1 (0.2) | 1: 270 (1.3%)<br>2: 904 (4.3%)<br>3: 3436 (16.3%)<br>4: 5319 (25.2%)<br>5: 11148 (52.9%) |
| **100** | **Sexually molested as a child** | Traumatic events | Mean (SD): 1.1 (0.5)<br>min < med < max:<br>1 < 1 < 5<br>IQR (CV): 0 (0.4) | 1: 19165 (90.9%)<br>2: 1085 (5.1%)<br>3: 664 (3.2%)<br>4: 90 (0.4%)<br>5: 73 (0.3%) |
| **101** | **Someone to take to a doctor when needed as a child** | Traumatic events | Mean (SD): 4.8 (0.7)<br>min < med < max:<br>1 < 5 < 5<br>IQR (CV): 0 (0.1) | 1: 369 (1.8%)<br>2: 147 (0.7%)<br>3: 473 (2.2%)<br>4: 1875 (8.9%)<br>5: 18213 (86.4%) |
| **102** | **Ever sought or received professional help for mental distress** | Mental distress | Min: 0<br>Mean: 0.4<br>Max: 1 | 0: 12990 (61.6%)<br>1: 8087 (38.4%) |
| **103** | **Ever suffered mental distress preventing usual activities** | Mental distress | Min: 0<br>Mean: 0.3<br>Max: 1 | 0: 14265 (67.7%)<br>1: 6812 (32.3%) |
| **104** | **Belittlement by partner or ex-partner as an adult** | Traumatic events | Mean (SD): 1.4 (0.9)<br>min < med < max:<br>1 < 1 < 5<br>IQR (CV): 0 (0.6) | 1: 16092 (76.3%)<br>2: 2124 (10.1%)<br>3: 2051 (9.7%)<br>4: 440 (2.1%)<br>5: 370 (1.8%) |
| **105** | **Been in a confiding relationship as an adult** | Traumatic events | Mean (SD): 4.1 (1.2)<br>min < med < max:<br>1 < 5 < 5<br>IQR (CV): 2 (0.3) | 1: 1472 (7.0%)<br>2: 925 (4.4%)<br>3: 3568 (16.9%)<br>4: 3594 (17.1%)<br>5: 11518 (54.6%) |
| **106** | **Physical violence by a partner or ex-partner as an adult** | Traumatic events | Mean (SD): 1.2 (0.6)<br>min < med < max:<br>1 < 1 < 5<br>IQR (CV): 0 (0.5) | 1: 18528 (87.9%)<br>2: 1343 (6.4%)<br>3: 893 (4.2%)<br>4: 137 (0.6%)<br>5: 176 (0.8%) |
| **107** | **Sexual interference by a partner or ex-partner without consent as an adult** | Traumatic events | Mean (SD): 1.1 (0.4)<br>min < med < max:<br>1 < 1 < 5<br>IQR (CV): 0 (0.4) | 1: 19963 (94.7%)<br>2: 645 (3.1%)<br>3: 363 (1.7%)<br>4: 49 (0.2%)<br>5: 57 (0.3%) |
| **108** | **Able to pay rent/mortgage as an adult** | Traumatic events | Mean (SD): 4.8 (0.8)<br>min < med < max:<br>1 < 5 < 5<br>IQR (CV): 0 (0.2) | 1: 590 (2.8%)<br>2: 98 (0.5%)<br>3: 416 (2.0%)<br>4: 1351 (6.4%)<br>5: 18622 (88.4%) |

| | | | | |
|---|---|---|---|---|
| 109 | **Been in a serious accident believed to be life-threatening** | Traumatic events | Mean (SD): 0.1 (0.3)<br>min < med < max:<br>0 < 0 < 2<br>IQR (CV): 0 (3.1) | 0: 19056 (90.4%)<br>1: 1971 (9.4%)<br>2: 50 (0.2%) |
| 110 | **Been involved in combat or exposed to war-zone** | Traumatic events | Mean (SD): 0 (0.2)<br>min < med < max:<br>0 < 0 < 2<br>IQR (CV): 0 (5.2) | 0: 20322 (96.4%)<br>1: 741 (3.5%)<br>2: 14 (0.1%) |
| 111 | **Diagnosed with a life-threatening illness** | Traumatic events | Mean (SD): 0.1 (0.4)<br>min < med < max:<br>0 < 0 < 2<br>IQR (CV): 0 (3) | 0: 18727 (88.9%)<br>1: 2074 (9.8%)<br>2: 276 (1.3%) |
| 112 | **Victim of physically violent crime** | Traumatic events | Mean (SD): 0.2 (0.4)<br>min < med < max:<br>0 < 0 < 2<br>IQR (CV): 0 (2) | 0: 16838 (79.9%)<br>1: 4147 (19.7%)<br>2: 92 (0.4%) |
| 113 | **Witnessed sudden violent death** | Traumatic events | Mean (SD): 0.2 (0.4)<br>min < med < max:<br>0 < 0 < 2<br>IQR (CV): 0 (2.5) | 0: 18003 (85.4%)<br>1: 2956 (14.0%)<br>2: 118 (0.6%) |
| 114 | **Victim of sexual assault** | Traumatic events | Mean (SD): 0.1 (0.4)<br>min < med < max:<br>0 < 0 < 2<br>IQR (CV): 0 (2.4) | 0: 17950 (85.2%)<br>1: 3104 (14.7%)<br>2: 23 (0.1%) |
| 115 | **Physical alcohol dependence ever** | Addictions | Min: 0<br>Mean: 0<br>Max: 1 | 0: 20989 (99.6%)<br>1: 88 (0.4%) |
| 116 | **Substance addiction** | Addictions | Min: 0<br>Mean: 0<br>Max: 1 | 0: 20513 (97.3%)<br>1: 564 (2.7%) |
| 117 | **Current addiction** | Addictions | Min: 0<br>Mean: 0<br>Max: 1 | 0: 20698 (98.2%)<br>1: 379 (1.8%) |
| 118 | **Cannabis ever** | Alcohol and cannabis use | Min: 0<br>Mean: 0.2<br>Max: 1 | 0: 16058 (76.2%)<br>1: 5019 (23.8%) |
| 119 | **Lifetime frequency of taking cannabis** | Alcohol and cannabis use | Mean (SD): 0.5 (1)<br>min < med < max:<br>0 < 0 < 4<br>IQR (CV): 0 (2.1) | 0: 16058 (76.2%)<br>1: 2213 (10.5%)<br>2: 1310 (6.2%)<br>3: 977 (4.6%)<br>4: 519 (2.5%) |
| 120 | **Cannabis daily** | Alcohol and cannabis use | Min: 0<br>Mean: 0<br>Max: 1 | 0: 20804 (98.7%)<br>1: 273 (1.3%) |
| 121 | **Childhood adverse events** | Traumatic events | Min: 0<br>Mean: 0.5<br>Max: 1 | 0: 11417 (54.2%)<br>1: 9660 (45.8%) |
| 122 | **Adult adverse events** | Traumatic events | Min: 0<br>Mean: 0.5<br>Max: 1 | 0: 10486 (49.8%)<br>1: 10591 (50.2%) |
| 123 | **Catastrophic trauma** | Traumatic events | Min: 0<br>Mean: 0.5<br>Max: 1 | 0: 10686 (50.7%)<br>1: 10391 (49.3%) |
| 124 | **Any distress** | Mental distress | Min: 0<br>Mean: 0.5<br>Max: 1 | 0: 10100 (47.9%)<br>1: 10977 (52.1%) |

| | | | | | |
|---|---|---|---|---|---|
| 125 | **Happiness** | Happiness and subjective well-being | Mean (SD): 4.5 (0.7)<br>min < med < max:<br>1 < 5 < 6<br>IQR (CV): 1 (0.2) | 1: 20 (0.1%)<br>2: 96 (0.5%)<br>3: 672 (3.2%)<br>4: 9578 (45.4%)<br>5: 9381 (44.5%)<br>6: 1330 (6.3%) | 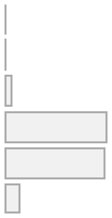 |
| 126 | **Health satisfaction** | Happiness and subjective well-being | Mean (SD): 4.4 (0.8)<br>min < med < max:<br>1 < 4 < 6<br>IQR (CV): 1 (0.2) | 1: 63 (0.3%)<br>2: 281 (1.3%)<br>3: 1290 (6.1%)<br>4: 9322 (44.2%)<br>5: 8741 (41.5%)<br>6: 1380 (6.5%) | 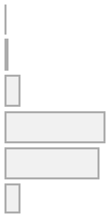 |
| 127 | **Family relationship satisfaction** | Happiness and subjective well-being | Mean (SD): 4.8 (0.8)<br>min < med < max:<br>1 < 5 < 6<br>IQR (CV): 1 (0.2) | 1: 73 (0.3%)<br>2: 173 (0.8%)<br>3: 855 (4.1%)<br>4: 5667 (26.9%)<br>5: 10204 (48.4%)<br>6: 4105 (19.5%) | 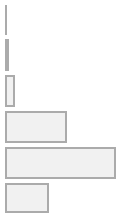 |
| 128 | **Friendships satisfaction** | Happiness and subjective well-being | Mean (SD): 4.8 (0.7)<br>min < med < max:<br>1 < 5 < 6<br>IQR (CV): 1 (0.2) | 1: 28 (0.1%)<br>2: 69 (0.3%)<br>3: 498 (2.4%)<br>4: 6262 (29.7%)<br>5: 11399 (54.1%)<br>6: 2821 (13.4%) | 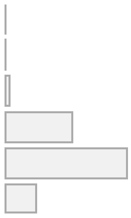 |
| 129 | **Financial situation satisfaction** | Happiness and subjective well-being | Mean (SD): 4.8 (0.8)<br>min < med < max:<br>1 < 5 < 6<br>IQR (CV): 1 (0.2) | 1: 73 (0.3%)<br>2: 169 (0.8%)<br>3: 711 (3.4%)<br>4: 6437 (30.5%)<br>5: 10155 (48.2%)<br>6: 3532 (16.8%) | 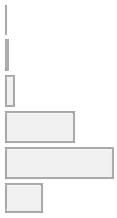 |
| 130 | **General happiness** | Happiness and subjective well-being | Mean (SD): 4.6 (0.8)<br>min < med < max:<br>1 < 5 < 6<br>IQR (CV): 1 (0.2) | 1: 41 (0.2%)<br>2: 143 (0.7%)<br>3: 873 (4.1%)<br>4: 8033 (38.1%)<br>5: 9784 (46.4%)<br>6: 2203 (10.5%) | 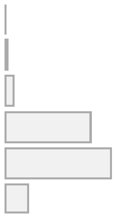 |
| 131 | **General happiness with own health** | Happiness and subjective well-being | Mean (SD): 4.5 (0.9)<br>min < med < max:<br>1 < 5 < 6<br>IQR (CV): 1 (0.2) | 1: 139 (0.7%)<br>2: 326 (1.5%)<br>3: 1523 (7.2%)<br>4: 8291 (39.3%)<br>5: 8597 (40.8%)<br>6: 2201 (10.4%) | 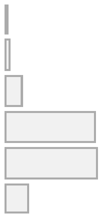 |
| 132 | **Belief that own life is meaningful** | Happiness and subjective well-being | Mean (SD): 3.7 (0.8)<br>min < med < max:<br>1 < 4 < 5<br>IQR (CV): 1 (0.2) | 1: 278 (1.3%)<br>2: 1145 (5.4%)<br>3: 5398 (25.6%)<br>4: 11504 (54.6%)<br>5: 2752 (13.1%) | 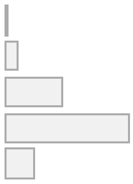 |
| 133 | **Wellbeing** | Happiness and subjective well-being | Mean (SD): 12.8 (1.9)<br>min < med < max:<br>3 < 13 < 17<br>IQR (CV): 2 (0.1) | 15 distinct values | 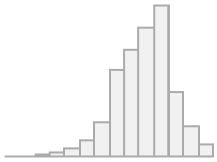 |

*SD* standard deviation, *IQR* interquartile range, *CV* coefficient of variation, *N-12* Eysenck Neuroticism score, *PTSD* posttraumatic stress disorder, *PCL-6* PTSD Checklist, *PHQ-9* Patient Health Questionnaire score, *PDS* Probable Depression Status, *RDS-4* Recent Depressive Symptoms, *AUDIT* Alcohol Use Disorders Identification Test, *GAD-7* Generalized Anxiety Disorder score, *GP* general practitioner.

### S3. Neuroimaging: MRI Acquisition and Preprocessing

MRI data were collected on a Siemens Magnetom Skyra 3T scanner with the Siemens 32-channel head coil across four imaging centres: Cheadle, Reading, Newcastle, and Bristol. The data were processed in FSL (FMRIB Software Library) and MRtrix [19].

#### Diffusion-weighted MRI (dwMRI)

dwMRI data were acquired with a SE-EPI sequence with ×3 multislice (multiband) acceleration in 100 distinct diffusion-encoding directions and two shells: 50 diffusion-encoding directions with $b$ = 1000 s/mm$^2$ and 50 diffusion-encoding directions with $b$ = 2000 s/mm$^2$ (TE = 92 ms, TR = 3600 ms, 2 mm isotropic voxel, partial Fourier 6/8, FOV: 104×104×72, $\delta$ = 21.4 ms) in two phase-encoding directions. Primary data were acquired with anterior-to-posterior phase encoding and three $b$ = 0 images were acquired with reversed phase encoding for fieldmap estimation and distortion correction.

**Imaging-derived phenotypes (IDPs)**

dwMRI IDPs provided by the UK Biobank were derived from head motion, eddy current, and gradient distortion-corrected dwMRI data. In diffusion tensor imaging (DTI) analysis, a diffusion tensor was fitted using DTIFit. This resulted in six diffusion metrics: fractional anisotropy (FA), mean diffusivity (MD), diffusion tensor mode (MO), and three eigenvalues of the diffusion tensor, L1, L2, and L3. Feeding preprocessed dwMRI data into Neurite Orientation Dispersion and Density Imaging (NODDI) resulted in three additional metrics: intracellular volume fraction (ICVF), isotropic or free water volume fraction (ISOVF), and the orientation dispersion index (OD) [20, 21].

Probabilistic tractography analysis was carried out in FLS using BEDPOSTx / PROBTRACKx. Diffusion metrics were mapped onto 27 fibre tracts using standard-space ROI masks defined by AutoPtx [22]. In probabilistic tractography, white matter tracts are reconstructed by building a probability distribution of connections between voxels to account for uncertainty in fiber orientation [23, 24]. FA images derived from fitting a diffusion tensor were also fed into Tract-Based Spatial Statistics (TBSS) [25]. The resulting skeletonized images and DTI/NODDI diffusion metrics were averaged across 48 standard-space tract masks from the Susumu Mori white matter atlas [26, 27]. Briefly, in TBSS, individual FA images are projected onto a standard-space mean FA skeleton to represent fibre tracts common to all subjects. The resulting standard-space warp is then applied to other microstructural indices. Skeletonized images are then averaged across white matter tracts defined by an atlas of interest. Having all indices in standard space allows between-subject comparisons [25].

FA values range from 0 to 1 and reflect directionality or the degree of anisotropy of water diffusion. MD reflects diffusion weighted along the three directions, i.e., directionality invariant diffusion rate [28–30]. MO is perpendicular to FA and quantifies tensor shape by measuring the extent to which anisotropy is planar as a flat cylinder or linear as a tube [31, 32]. L1, L2, and L3 reflect the magnitude of water diffusion along the three principal diffusion axes within a voxel [33]. ICVF measures water diffusion within axons or neurites and is a marker of neurite density. ISOVF reflects the freely diffusing water such as in cerebrospinal fluid (in CSF). OD measures neurite dispersion or spatial configuration. Higher values of OD correspond to grey matter where neurites are spread out in many different directions and lower OD values are characteristic of white matter where neurites are aligned in one direction [20, 34, 35].

**Structural connectomes**

A procedure for computing structural and functional connectomes is discussed in detail in Mansour et al. [23]. In brief, the researchers derived structural connectomes following the Anatomically-Constrained Tractography (ACT) with Fiber Orientation Distributions (iFOD2) framework [36]. In this method, fibres or streamlines are propagated from single seed points set from within white matter or at the grey matter/white matter interface based on local fibre orientation distributions [37] or using dynamic seeding (Spherical-Deconvolution Informed Filtering of Tractograms, SIFT and SIFT2) [38, 39]. SIFT and SIFT2 address a streamline reconstruction bias that states that streamlines density is not representative of the density of underlying biological pathways [38, 39]. These algorithms improve the biological accuracy of streamline reconstruction by identifying a subset of streamlines that best align with the diffusion signal. To filter out certain streamlines, this technique leverages spherical deconvolution results (hence "spherical-deconvolution informed filtering of tractograms"). SIFT2 is usually referred to as "fibre bundle capacity" because it assigns weights to each streamline in a tractogram to ensure that the density of streamlines matches the underlying fibre density estimated from the diffusion signal. These weights represent the fibre bundles' cross-sectional area. The sum of these weights within a bundle quantifies the bundle's total intracellular cross-sectional area and connectivity, thus aligning the streamline density with the actual fibre density [40].

First, Mansour and colleagues performed skull stripping on the averaged $b = 0$ volumes using FSL *bet* to improve registration accuracy between the T1w and dwMRI images. Macroscopic tissue response functions for white and grey matter and cerebrospinal fluid were used together with fibre orientation distributions for intensity normalization and bias field correction. Whole-brain probabilistic tractography was carried out within the ACT framework based on tissue-type segmentation obtained with FreeSurfer and FSL FIRST. Streamline seeding was initiated at the interface between grey and white matter. This resulted in 10 million streamline seeds that satisfied the length constraints of the ACT [36, 37]. Whole-brain streamlines were then resampled using cortical and subcortical atlases. For a given grey matter parcellation, in addition to DTI indices averaged within each set of streamlines, it is possible to count the number and the mean length of streamlines connecting each pair of regions. Mansour and colleagues parcellated individual structural brain images into cortical and subcortical grey matter nodes based on cortical and subcortical atlases defined in template space and transformed into each subject's native space [36, 41–46]. For subcortical parcellations, the inverse of the native T1w to MNI152 template warp was obtained and applied to the four subcortical parcellations using FSL's *applywarp*. For cortical atlases, the surface-based labels were aligned to the cortical surfaces in the individual's native space using FreeSurfer's *fsnative*. Cortical and subcortical parcellations were derived independently and were combined before structural connectome construction.

#### Resting-state MRI (rsMRI)

rsMRI was acquired with a gradient echo EPI sequence with ×8 multislice acceleration (TE = 39 ms, TR = 735 ms, 2.4 mm isotropic voxel, 490 volumes, FOV: 88×88×64, flip angle 52$^{o}$). According to the UK Biobank pipeline, rsMRI data preprocessing included correction for artifacts, i.e., susceptibility and gradient distortion correction, motion correction (Motion Correction using FMRIB's Linear Image Registration Tool), grand mean scaling, high-pass temporal filtering, and structured artifact removal with Independent Component Analysis (ICA) followed by FMRIB's ICA-based X-noiseifier (FIX), and discarding non-neural components. Group ICA was performed using FSL MELODIC (Multivariate Exploratory Linear Optimized Decomposition into Independent Components) ICA to obtain spatial maps of resting state networks and was followed by dual regression analysis to estimate the subject-specific Blood Oxygenation Level Dependent (BOLD) time series for each network [47–50].

**Imaging-derived phenotypes (IDPs)**

Resting-state functional connectivity (RSFC) is usually measured as full of partial correlations between each pair of brain regions defined by a given grey matter parcellation (atlas) or between independent components (ICs). ICs are statistically independent or spatially distinct resting-state functional networks derived from ICA [51]. rsMRI IDPs were comprised of the outputs of ICA applied at 25 and 100 dimensions. Removal of “bad” ICs resulted in 21 and 55 networks. For both normalized temporal and partial temporal correlations between ICs time series were calculated. Unlike full correlation, partial correlation coefficients allow evaluation of the strength of direct connections between each node pair. Partial correlation coefficients were additionally regularized using Ridge regression (L2 regularization, *rho* = 0.5). All correlation coefficients were converted into *z*-statistics (Gaussianised) and corrected for temporal autocorrelation.

**Functional connectomes**

To obtain functional connectomes, Mansour and colleagues parcellated motion- and artifact-corrected resting state time series data by averaging the BOLD time series within regions defined by parcellations of interest. The parcellations were resampled from the individual’s T1w image to the fMRI voxel grid using FreeSurfer’s *mri_vol2vol*. The BOLD time series were then averaged across the fMRI voxels within each parcellation. In our study, we derived functional connectivity metrics from the BOLD time series using the *ConnectivityMeasure* function of the *Nilearn Python library* (https://nilearn.github.io/dev/modules/generated/nilearn.connectome.ConnectivityMeasure.html).

For both the rsMRI IDPs and BOLD time series, in addition to full and partial correlations, we computed tangent space parameterization matrices. Computation of covariance tangent space parameterization requires derivation of a group average covariance (group connectivity matrix or the group mean set to 0) from the individual subject covariances in the train set using the Ledoit-Wolf estimator and transformation of these covariance matrices in the train and test sets [52, 53]. To do that, we applied tangent space parameterization to ICA node or cortical region of interest (ROI) time series with *z*-score standardization which shifts time series to zero means and scales them to unit variance. The matrices were vectorized and diagonal values were discarded.

The advantage of tangent space parameterization resides in the fact that functional connectivity measures in the form of vectorized correlation matrices are not Euclidean objects and lie on a high-dimensional nonlinear curved surface called the symmetric positive definite or Riemannian manifold [54, 55]. The projection of covariances onto a common tangent space via Riemannian geometry reduces statistical dependencies between the covariance estimates and allows the application of conventional algebraic operations on functional connectivity matrices. The method often outperforms Pearson correlation by overcoming limitations associated with the non-linear geometry of conventional correlation matrices [52, 54–58].

For the BOLD time series of each node pair in each atlas combination, full and partial correlations were calculated within the whole sample, and tangent space parameterization matrices were obtained by fitting the function to the train set and applying it to the train and test sets in each fold. We additionally applied the inverse hyperbolic tangent function (*arctanh*) to Pearson correlation coefficients to reduce skewness.

#### Structural MRI (sMRI)

T1-weighted (T1w) scans were acquired with a 3D MPRAGE sequence (1 mm isotropic voxel, FOV: 208×256×256). Overall, structural image preprocessing included gradient distortion correction, brain extraction and defacing, and registration of images in native space to MNI152 standard space using linear and non-linear affine registration [19, 36, 59–61]. Tissue and total brain volumes were obtained with SIENAX-style analysis (Structural Image Evaluation, using Normalization, of Atrophy: Cross-sectional) [62]. Shapes and volumes of 15 subcortical structures were modeled using FIRST (FMRIB's Integrated Registration and Segmentation Tool) [63]. Cortical surface area and thickness were estimated in FreeSurfer, and subcortical regions were extracted using FreeSurfer's ASEG tool [64].

T2-weighted (T2w) scans were obtained using the 3D SPACE acquisition with fluid-attenuated inversion recovery (FLAIR) contrast to enhance white matter hyperintensities visualization (1.05×1×1 mm voxel, FOV: 192×256×256). T2w images were linearly aligned to T1w images using FSL FLIRT and then to MNI and were bias-corrected.

T1w and T2-weighted (T1w)-FLAIR scans were used to evaluate the total volume of white matter hyperintensities i.e., white matter lesions (Brain Intensity AbNormality Classification Algorithm,

BIANCA) [65]. Tissue segmentation and bias-field correction were performed in FSL using FAST (FMRIB's Automated Segmentation Tool) [66].

**Table S6.** Summary of the neuroimaging phenotypes used as features in machine learning models

| MRI Modality | IDP | N features | Field / Category ID |
|---|---|---|---|
| | **T1w sMRI** | | |
| **sMRI UK Biobank IDPs** | **Parcellated T1w indices** | | |
| | Regional grey matter volumes (FAST) | **139** | 1101 |
| | Subcortical volumes (FIRST) | **14** | 1102 |
| | FreeSurfer ASEG: | **95** | 190 |
| | • mean intensity | 41 | |
| | • volume | 54 | |
| | FreeSurfer BA *ex vivo* | **84** | 195 |
| | • area | 28 | |
| | • mean thickness | 28 | |
| | • volume | 28 | |
| | FreeSurfer a2009s | **444** | 197 |
| | • area | 148 | |
| | • mean thickness | 148 | |
| | • volume | 148 | |
| | FreeSurfer DKT | **186** | 196 |
| | • area | 62 | |
| | • mean thickness | 62 | |
| | • volume | 62 | |
| | FreeSurfer Desikan | **338** | |
| | • grey/white contrast | 70 | 194 |
| | • pial surface area | 66 | 193 |
| | • white matter area | 68 | 192 |
| | • white matter mean thickness | 68 | 192 |
| | • white matter volume | 66 | 192 |
| | FreeSurfer subcortical subsegmentation | 121 | 191 |
| | **Whole-brain T1w indices** | | |
| | Volume of brain, grey and white matter | 1 | 25010 |
| | Volume of white matter | 1 | 25008 |
| | Volume of grey matter | 1 | 25006 |
| | Volume of peripheral cortical grey matter | 1 | 25002 |
| | Volume of ventricular cerebrospinal fluid | 1 | 25004 |

| | | | |
|---|---|---|---|
| | Volume of the brain stem and 4th ventricle | 1 | 25025 |
| | Volume-ratio of BrainSegVol-to-eTIV (whole brain) | 1 | 26536 |
| | Volume-ratio of MaskVol-to-eTIV (whole brain) | 1 | 26537 |
| | **T2w sMRI** | | |
| **sMRI UK Biobank IDPs** | Total volume of deep white matter hyperintensities<br>Total volume of peri-ventricular white matter hyperintensities<br>Total volume of white matter hyperintensities (from T1w and T2w FLAIR images) | 1<br>1<br>1 | 24486<br>24485<br>25781 |
| | **dwMRI** | | |
| **dwMRI UK Biobank IDPs** | **TBSS (tract skeleton measurements):**<br>FA, MD, MO, L1, L2, L3, OD, ICVF, ISOVF | 432:<br>48×9 | 134 |
| | **Probabilistic tractography (weighted means of the parameter within the tract):**<br>FA, MD, MO, L1, L2, L3, OD, ICVF, ISOVF | 243:<br>27×9 | 135 |
| **Structural connectomes from parcellated data** | • Streamline count<br>• Fibre bundle capacity (from SIFT2)<br>• Mean streamline length<br>• Mean FA<br>*for*<br>Glasser and Tian Subcortex S1 3T<br>Glasser and Tian Subcortex S4 3T<br>Schaefer7n200p and Tian Subcortex S1 3T<br>Schaefer7n500p and Tian Subcortex S4 3T<br>aparc a2009s and Tian Subcortex S1 3T<br>aparc and Tian Subcortex S1 3T | 376×376 (141188)<br>414×414 (85491)<br>216×216 (23220)<br>554×554 (153181)<br>164×164 (13366)<br>84×84 (3486) | 31022<br>31023<br>31025<br>31026<br>31020<br>31021 |
| | **rsMRI** | | |
| **rsMRI UK Biobank IDPs** | **Dimension 25, 21 nodes:** | | |
| | • Full correlation matrix | 210 | 25750 |
| | • Partial correlation matrix | 210 | 25752 |
| | • *Tangent space parameterization matrix | 210 | 20227 |
| | • Resting-state component amplitudes | 210 | 25754 |

| | | | |
|---|---|---|---|
| | **Dimension 100, 55 nodes:** | | |
| | • Full correlation matrix | 1485 | 25751 |
| | • Partial correlation matrix | 1485 | 25753 |
| | • *Tangent space parameterization matrix | 1485 | 20227 |
| | • Resting-state component amplitudes | 1485 | 25755 |
| | fMRI time series: Glasser | | 31016 |
| | fMRI time series: Schaefer7n200p & Schaefer7n500p | | 31018 |
| | fMRI time series: Tian Subcortex S1 to S4 3T | | 31019 |
| | fMRI time series: aparc | | 31015 |
| | fMRI time series: aparc a2009s | | 31014 |
| | | 376×376 | |
| **Functional connectomes from parcellated data** | • *Full correlation matrix | (141188) | |
| | • *Partial correlation matrix | 414×414 | |
| | • *Tangent space parameterization matrix | (85491) | – |
| | *from BOLD time series for:* | 216×216 | – |
| | Glasser and Tian Subcortex S1 3T | (23220) | – |
| | Glasser and Tian Subcortex S4 3T | 554×554 | – |
| | Schaefer7n200p and Tian Subcortex S1 3T | (153181) | – |
| | Schaefer7n500p and Tian Subcortex S4 3T | 164×164 | – |
| | aparc a2009s and Tian Subcortex S1 3T | (13366) | |
| | aparc and Tian Subcortex S1 3T | 84×84 | |
| | | (3486) | |

**computed manually (i.e., not provided by the UK Biobank)*

*IDP* imaging-derived phenotype, *T1w* T1-weighted structural MRI, *T2w* T2-weighted structural MRI, *dwMRI* diffusion-weighted MRI, *rsMRI* resting-state functional MRI, *FA* fractional anisotropy, *MD* mean diffusivity, *MO* diffusion tensor mode, *L1*, *L2*, *L3* eigenvalues of the diffusion tensor, *OD* orientation dispersion index, *ICVF* intracellular volume fraction, *ISOVF* isotropic volume fraction, *BrainSegVol* sum of the volume of the structures, *eTIV* estimated total intracranial volume, *SIFT2* Spherical-Deconvolution Informed Filtering of Tractograms, *DKT* Desikan-Killiany-Tourville, *BA* Brodmann Area Maps.

#### S4. Neuroimaging: MRI Confounds

A common set of confounds included scanning site (to account for site-specific variability), MRI acquisition date, head size, scanner table and radiofrequency receive coil positions, scanner brain position, the discrepancy between T1w brain image and standard-space brain template, and structural motion (a measure of head motion in T1w structural image) [67]. Head size is a volumetric scaling factor derived when transforming from the native T1w to the standard MNI template with SIENAX [62, 68]. We also converted UK Biobank's imaging centres which included four sites into a binary confound variable using dummy encoding, and the date of attending the assessment centre into Unix format.

Modality-specific confounds included the total amount of head motion calculated as the mean displacement for each consecutive timepoint pair (averaged across the brain and all time points), intensity scaling, and the discrepancy between non-structural brain image and T1w brain image. Motion estimates included the median absolute and relative head motions.

For dwMRI data, motion estimates were obtained with FSL *Eddy*. To control for slow effects related to heating we also used confounds associated with a more robust version of eddy current correction such as a necessity to use an increased search space in eddy current estimation. An additional motion-related variable for dwMRI included the total number of slices which *Eddy* considered outliers.

For rsMRI data, motion estimates were computed in FSL FEAT. An additional estimate of the head motion for rsMRI was obtained using the DVARS approach [69]. This approach is based on a decomposition of the total and global variability of the data into fast (D-var), slow (S-var), and edge (E-var) components. D-var (differenced variability) is defined as the sum of squares of the half difference between the two consecutive time points or adjacent scans at each voxel averaged over space; S-var corresponds to the sum of squares of the average of adjacent scans; E-var is computed as the edge sum of squares at times 1 and T. DVARS-related set of confounds therefore comprised of the mean, median and 90th percentile of S-var and D-var normalized by the total variability (A-var) [69].

Confounds were regressed from MRI features following standardization of confounds and features. Interactive violin plots of the variance explained by each confound group can be found at https://www.fmrib.ox.ac.uk/ukbiobank/confounds/plots_2020_03_11/index.html.

**Table S7.** Confound variables regressed from MRI data

| Confound Variable | Field ID |
|---|---|
| **dwMRI** | |
| **Outliers detected by Eddy** | |
| Number of dwMRI outlier slices detected and corrected | 25746 |
| Standard deviation of apparent translation in the Y axis measured by Eddy (Y-Translation) | 25922 |
| Number of slices that Eddy estimated to be outliers in dwMRI data | 24456 |
| Whether increased search space in Eddy current estimation was used for dwMRI (“NewEddy”) | 25921 |
| **Head Motion** | |
| Median absolute head motion from dwMRI | 24451 |
| Median relative head motion from dwMRI | 24454 |
| Structural motion | 24419 |
| **Table position** | |
| Scanner lateral (X) brain position (X-position of centre-of-gravity of brain mask in scanner coordinates) | 25756 |
| Scanner transverse (Y) brain position (Y-position of centre-of-gravity of brain mask in scanner coordinates) | 25757 |
| Scanner longitudinal (Z) brain position (Z-position of centre-of-gravity of brain mask in scanner coordinates) | 25758 |
| Scanner table position (Z-coordinate of the coil and the scanner table that the coil sits on within the scanner. The Z axis points down the centre of the magnet) | 25759 |
| **T1w-related confounds** | |
| Volumetric scaling from T1 head image to standard space (Head size) | 25000 |

| | |
|---|---|
| Discrepancy between T1 brain image and standard-space brain template (linearly aligned) | 25731 |
| Discrepancy between T1 brain image and standard-space brain template (nonlinearly-aligned) | 25732 |
| Discrepancy between dwMRI brain image and T1 brain image | 25737 |
| Intensity scaling for dwMRI | 25928 |
| **Assessment centre** | |
| Acquisition date | 53 |
| Site | 54 |
| **sMRI (parcellated T1w sMRI)** | |
| **Head Motion** | |
| Structural motion | 24419 |
| **Table position** | |
| Scanner lateral (X) brain position (X-position of centre-of-gravity of brain mask in scanner coordinates) | 25756 |
| Scanner transverse (Y) brain position (Y-position of centre-of-gravity of brain mask in scanner coordinates) | 25757 |
| Scanner longitudinal (Z) brain position (Z-position of centre-of-gravity of brain mask in scanner coordinates) | 25758 |
| Scanner table position (Z-coordinate of the coil (and the scanner table that the coil sits on) within the scanner. The Z axis points down the centre of the magnet) | 25759 |
| **T1w-related confounds** | |
| Volumetric scaling from T1 head image to standard space (Head size) | 25000 |
| Discrepancy between T1 brain image and standard-space brain template (linearly aligned) | 25731 |
| Discrepancy between T1 brain image and standard-space brain template (nonlinearly-aligned) | 25732 |
| Amount of warping applied to non-linearly align T1 brain image to standard-space | 25733 |
| Inverted signal-to-noise ratio in T1 | 25734 |
| Inverted contrast-to-noise ratio in T1 | 25735 |
| Intensity scaling for T1 | 25925 |

**Assessment centre**

| | |
|---|---|
| Acquisition date | 53 |
| Site | 54 |

**sMRI**
**(whole-brain T1w and T2w sMRI)**

**Head Motion**

| | |
|---|---|
| Structural motion | 24419 |

**Table position**

| | |
|---|---|
| Scanner lateral (X) brain position (X-position of centre-of-gravity of brain mask in scanner coordinates) | 25756 |
| Scanner transverse (Y) brain position (Y-position of centre-of-gravity of brain mask in scanner coordinates) | 25757 |
| Scanner longitudinal (Z) brain position (Z-position of centre-of-gravity of brain mask in scanner coordinates) | 25758 |
| Scanner table position (Z-coordinate of the coil (and the scanner table that the coil sits on) within the scanner. The Z axis points down the centre of the magnet) | 25759 |

**T1w-related confounds**

| | |
|---|---|
| Volumetric scaling from T1 head image to standard space (Head size) | 25000 |
| Discrepancy between T1 brain image and standard-space brain template (linearly aligned) | 25731 |
| Discrepancy between T1 brain image and standard-space brain template (nonlinearly-aligned) | 25732 |
| Amount of warping applied to non-linearly align T1 brain image to standard-space | 25733 |
| Inverted signal-to-noise ratio in T1 | 25734 |
| Inverted contrast-to-noise ratio in T1 | 25735 |
| Intensity scaling for T1 | 25925 |

**T2w-related confounds**

| | |
|---|---|
| Discrepancy between T2 FLAIR and T1 brain images | 25736 |
| Intensity scaling for T2 FLAIR | 25926 |

| | |
|---|---|
| **Assessment centre** | |
| Acquisition date | 53 |
| Site | 54 |
| **rsMRI** | |
| **Head Motion** | |
| Structural motion | 24419 |
| Median absolute head motion from rsMRI | 24439 |
| Median relative head motion from rsMRI | 24442 |
| Mean rsMRI head motion, averaged across space and time points | 25741 |
| **Table position** | |
| Scanner lateral (X) brain position (X-position of centre-of-gravity of brain mask in scanner coordinates) | 25756 |
| Scanner transverse (Y) brain position (Y-position of centre-of-gravity of brain mask in scanner coordinates) | 25757 |
| Scanner longitudinal (Z) brain position (Z-position of centre-of-gravity of brain mask in scanner coordinates) | 25758 |
| Scanner table position (Z-coordinate of the coil (and the scanner table that the coil sits on) within the scanner. The Z axis points down the centre of the magnet) | 25759 |
| **T1w-related confounds** | |
| Volumetric scaling from T1 head image to standard space (Head size) | 25000 |
| Discrepancy between T1 brain image and standard-space brain template (linearly aligned) | 25731 |
| Discrepancy between T1 brain image and standard-space brain template (nonlinearly-aligned) | 25732 |
| Discrepancy between rsMRI brain image and T1 brain image | 25739 |
| **rsMRI-specific confounds** | |
| DVARS Mean S from cleaned rsMRI | 24429 |
| DVARS Median S from cleaned rsMRI | 24430 |
| DVARS 90th percentile S from cleaned rsMRI | 24431 |
| DVARS Mean D from cleaned rsMRI | 24432 |

| | |
|---|---|
| DVARS Median D from cleaned rsMRI | 24433 |
| DVARS 90th percentile D from cleaned rsMRI | 24434 |
| DVARS Mean SD from cleaned rsMRI | 24435 |
| DVARS 90th percentile SD from cleaned rsMRI | 24437 |
| Inverted temporal signal-to-noise ratio in pre-processed rsMRI | 25743 |
| Inverted temporal signal-to-noise ratio in artifact-cleaned pre-processed rsMRI | 25744 |
| Echo Time for rsMRI | 25923 |
| Intensity scaling for rsMRI | 25929 |
| **Assessment centre** | |
| Acquisition date | 53 |
| Site | 54 |

### S5. First-Level Model: Partial Least Squares Regression (PLSR)

#### PLSR

The first-level machine learning model applied to mental health and MRI data was PLSR. PLSR is a multivariate regression method that enables the assessment of linear relationships between a large number of correlated predictor variables and single or multiple response variables. Unlike Principal Component Analysis which is applied solely to features (i.e., predictor variables), PLSR performs dimensionality reduction of the features and makes predictions on the target variable(s) simultaneously

PLSR addresses the problem of feature multicollinearity by decomposing the features into orthogonal scores and loadings and regressing the response variable on these scores or latent variable estimates of the predictor variables [70]. It decomposes the feature matrix into orthogonal scores and loadings considering information on both the response, Y, and predictor, X, variable such that the response variable will be then regressed on scores but not predictors themselves. To achieve this, the PLSR model finds a new set of predictor variables, X-scores, which are the weighted linear combination of the original predictor variables, via rotation of the original X variables:

$$X = TP' + E, \text{ with } T = XW^{*}, \tag{1}$$

where T – matrix of X-scores ($N_{observations} \times A_{components}$) with columns $t_a$ (X-scores of component a); P – the matrix of loadings with columns $p_a$; E – matrix of X-residuals ($N_{observations} \times K_{predictors}$); W – matrix of X-weights ($K_{predictors} \times A_{components}$) with the columns $w_a$ (weights that are independent between components); $W^*$ – matrix of transformed (rotated) PLSR weights ($K_{predictors} \times A_{components}$) with columns $w_a^*$ ($W^* = W(P'W)^{-1}$)) [70].

The new predictor variables are the latent variable estimates (i.e., they represent X in the latent space), and predictors of the response variable, Y. X-scores, T, are the projections of X onto the latent variables. Weights, W, are used to calculate the scores, T, from the original predictor matrix, X. They are linear combinations of the original predictor variables which determine how much each original variable contributes to the latent variables (scores). Weights are computed by maximizing the covariance between the X and the response variable, Y, and then normalized such that the X-scores have a unit variance. Loadings, P, are the weights that describe the relationship between the original variables and the latent variables (scores). In other words, loadings determine how the original variables are represented in the latent space. Loadings are obtained by projecting the original variables onto the latent variables (scores).

The PLSR model assumes that the response and predictor variables can be modeled by the same latent variable [70]. Similarly, in the case of multiple response variables, Y-scores multiplied by corresponding weights are a linear combination of the original values of the response variable:

$$Y = UC' + G, \text{ with } U = YC \tag{2}$$

where U – matrix of Y-scores ($N_{observations} \times A_{components}$) with columns $u_a$ (Y-scores of component a); C – matrix of Y-weights ($M_{targets} \times A_{components}$) with columns $c_a$; G – the number of cross-validation groups [70].

X-scores are therefore predictors of Y:

$$Y = TC' + F, \quad (3)$$

where T – matrix of X-scores ($N_{observations} \times A_{components}$) with columns $t_a$ (X-scores of component a); C – matrix of Y-weights ($M_{targets} \times A_{components}$) with columns $c_a$; F – matrix of Y-residuals ($N_{observations} \times M_{targets}$).

Finally, a multiple regression model equation is as follows:

$$Y = XW^{*}C' + F = XB + F, \quad (4)$$

where B – matrix of regression coefficients for all response variables ($K_{predictors} \times M_{targets}$), and

$$B = W^{*}C'. \quad (5)$$

If there are no correlations among predictor variables (i.e., X matrix is orthogonal) and there is only one response variable, the PLSR result is a multiple regression solution with one component. After computing each component, the X-matrix is deflated by subtracting the modified values of X from the original X, such that the X-scores are expressed as weighted residuals left after the previous dimension [70]. In this way, the first weight vector of the X is the first eigenvector of the combined variance-covariance matrix for X and Y (X′YY′X), and the weight vectors for further components are eigenvectors of the deflated matrix. Weights, $w_a$, are orthonormal, X-scores, $t_a$, are orthogonal, and X-loadings, $p_a$, and Y-scores, $u_a$, are not orthogonal to each other but orthogonal to X-scores, $t_a$, and X-weights, $w_a$, from previous components.

To put it simply, PLSR calculates the “new” predictor variables or X-scores which are (1) the latent variable estimates, (2) a linear combination of the real X-variables, and (3) predictors of the response variable, Y. The X-weights reflect the quantitative relationship between X and Y and provide information about which of the X-variables are important or similar. However, weights do not have to be interpreted directly as they may reflect the true correlations between the predictor and response variables and the correlations required to predict Y from variation in X that is unrelated to Y. In addition to the residuals of Y which may be used to characterize the model (large Y-residuals indicate a poor model), PLSR provides X-residuals that can be used to detect outliers in predictor variables.

Geometrically, X-scores, $t_a$, are coordinates of the X-matrix projection onto the hyperplane with dimension A that approximates X. The hyperplane is defined by one line and one direction per component such that X-loadings, $p_a$, correspond to the direction coefficients of these lines or slopes, and these lines define the best correlation with Y and the plane [70]. A comprehensive overview of the PLSR concept and its mathematical and geometric interpretation is given at https://learnche.org/pid/latent-variable-modelling/projection-to-latent-structures/index.

The only hyperparameter in PLSR is the number of components. Therefore, the tuning grid included only one parameter – the number of components to iterate over. To select the optimal number of components and predict the *g*-factor, we applied nested 10-fold cross-validation (CV) with random shuffling. Nested CV implies that within each of the five outer folds, the train sets are subdivided into 10 inner folds such that 90% of the data are used as an inner-fold training set for hyperparameter tuning and 10% of the data are used as a validation infer-fold set for hyperparameter selection.

To select the best hyperparameters, i.e., the optimal number of components, we used the *GridSearchCV* function of the *scikit-learn* library:

*model = GridSearchCV(estimator = PLSRegression(), param_grid={'n_components': range(1, X_train.shape[1] + 1)}, scoring='mean_absolute_error', cv=KFold(n_splits=10, shuffle=True, random_state=seed), verbose=4)*

The best hyperparameters were selected based on the mean absolute error (*MAE*) whose values range from negative infinity to zero with less negative values indicating better performance [71]. We then obtained out-of-sample predictions from unseen test data in each outer fold.

#### Performance metrics

To evaluate model performance (goodness of fit) and generalizability, we computed the coefficient of determination (*$R^2$*) which reflects the proportion of the variance in the target (dependent variable) explained by the features (independent variables), the Pearson correlation coefficient (*r*), *MAE,* and the mean squared error (*MSE*) for the values of the *g*-factor predicted from unseen features and original values of the *g*-factor (i.e., derived from ESEM) in each outer-fold test set according to the following formulas [72]:

$$R^2 = 1 - \frac{\sum_{i=1}^{n}(y_i - \hat{y}_i)^2}{\sum_{i=1}^{n}(y_i - \bar{y}_i)^2}, \quad (6)$$

$$r = \frac{\sum_{i=1}^{n}(y_i - \bar{y}_i)(\hat{y}_i - \bar{\hat{y}}_i)}{\sqrt{\sum_{i=1}^{n}(y_i - \bar{y}_i)^2 \sum_{i=1}^{n}(y_i - \hat{y}_i)^2}}, \quad (7)$$

$$MAE = \frac{1}{n}\sum_{i=1}^{n}|(y_i - \hat{y}_i)|, \quad (8)$$

$$MSE = \frac{1}{n}\sum_{i=1}^{n}(y_i - \hat{y}_i)^2, \quad (9)$$

where $y_i$, $\bar{y}i$, $\hat{y}i$ are the observed, observed mean, and predicted response variables, respectively.

To evaluate the performance of MRI-based models, we averaged the goodness-of-fit metrics across 5 folds for each set of features. For functional connectivity metrics, we selected those that demonstrated the best performance in the training set.

### S6. Second-Level Model: Stacking

For the second-level model, we combined (stacked) values of the *g*-factor predicted with PLSR from individual neuroimaging phenotypes within each and across all MRI modalities and used them as features. As such, we got four additional models: “rsMRI stacked”, “dwMRI stacked”, “sMRI stacked”, and “All MRI Stacked”. We used four machine learning algorithms to train the second-level models – ElasticNet, Random Forest, XGBoost, and Support Vector Regression, – and for further analyses selected the one which resulted in the highest out-of-sample *$R^2$*, as follows:

*model = GridSearchCV(regressor, parameter_grid, cv=5, verbose = 4)*

#### ElasticNet

ElasticNet is a linear regression model that combines Ridge and Lasso regression by incorporating L1 and L2 regularization terms to control for multicollinearity. The L1 penalty from Lasso regression can pick one of the correlated parameters by zeroing out less important ones which results in more sparse solutions and variable selection, whereas the L2 penalty from Ridge regression makes predictions less sensitive to a single feature by shrinking the slope asymptotically close to zero and thus guarantees even shrinkage of the regression coefficients [73–76]. In particular, ElasticNet addresses three main disadvantages of the Lasso: it handles situations when there are more predictors than observations; it does not eliminate all but one predictor out of a group of correlated predictors; it reduces the prediction error in situations when there are more observations than predictors that are multicollinear [77]. Furthermore, it makes the model more sparse or parsimonious by resolving the problem of redundancy characteristic for Ridge regression which tends to keep all predictors in the model even if they are highly correlated (i.e., continuous shrinkage) [73].

The hyperparameters for the ElasticNet model (*regressor = ElasticNet()*) included the maximum number of iterations (*max_iter*), alpha and l1 ratio, as indicated in the parameter grid:

- alpha: np.logspace(-6, 4, 500)
- l1_ratio: np.linspace(0, 1, 100)
- max_iter: 1000

*Alpha* is a scalar value that multiplies the penalty terms and therefore controls the degree of the regularization such that a larger value enhances the regularization and decreases the chance of overfitting, and a lower value approximates the ElasticNet model to the ordinary least square model. *l1 ratio* defines the contributions of L1 and L2 regularization terms such that at *l1 ratio* of 0 there is an L2 penalty (Ridge regression) only and at a value of 1 there is L1 penalty (Lasso regression) only; at 0 < *l1 ratio* < 1 the penalty is a combination of L1 and L2.

#### Support Vector Regression

Support Vector Regression (SVR) is a generalization of the Support Vector Machines (SVM), a supervised learning method introduced in 1979 by Vapnik [78, 79]. In binary classification problems, SVM can capture nonlinear patterns and deal with a large number of predictors along with a small number of observations by transforming the original data into a hyperplane or high-dimensional feature space (which is at least one dimension less than the original feature space is) using a so-called kernel trick which is a way of calculating scalar (inner) products between all pairs of data representations in nonlinear high-dimensional feature spaces [78, 80–82]. In the SVM context, the classification problem is formulated as a convex optimization problem that aims to find the maximum margin separating the hyperplane to enable the correct classification of as many training data points as possible [80, 83]. The optimal hyperplane is therefore considered a classification boundary that separates two classes; the margin of the hyperplane is defined as the distance between two classes in the original feature space; support vectors (SV) define the margin of the hyperplane by finding the data points of one class that are the closest to the other class. Instead of directly mapping original data onto a higher-dimensional space, non-linear SVMs are derived using a kernel function that calculates a modified inner product of vectors from the original feature space in a higher-dimensional space (the kernel trick). Once observations are mapped, the predictions of the class for new data points are made based on the side of the gap they fall [82].

In SVR, an equivalent to the SVM's margin is the ε-insensitive region around the hyperplane or the $\varepsilon$-insensitive tube that defines the degree to which deviations in predictions are tolerated (i.e., a margin of tolerance) such that deviations that are larger than $\varepsilon$ (i.e., farther than $\varepsilon$ from the actual values) are penalized. This is achieved by minimizing the $\varepsilon$-insensitive loss function to find the flattest and the narrowest $\varepsilon$-tube that contains the majority of the training data points and minimizes prediction errors (i.e., the geometrical distance between the predicted and the real values). The width of the $\varepsilon$-tube is defined by the two parallel boundaries, one above and one below the hyperplane, and $\varepsilon$ is therefore the distance from boundaries to the hyperplane (such that tube width is equal to $2\varepsilon$). SVs in this context are the training instances that fall outside the $\varepsilon$-tube boundary and define the shape of the *ε*-tube as well as the position and orientation of the regression hyperplane that runs through the middle of the *ε*-tube (output of the SVR model) [80, 81].

In the hyperparameter tuning grid for SVR (*regressor = SVR()*), we specified three parameters, namely *kernel*, *gamma*, and *C*:

- kernel: rbf
- gamma: scale, auto, 1e-08, 1e-07, 1e-06, 1e-05, 1e-04, 1e-03, 3e-08, 3e-07, 3e-06, 3e-05, 3e-04, 3e-03,6e-08, 6e-07, 6e-06, 6e-05, 6e-04, 6e-03
- C: 1, 6, 9, 10, 12, 15, 20, 50

For the kernel parameter, we used the Gaussian kernel or Radial Basis Function (RBF) whose values decrease as the Euclidean distance between any two data points in the input space increases. In this way, a linear combination of a set of RBFs approximates a nonlinear function in the original input feature space. In other words, RBF finds a linear solution in a higher-dimensional space that corresponds to a nonlinear solution in the original space by transforming the data and linearizing the relationships between them [84, 85]. The smoothness of the decision boundary or the distance at which the data points are considered similar (i.e., how quickly the similarity measure decreases with distance) is controlled by the kernel width, *σ*, or *γ*. The RBF kernel is defined as follows:

$$K(x, x') = exp\left(\frac{-|x-x\prime|^2}{2\sigma^2}\right), \quad (10)$$

where $\frac{1}{2\sigma^2} = \gamma$.

In this way, the points that are close to each other will be assigned higher similarity values than those that are farther away. Therefore, $\gamma$ determines the strength of the influence of a single training instance, with a larger $\gamma$ corresponding to a more significant impact from closer data points. In a regression problem, RBF enables the estimation of similarity between a new data point and the SVs [85–88]. *C* is a regularization parameter or a cost factor that controls the balance between the decision boundary (the margin) and the training error to avoid overfitting and reach an optimal trade-off between accuracy and generalizability. Higher values of *C* allow a more robust training performance by minimizing the error and increasing model complexity, but increasing the chance of overfitting, whereas lower values lead to a less complex model with a smoother decision boundary and therefore better generalizability but a higher risk of underfitting [80, 81, 89, 90].

#### Random Forest

Random Forest is an ensemble tree-based learning algorithm based on aggregating classification and regression decision trees, each of which is built independently from a bootstrap sample of training data randomly and recursively drawn from the original dataset (the root node) [91–96]. Random Forest belongs to the family of Classification And Regression Tree (CART) algorithms, albeit it uses two-step randomization to achieve higher accuracy and reduce the risk of overfitting. Unlike CART, at each node of the tree, Random Forest uses a subset of randomly sampled predictor variables determined by the parameter *mtry*. Random feature selection together with bootstrap sampling helps reduce the variance and decorrelate the trees. Further, additional reduction of bias is achieved via growing the trees to maximum depth constrained by a minimum node size that may be no fewer than 1 case compared to CART which builds a single decision tree [97]. Predictions drawn from each decision tree are aggregated and averaged to make a final prediction, smoothing out nonlinearities.

The Splitting rule in Random Forest relies on maximizing the reduction of the impurity introduced by a split, thus ensuring the homogeneity of the daughter nodes. The degree of impurity is measured by the Gini index in classification problems and by the sum of squares in regression problems [96–98]. For the regression problem, Random Forest selects the split that results in the minimal mean squared error for a given node thus minimizing variance within each new node. A decrease in impurity indicates the contribution of each feature to node homogenization with features that are associated with greater impurity reductions are considered more important. To summarize, Random Forest can capture nonlinear relationships and deal with a large number of predictor variables while preserving a high level of robustness and allowing the evaluation of feature importance [94].

The following hyperparameters were tuned during the grid search CV procedure before training the model (*regressor* = *RandomForestRegressor()*): the number of estimators, the maximum depth, and the maximum number of features, as indicated below in the parameter grid:

- n_estimators: 5000
- max_depth: 1, 2, 3, 4, 5, 6
- max_features: sqrt, log2

The number of estimators (*n_estimators*) controls the number of individual decision trees built on different bootstrap samples. The maximum depth (*max_depth*) determines the number of splits that can be made within one tree from the root node to the farthest leaf node thus controlling the length of the path from the top to the bottom node of a given tree. The maximum number of features (*max_features*) refers to the number of predictor variables selected at each split [94, 97].

##### XGBoost

XGBoost (eXtreme Gradient Boosting) belongs to a family of decision tree-based optimization methods. As it follows from its name, XGBoost combines decision trees using the gradient boosting algorithm that relies on the gradient descent method to optimize the loss function, i.e., the error between the real and predicted value of the response variable. The decision trees are trained on the gradient of the loss or error produced by the previous tree. To avoid overfitting, XGBoost additionally incorporates regularization of feature weights [99, 100].

The hyperparameters used to tune the model (*regressor = XGBRegressor()*) included the maximum depth of a decision tree (*max_depth*), the learning rate ($\eta$), the size of the subsample randomly drawn from the training data to grow the tree (*subsample*), and L1 ($\alpha$) and L2 ($\lambda$) regularization parameters:

- booster: gbtree (gradient boosted tree)
- eta: 0.1, 0.2, 0.3
- max_depth: 1, 2, 3, 4, 5, 6
- subsample: 0.3, 0.6, 1
- lambda: 0, 0.5, 1
- alpha: 0, 0.5, 1

#### S7. Commonality analysis

To quantify the contribution of each neuroimaging phenotype and each MRI modality to the relationship between mental health and the *g*-factor, we applied commonality analyses. We performed commonality analyses using predicted values of the *g*-factor based on mental health or MRI data from all outer-fold test sets. For each set of MRI data (i.e., individual neuroimaging phenotypes, neuroimaging phenotypes stacked within each MRI modality, and neuroimaging phenotypes stacked across all MRI modalities), we built three linear regression models (*LinearRegression* function of Python's scikit-learn package): 1) a model including the observed (i.e., derived directly from ESEM) *g*-factor values as response variables and *g*-factor values predicted from mental health as predictor variables, 2) a model including the observed (i.e., derived directly from ESEM) *g*-factor values as response variables and *g*-factor values predicted from a given set of MRI data as predictor variables, and 3) a model including the observed (i.e., derived directly from ESEM) *g*-factor values as response variables and *g*-factor values predicted from mental health and from a given set of MRI data as two independent predictor variables. For each linear regression model, we computed $R^2$ between observed and predicted values of the *g*-factor in unseen participants to estimate how much of the variation in the *g*-factor is captured only by mental health, only by MRI data, and by mental health and MRI data together. We then decomposed $R^2$ into the unique variance, i.e., the variance in the *g*-factor uniquely explained by mental health alone and MRI data alone, and the common variance, i.e., the variance that is captured by mental health and MRI data [101] e.g.,

$$U_{\mathrm{i}} = R^2_{\mathrm{y.ij}} - R^2_{\mathrm{y.j}}, \quad (11)$$

$$U_{\mathrm{j}} = R^2_{\mathrm{y.ij}} - R^2_{\mathrm{y.i}} \quad (12)$$

$$C_{\mathrm{ij}} = R^2_{\mathrm{y.ij}} - U_{\mathrm{i}} - U_{\mathrm{j}} = R^2_{\mathrm{yj}} + R^2_{\mathrm{y.i}} - R^2_{\mathrm{y.ij}}, \quad (13)$$

where $U_i$ is the variance in the target variable uniquely explained by the first regressor (i.e., the unique contribution of mental health); $U_j$ is the variance in the target variable uniquely explained by the second regressor (i.e., the unique contribution of MRI modalities or neuroimaging phenotypes); $C_{ij}$ is the variance explained by all regressors; $R^2_{\mathrm{y.j}}$, $R^2_{\mathrm{y.i}}$, and $R^2_{\mathrm{y.ij}}$ are the proportions of variance in the target variable attributed to the first, second, and the two predictor variables, respectively, in independent linear regression models [101, 102]. The unique contribution of each predictor variable in commonality analysis is defined as the squared semipartial correlation between the predictor and the dependent variables after removing the effects of other predictor variables.

The total effect of mental health included the unique effect of mental health and the effect shared with MRI data. By computing the relation of the effect of mental health shared with MRI data (i.e., common variance) to the total effect of mental health, we were able to estimate how much of the variance in the *g*-factor that mental health captures is attributed to MRI data. Each commonality analysis was carried out independently and therefore included a different number of participants, depending on how many participants were available for a given neuroimaging phenotype or modality [103].

### Supplementary Results

#### S8. *g*-factor

##### Cognitive scores

**Fig. S1.** Heatmap plot of the correlations between twelve scores from eleven cognitive tests of the UK Biobank cognitive test battery (N = 31 614)

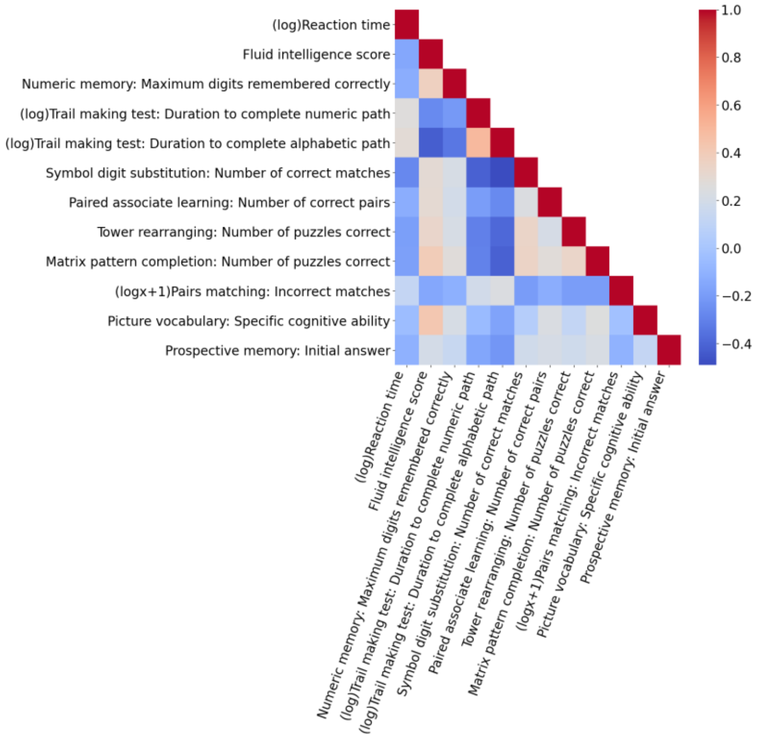

**Table S8.** Correlation matrix (Pearson *r*) for the twelve scores from eleven cognitive tests of the UK Biobank cognitive test battery. All correlations are statistically significant

| | RT | FIS | NM | TMT-num. | TMT-alphab. | SDS | PAL | Tower Rear. | Matrix PC | Pairs Match. | Pict. Vocab. | Prosp. Mem. |
|---|---|---|---|---|---|---|---|---|---|---|---|---|
| **RT** | 1 | | | | | | | | | | | |
| **FIS** | -0.15 | 1 | | | | | | | | | | |
| **NM** | -0.12 | 0.37 | 1 | | | | | | | | | |
| **TMT-num.** | 0.26 | -0.27 | -0.21 | 1 | | | | | | | | |
| **TMT-alphab.** | 0.29 | -0.43 | -0.34 | 0.5 | 1 | | | | | | | |
| **SDS** | -0.27 | 0.29 | 0.22 | -0.42 | -0.49 | 1 | | | | | | |
| **PAL** | -0.12 | 0.3 | 0.19 | -0.19 | -0.27 | 0.24 | 1 | | | | | |
| **Tower Rear.** | -0.19 | 0.33 | 0.22 | -0.3 | -0.39 | 0.34 | 0.21 | 1 | | | | |
| **Matrix PC** | -0.19 | 0.4 | 0.27 | -0.3 | -0.41 | 0.35 | 0.28 | 0.33 | 1 | | | |
| **Pairs Match.** | 0.13 | -0.16 | -0.11 | 0.19 | 0.24 | -0.2 | -0.13 | -0.2 | -0.19 | 1 | | |
| **Pict. Vocab.** | -0.05 | 0.43 | 0.22 | -0.06 | -0.17 | 0.05 | 0.23 | 0.12 | 0.25 | -0.03 | 1 | |
| **Prosp. Mem.** | -0.11 | 0.2 | 0.14 | -0.16 | -0.22 | 0.18 | 0.21 | 0.18 | 0.22 | -0.1 | 0.12 | 1 |

*RT* ($\log_x$)Reaction time, *FIS* Fluid intelligence score, *NM* Numeric memory, *TMT-num.* ($\log_x$)Trail Making Test: Duration to complete numeric path, *TMT-alphab.* ($\log_x$)Trail Making Test: Duration to complete alphabetic path, *SDS* Symbol digit substitution: Number of correct matches, *PAL* Paired associate learning: Number of correct pairs, *Tower Rear.* Tower rearranging: Number of puzzles correct, *Matrix PC* Matrix pattern completion: Number of puzzles correct, *Pairs Match.* ($\log_{x+1}$)Pairs matching: Incorrect matches, *Pict. Vocab.* Picture vocabulary: Specific cognitive ability, *Prosp. Mem.* Prospective memory: Initial answer.

#### *g*-factor modeling

In each fold, Kaiser-Meyer-Olkin statistics (KMO > 0.87) and Bartlett's sphericity test ($p < 0.05$) suggested good factorability of the data. Parallel factor analysis suggested that four factors are sufficient to explain the latent structure of the dataset. For all four factors, correlations of the regression-based factor scores with factors were > 0.7 indicating that the factor scores are good representations (predictors) of the underlying cognitive domains measured by the cognitive tests. Construct validity of the factor structure of the dataset was confirmed by ESEM/CFA results. For each of the five folds, the CFI was > 0.96, the TLI > 0.92, the RMSEA ≤ 0.05, and the SRMR < 0.03, indicating a good fit [8, 104–107]. Four latent factors captured around 27% of the covariance structure of cognitive tests.

**Table S9.** Loadings of the cognitive scores onto four latent factors in five folds

| Cognitive score | Factor 1 | Factor 2 | Factor 3 | Factor 4 |
|---|---|---|---|---|
| | | **Fold 1** | | |
| ($log_x$)Reaction time | 0.414 | | | |
| Fluid intelligence score | | 0.325 | 0.565 | |
| Numeric memory: Maximum digits remembered correctly | -0.167 | 0.145 | 0.312 | |
| ($log_x$)Trail making test: Duration to complete numeric path | 0.709 | | | |
| ($log_x$)Trail making test: Duration to complete alphabetic path | 0.651 | | -0.162 | |
| Symbol digit substitution: Number of correct matches | -0.510 | 0.154 | | |
| Paired associate learning: Number of correct pairs | | | | 0.438 |
| Tower rearranging: Number of puzzles correct | | 0.573 | | |
| Matrix pattern completion: Number of puzzles correct | | 0.410 | 0.129 | 0.163 |
| ($log_{x+1}$)Pairs matching: Incorrect matches | | -0.318 | | |
| Picture vocabulary: Specific cognitive ability | | | 0.553 | 0.213 |
| Prospective memory: Initial answer | | 0.166 | | 0.269 |
| **SS loadings** | 1.4 | 0.798 | 0.783 | 0.347 |
| **Proportion Var** | 0.117 | 0.066 | 0.065 | 0.029 |
| **Cumulative Var** | 0.117 | 0.183 | 0.248 | 0.277 |

| | | | | |
|---|---|---|---|---|
| | | **Fold 2** | | |
| ($log_x$)Reaction time | 0.384 | | | |
| Fluid intelligence score | | 0.447 | 0.502 | |
| Numeric memory: Maximum digits remembered correctly | -0.143 | 0.221 | 0.263 | |
| ($log_x$)Trail making test: Duration to complete numeric path | 0.709 | | | |
| ($log_x$)Trail making test: Duration to complete alphabetic path | 0.610 | -0.177 | -0.124 | |
| Symbol digit substitution: Number of correct matches | -0.472 | 0.174 | -0.113 | |
| Paired associate learning: Number of correct pairs | | | | 0.455 |
| Tower rearranging: Number of puzzles correct | | 0.556 | | |
| Matrix pattern completion: Number of puzzles correct | | 0.404 | | 0.203 |
| ($log_{x+1}$)Pairs matching: Incorrect matches | | -0.321 | | |
| Picture vocabulary: Specific cognitive ability | | | 0.523 | 0.249 |
| Prospective memory: Initial answer | | | | 0.307 |
| **SS loadings** | 1.273 | 0.899 | 0.645 | 0.418 |
| **Proportion Var** | 0.106 | 0.075 | 0.054 | 0.035 |
| **Cumulative Var** | 0.106 | 0.181 | 0.235 | 0.27 |
| | | **Fold 3** | | |
| ($log_x$)Reaction time | 0.402 | | | |
| Fluid intelligence score | | 0.427 | 0.513 | |
| Numeric memory: Maximum digits remembered correctly | -0.118 | 0.243 | 0.265 | |
| ($log_x$)Trail making test: Duration to complete numeric path | 0.688 | | | |

| | | | | |
|---|---|---|---|---|
| ($log_x$)Trail making test: Duration to complete alphabetic path | 0.575 | -0.215 | -0.113 | |
| Symbol digit substitution: Number of correct matches | -0.463 | 0.203 | -0.103 | |
| Paired associate learning: Number of correct pairs | | | | 0.446 |
| Tower rearranging: Number of puzzles correct | | 0.573 | | |
| Matrix pattern completion: Number of puzzles correct | | 0.438 | | 0.175 |
| ($log_{x+1}$)Pairs matching: Incorrect matches | | -0.308 | | |
| Picture vocabulary: Specific cognitive ability | | | 0.541 | 0.232 |
| Prospective memory: Initial answer | | 0.165 | | 0.291 |
| **SS loadings** | 1.206 | 0.974 | 0.673 | 0.378 |
| **Proportion Var** | 0.101 | 0.081 | 0.056 | 0.031 |
| **Cumulative Var** | 0.101 | 0.182 | 0.238 | 0.269 |

**Fold 4**

| | | | | |
|---|---|---|---|---|
| ($log_x$)Reaction time | | 0.384 | | |
| Fluid intelligence score | 0.760 | | | |
| Numeric memory: Maximum digits remembered correctly | 0.408 | -0.134 | | |
| ($log_x$)Trail making test: Duration to complete numeric path | | 0.685 | | |
| ($log_x$)Trail making test: Duration to complete alphabetic path | -0.243 | 0.612 | | |
| Symbol digit substitution: Number of correct matches | | -0.491 | 0.136 | 0.135 |
| Paired associate learning: Number of correct pairs | | | 0.454 | |
| Tower rearranging: Number of puzzles correct | 0.199 | | | 0.416 |
| Matrix pattern completion: Number of puzzles correct | 0.282 | | 0.216 | 0.209 |

| | | | | |
|---|---|---|---|---|
| $(\log_{x+1})$Pairs matching: Incorrect matches | | | | -0.272 |
| Picture vocabulary: Specific cognitive ability | 0.601 | | 0.145 | -0.309 |
| Prospective memory: Initial answer | | | 0.321 | |
| **SS loadings** | 1.295 | 1.260 | 0.416 | 0.420 |
| **Proportion Var** | 0.108 | 0.105 | 0.035 | 0.035 |
| **Cumulative Var** | 0.108 | 0.213 | 0.248 | 0.283 |
| | | **Fold 5** | | |
| $(\log_{x})$Reaction time | 0.392 | | | |
| Fluid intelligence score | | 0.322 | 0.589 | |
| Numeric memory: Maximum digits remembered correctly | -0.174 | 0.132 | 0.308 | |
| $(\log_{x})$Trail making test: Duration to complete numeric path | 0.712 | | | |
| $(\log_{x})$Trail making test: Duration to complete alphabetic path | 0.672 | | -0.171 | |
| Symbol digit substitution: Number of correct matches | -0.491 | 0.180 | | |
| Paired associate learning: Number of correct pairs | | | | 0.429 |
| Tower rearranging: Number of puzzles correct | | 0.573 | | |
| Matrix pattern completion: Number of puzzles correct | | 0.374 | 0.143 | 0.172 |
| $(\log_{x+1})$Pairs matching: Incorrect matches | | -0.301 | | |
| Picture vocabulary: Specific cognitive ability | | 0.535 | 0.243 | |
| Prospective memory: Initial answer | | 0.170 | | 0.283 |
| **SS loadings** | 1.402 | 0.762 | 0.794 | 0.362 |
| **Proportion Var** | 0.117 | 0.063 | 0.066 | 0.030 |
| **Cumulative Var** | 0.117 | 0.180 | 0.246 | 0.277 |

*SS* sum of squares, *Var* variance.

**Table S10.** Goodness-of-fit indices for the hierarchical model in five folds

| | $\chi^2$ | ***p*-value for $\chi^2$** | **df** | **CFI** | **TLI** | **BIC** | **RMSEA** | **SRMR** |
|---|---|---|---|---|---|---|---|---|
| **Fold 1** | 1812.236 | <0.001 | 30 | 0.969 | 0.933 | 805169.905 | 0.048 | 0.026 |
| **Fold 2** | 892.577 | <0.001 | 30 | 0.985 | 0.967 | 804456.322 | 0.034 | 0.016 |
| **Fold 3** | 805.374 | <0.001 | 30 | 0.987 | 0.971 | 804114.165 | 0.032 | 0.017 |
| **Fold 4** | 1335.887 | <0.001 | 30 | 0.978 | 0.951 | 804537.156 | 0.041 | 0.019 |
| **Fold 5** | 1926.098 | <0.001 | 30 | 0.967 | 0.928 | 805461.947 | 0.050 | 0.027 |

$\chi^2$ Chi-square test statistic, *df* degrees of freedom, *CFI* Comparative Fit Index, *TLI* Tucker-Lewis Index, *BIC* Bayesian Information Criteria, *RMSEA* Root Mean Square Error of Approximation, *SRMR* Standardized Root Mean Square Residual.

The Chi-square ($\chi^2$) test compares the population covariance matrix to the model-implied covariance matrix. $\chi^2$ values that are much higher than the number of degrees of freedom suggest significant differences between the population covariance matrix and the model-implied covariance matrix. At the same time, a significant *p*-value for the Chi-square test rejects the null hypothesis that the differences between elements of the empirical covariance matrix and model-implied covariance matrix are zero. However, because the Chi-square test is sensitive to sample size, in a large sample it can show a significant *p*-value even if the model fit is acceptable [108].

Fit indices are designed to compare the fit of a model of interest and a baseline or independence model. The latter assumes the lack of measurement error in and correlation between the observed variables (i.e., error variances are fixed to zero and factor loadings are fixed to one). A bad baseline model fit is used as a comparison value that identifies the improvement in the target model over the independence model. The TLI, also referred to as the Nonnormed Fit Index, measures relative fit by evaluating the difference between the $\chi^2$ of the baseline model and the target model. Unlike the Normed Fit Index, TLI is not affected by the sample size as it considers the number of degrees of freedom in both models [109]. TLI values > 0.90 indicate a good model fit relative to the baseline model. The CFI is an adjusted version of the Relative Noncentrality Index that overcomes a problem of fit underestimation in small samples. The CFI values > 0.97 are interpreted as a good fit and values > 0.95 are indicative of an acceptable fit. Some research suggests a less conservative cut-off of 0.95 for CFI [8, 109]. Values of the BIC are relative and used to compare different models. Consistency in BIC values across five folds indicates the stability of model performance. RMSA measures approximate or “close” fit instead of the exact fit implied by the Chi-square test. It is computed as the square root of the estimated discrepancy due to approximation per degree of freedom. RMSEA values ≤ 0.05, between 0.05 and 0.08, between 0.08 and 0.10, and > 0.10 can be interpreted as a “close”, an adequate, a mediocre, and a poor fit, respectively. RMSEA values > 0.10 are not acceptable. SRMR is a standardized version of the Root Mean Square Residual (RMR) index which measures the badness-of-fit based on the fitted residuals. SRMR overcomes the problem of the dependency of RMR on the scales of the observed variable (i.e., their variance and covariances). The SRMR values < 0.05 and < 0.1 indicate a good and an acceptable fit, respectively [110].

**Table S11.** The unique variance of each cognitive score that is not explained by the *g*-factor, the variance of each score explained by the *g*-factor, and the proportion of covariance in cognitive scores captured by the *g*-factor

| | **Fold 1: Unique Var** | **Fold 1: Var Exp.** | **Fold 2: Unique Var** | **Fold 2: Var Exp.** | **Fold 3: Unique Var** | **Fold 3: Var Exp.** | **Fold 4: Unique Var** | **Fold 4: Var Exp.** | **Fold 5: Unique Var** | **Fold 5: Var Exp.** |
|---|---|---|---|---|---|---|---|---|---|---|
| **RT** | 0.85 | 0.15 | 0.85 | 0.15 | 0.85 | 0.16 | 0.85 | 0.15 | 0.85 | 0.15 |
| **FIS** | 0.35 | 0.65 | 0.43 | 0.57 | 0.43 | 0.57 | 0.24 | 0.76 | 0.34 | 0.66 |
| **NM** | 0.77 | 0.23 | 0.77 | 0.23 | 0.77 | 0.23 | 0.78 | 0.22 | 0.78 | 0.22 |
| **TMT-num.** | 0.28 | 0.72 | 0.38 | 0.62 | 0.43 | 0.57 | 0.26 | 0.74 | 0.24 | 0.76 |
| **TMT-alphab.** | 0.46 | 0.54 | 0.44 | 0.56 | 0.42 | 0.58 | 0.45 | 0.55 | 0.46 | 0.54 |
| **SDS** | 0.58 | 0.42 | 0.55 | 0.45 | 0.56 | 0.44 | 0.57 | 0.43 | 0.58 | 0.43 |
| **PAL** | 0.65 | 0.35 | 0.74 | 0.27 | 0.74 | 0.26 | 0.65 | 0.35 | 0.67 | 0.33 |
| **Tower Rear.** | 0.55 | 0.45 | 0.02 | 0.98 | -0.07 | 1.07 | 0.62 | 0.39 | 0.56 | 0.44 |
| **Matrix PC** | 0.64 | 0.37 | 0.64 | 0.36 | 0.64 | 0.37 | 0.64 | 0.36 | 0.63 | 0.37 |

| | | | | | | | | | | |
|---|---|---|---|---|---|---|---|---|---|---|
| **Pairs Match.** | 0.89 | 0.11 | 0.89 | 0.11 | 0.90 | 0.11 | 0.89 | 0.11 | 0.89 | 0.11 |
| **Pict. Vocab.** | 0.62 | 0.38 | 0.62 | 0.38 | 0.62 | 0.38 | 0.67 | 0.33 | 0.62 | 0.38 |
| **Prosp. Mem.** | 0.86 | 0.14 | 0.88 | 0.12 | 0.86 | 0.14 | 0.86 | 0.14 | 0.86 | 0.14 |
| **% Var Exp.** | | 38% | | 40% | | 40% | | 38% | | 38% |

*Unique Var* the standardized residual variances (the unexplained or unique variance of each cognitive score), *Var Exp.* the proportion of variance in each cognitive score explained by the *g*-factor (standardized loadings), *% Var Exp.* the proportion of variance in the cognitive scores captured by the *g*-factor (∑Var Exp. ÷ N scores), *RT* ($log_x$)Reaction time, *FIS* Fluid intelligence score, *NM* Numeric memory, *TMT-num.* ($log_x$)Trail Making Test: Duration to complete numeric path, *TMT-alphab.* ($log_x$)Trail Making Test: Duration to complete alphabetic path, *SDS* Symbol digit substitution: Number of correct matches, *PAL* Paired associate learning: Number of correct pairs, *Tower Rear.* Tower rearranging: Number of puzzles correct, *Matrix PC* Matrix pattern completion: Number of puzzles correct, *Pairs Match.* ($log_{x+1}$)Pairs matching: Incorrect matches, *Pict. Vocab.* Picture vocabulary: Specific cognitive ability, *Prosp. Mem.* Prospective memory: Initial answer.

### S9. Mental Health

#### Partial Least Squares Regression (PLSR)

**Table S12.** Out-of-sample predictive performance of mental health features in the PLSR model in five folds

| | n components | MSE | MAE | $R^2$ | *r* | *p*-value |
|---|---|---|---|---|---|---|
| **Fold 1** | 124 | 0.425 | 0.521 | 0.142 | 0.377 | 0.0 |
| **Fold 2** | 15 | 0.621 | 0.624 | 0.061 | 0.25 | 0.0 |
| **Fold 3** | 15 | 0.696 | 0.667 | 0.059 | 0.244 | 0.0 |
| **Fold 4** | 25 | 0.448 | 0.531 | 0.12 | 0.347 | 0.0 |
| **Fold 5** | 25 | 0.448 | 0.527 | 0.114 | 0.34 | 0.0 |

| | | | | |
|---|---|---|---|---|
| **Mean performance:** | **0.53** | **0.57** | **0.10** | **0.31** |

*MSE* mean squared error, *MAE* mean absolute error, *r*, Pearson *r*.

### S10. Neuroimaging

#### Partial Least Squares Regression (PLSR)

**Table S13.** Out-of-sample predictive performance of dwMRI in the PLSR model averaged across five folds

| dwMRI neuroimaging phenotypes | $R^2$ | Pearson *r* | MSE | MAE |
|---|---|---|---|---|
| **aparc MSA I Connectome Streamline Count** | 0.052 | 0.226 | 0.569 | 0.596 |
| **aparc MSA I Connectome SIFT2** | 0.051 | 0.224 | 0.569 | 0.596 |
| **aparc a2009s MSA I Connectome Streamline Count** | 0.05 | 0.227 | 0.57 | 0.597 |
| **Schaefer7n200p MSA I Connectome Streamline Count** | 0.049 | 0.221 | 0.57 | 0.597 |
| **Schaefer7n200p MSA I Connectome SIFT2** | 0.048 | 0.221 | 0.571 | 0.598 |
| **aparc MSA I Connectome FA** | 0.048 | 0.214 | 0.571 | 0.597 |
| **aparc a2009s MSA I Connectome FA** | 0.047 | 0.213 | 0.572 | 0.597 |
| **aparc MSA I Connectome Mean Length** | 0.047 | 0.215 | 0.571 | 0.597 |
| **aparc a2009s MSA I Connectome SIFT2** | 0.046 | 0.215 | 0.572 | 0.598 |
| **L2 TBSS** | 0.045 | 0.207 | 0.576 | 0.601 |
| **Schaefer7n200p MSA I Connectome FA** | 0.045 | 0.21 | 0.573 | 0.598 |
| **aparc a2009s MSA I Connectome Mean** | 0.043 | 0.207 | 0.574 | 0.599 |

| | | | | |
|---|---|---|---|---|
| **Length** | | | | |
| **Glasser MSA IV Connectome SIFT2** | 0.043 | 0.204 | 0.574 | 0.599 |
| **Glasser MSA IV Connectome Streamline Count** | 0.043 | 0.202 | 0.574 | 0.599 |
| **L3 TBSS** | 0.042 | 0.2 | 0.578 | 0.602 |
| **ICVF TBSS** | 0.042 | 0.199 | 0.578 | 0.602 |
| **Glasser MSA IV Connectome FA** | 0.042 | 0.195 | 0.574 | 0.599 |
| **Schaefer7n500p MSA IV Connectome SIFT2** | 0.041 | 0.204 | 0.575 | 0.599 |
| **L1 TBSS** | 0.041 | 0.197 | 0.579 | 0.603 |
| **Schaefer7n500p MSA IV Connectome Streamline Count** | 0.041 | 0.202 | 0.575 | 0.599 |
| **FA TBSS** | 0.041 | 0.198 | 0.579 | 0.602 |
| **Glasser MSA I Connectome FA** | 0.04 | 0.191 | 0.575 | 0.6 |
| **Schaefer7n200p MSA I Connectome Mean Length** | 0.039 | 0.196 | 0.576 | 0.6 |
| **MD TBSS** | 0.039 | 0.194 | 0.58 | 0.603 |
| **MD Probabilistic** | 0.039 | 0.19 | 0.58 | 0.603 |
| **Glasser MSA I Connectome Streamline Count** | 0.037 | 0.191 | 0.577 | 0.6 |
| **L3 Probabilistic** | 0.037 | 0.186 | 0.582 | 0.604 |
| **Schaefer7n500p MSA IV Connectome FA** | 0.037 | 0.19 | 0.577 | 0.6 |
| **Glasser MSA I Connectome SIFT2** | 0.037 | 0.192 | 0.577 | 0.6 |
| **L2 Probabilistic** | 0.035 | 0.181 | 0.582 | 0.604 |
| **L1 Probabilistic** | 0.033 | 0.178 | 0.583 | 0.605 |
| **ISOVF TBSS** | 0.032 | 0.175 | 0.584 | 0.605 |
| **Glasser MSA IV Connectome Mean Length** | 0.031 | 0.181 | 0.58 | 0.602 |
| **Glasser MSA I** | 0.03 | 0.178 | 0.581 | 0.602 |

| | | | | |
|---|---|---|---|---|
| **Connectome Mean Length** | | | | |
| **OD TBSS** | 0.029 | 0.165 | 0.586 | 0.606 |
| **ICVF Probabilistic** | 0.029 | 0.165 | 0.586 | 0.606 |
| **MO TBSS** | 0.028 | 0.163 | 0.586 | 0.606 |
| **ISOVF Probabilistic** | 0.023 | 0.15 | 0.588 | 0.608 |
| **FA Probabilistic** | 0.022 | 0.141 | 0.59 | 0.609 |
| **Schaefer7n500p MSA IV Connectome Mean Length** | 0.018 | 0.145 | 0.587 | 0.606 |
| **OD Probabilistic** | 0.01 | 0.099 | 0.596 | 0.612 |
| **MO Probabilistic** | 0.006 | 0.076 | 0.598 | 0.613 |

*dwMRI* diffusion-weighted MRI, $R^2$ coefficient of determination, *MSE* mean squared error, *MAE* mean absolute error, *FA* fractional anisotropy, *MD* mean diffusivity, *MO* diffusion tensor mode, *L1, L2, L3* eigenvalues of the diffusion tensor, *OD* orientation dispersion index, *ICVF* intracellular volume fraction, *ISOVF* isotropic volume fraction, *TBSS* Tract-Based Spatial Statistics, *aparc* Desikan-Killiany cortical atlas, *aparc a2009s* Destrieux cortical atlas, *Schaefer7n200p* Schaefer Atlas for 7 networks and 200 parcels, *Schaefer7n500p* Schaefer Atlas for 7 networks and 500 parcels, *MSA* Melbourne Subcortical Atlas, *SIFT2* Spherical-Deconvolution Informed Filtering of Tractograms 2.

**Table S14.** Out-of-sample predictive performance of rsMRI in the PLSR model averaged across five folds

| rsMRI neuroimaging phenotypes | $R^2$ | Pearson $r$ | MSE | MAE |
|---|---|---|---|---|
| **55 IC Functional connectivity (Tangent)** | 0.088 | 0.302 | 0.552 | 0.587 |
| **Schaefer7n200p MSA I Functional connectivity (Full correlation)** | 0.071 | 0.271 | 0.557 | 0.589 |
| **Schaefer7n500p MSA IV Functional connectivity (Full correlation)** | 0.058 | 0.252 | 0.563 | 0.593 |
| **Glasser MSA I Functional connectivity (Full correlation)** | 0.057 | 0.249 | 0.564 | 0.593 |

| | | | | |
|---|---|---|---|---|
| **21 IC Functional connectivity** | 0.054 | 0.232 | 0.571 | 0.599 |
| **Glasser MSA IV Functional connectivity (Full correlation)** | 0.053 | 0.243 | 0.566 | 0.595 |
| **aparc MSA I Functional connectivity (Full correlation)** | 0.052 | 0.232 | 0.567 | 0.596 |
| **aparc a2009s MSA I Functional connectivity (Full correlation)** | 0.051 | 0.233 | 0.569 | 0.597 |
| **55 IC Amplitudes** | 0.019 | 0.14 | 0.59 | 0.608 |
| **21 IC Amplitudes** | 0.013 | 0.112 | 0.594 | 0.611 |

*rsMRI* resting-state MRI, $R^2$ coefficient of determination, *MSE* mean squared error, *MAE* mean absolute error, *IC* independent components, *aparc* Desikan-Killiany cortical atlas, *aparc a2009s* Destrieux cortical atlas, *Schaefer7n200p* Schaefer Atlas for 7 networks and 200 parcels, *Schaefer7n500p* Schaefer Atlas for 7 networks and 500 parcels, *MSA* Melbourne Subcortical Atlas.

**Table S15.** Out-of-sample predictive performance of T1w and T2w MRI in the PLSR model averaged across five folds

| sMRI neuroimaging phenotypes | $R^2$ | Pearson *r* | MSE | MAE |
|---|---|---|---|---|
| **Subcortical volumetric subsegmentation** | 0.068 | 0.244 | 0.564 | 0.593 |
| **ASEG volume** | 0.059 | 0.235 | 0.568 | 0.595 |
| **FSL FAST** | 0.051 | 0.219 | 0.572 | 0.598 |
| **Whole-brain T1w/T2w** | 0.047 | 0.209 | 0.567 | 0.596 |
| **ASEG mean intensity** | 0.043 | 0.199 | 0.577 | 0.601 |
| **Desikan grey/white matter intensity** | 0.043 | 0.203 | 0.576 | 0.601 |
| **a2009s volume** | 0.034 | 0.18 | 0.582 | 0.604 |
| **a2009s mean thickness** | 0.027 | 0.161 | 0.586 | 0.606 |
| **Desikan-Killiany-Tourville volume** | 0.025 | 0.157 | 0.586 | 0.606 |

| | | | | |
|---|---|---|---|---|
| **Desikan white matter mean thickness** | 0.024 | 0.151 | 0.587 | 0.607 |
| **Desikan-Killiany-Tourville mean thickness** | 0.024 | 0.152 | 0.587 | 0.606 |
| **a2009s area** | 0.024 | 0.155 | 0.587 | 0.606 |
| **Desikan white matter volume** | 0.023 | 0.152 | 0.587 | 0.607 |
| **FSL FIRST** | 0.023 | 0.146 | 0.588 | 0.607 |
| **BA *ex vivo* volume** | 0.021 | 0.143 | 0.589 | 0.608 |
| **Desikan pial** | 0.021 | 0.144 | 0.589 | 0.608 |
| **Desikan-Killiany-Tourville area** | 0.018 | 0.132 | 0.59 | 0.608 |
| **BA *ex vivo* mean thickness** | 0.018 | 0.132 | 0.591 | 0.608 |
| **Desikan white matter area** | 0.017 | 0.131 | 0.591 | 0.608 |
| **BA *ex vivo* area** | 0.008 | 0.089 | 0.596 | 0.611 |

*sMRI* T1-weighted (T1w) and T2-weighted (T2w) structural MRI, $R^2$ coefficient of determination, *MSE* mean squared error, *MAE* mean absolute error, *ASEG* FreeSurfer automated subcortical volumetric segmentation, *FSL FAST* FSL FMRIB's Automated Segmentation Tool, *a2009s* FreeSurfer Destrieux, *FSL FIRST* FMRIB's Integrated Registration and Segmentation Tool, *BA* FreeSurfer *ex-vivo* Brodmann Area Maps.

#### Feature importance

**Fig. S2.** Feature importance plot for dwMRI neuroimaging phenotype with the highest predictive performance for cognition. Bars display the direction and strength of Pearson correlations ($r \geq 0.2$) between the predicted *g*-factor and white matter streamline counts across each pair of brain regions parcellated using the aparc (Desikan-Killiany) MSA-I atlases. Pearson *r* was computed in outer-fold test sets pooled across five folds.

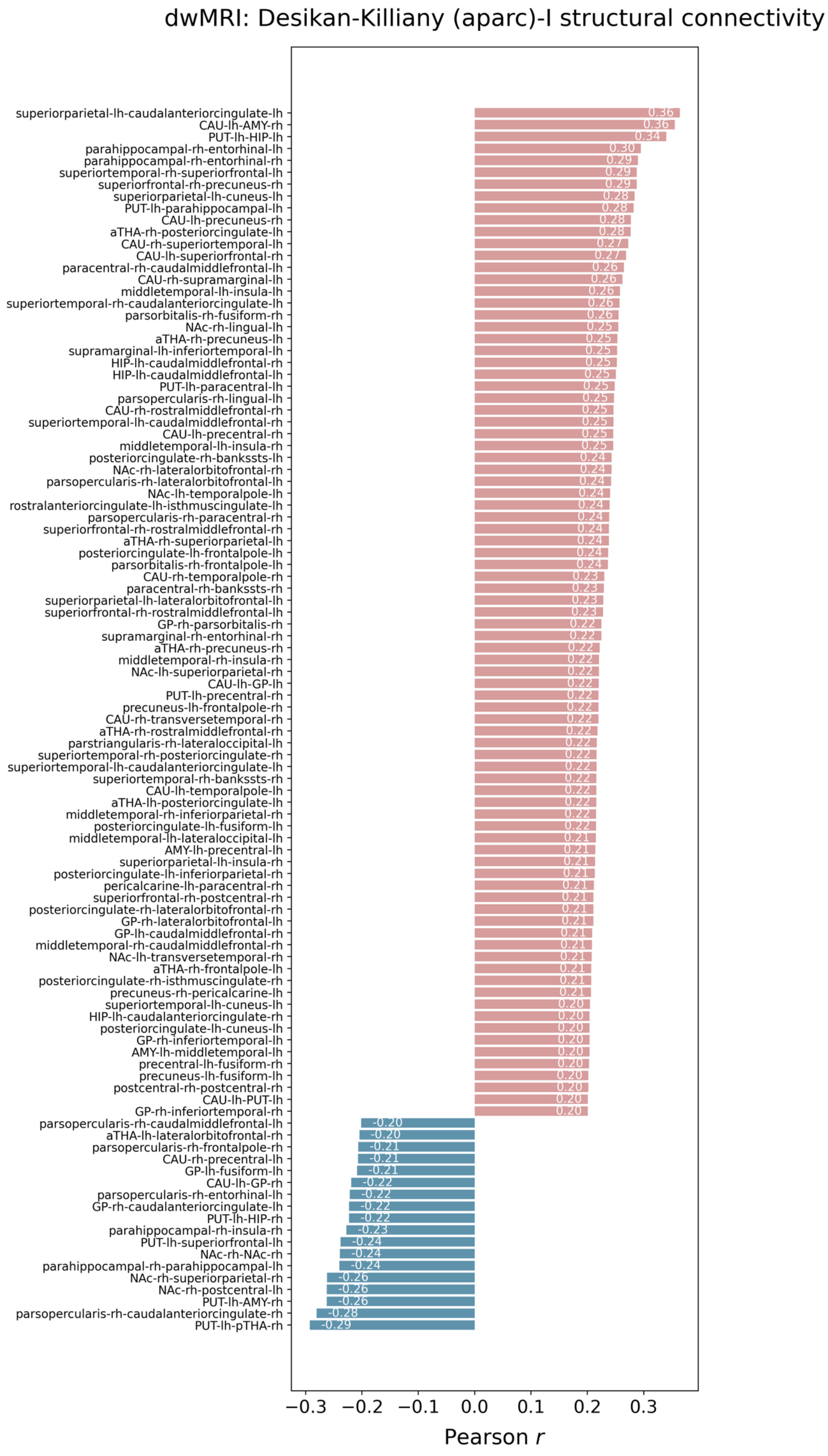
dwMRI: Desikan-Killiany (aparc)-I structural connectivity
superiorparietal-lh-caudalanteriorcingulate-lh 0.36
CAU-lh-AMY-rh 0.36
PUT-lh-HIP-lh 0.34
parahippocampal-rh-entorhinal-lh 0.30
parahippocampal-rh-entorhinal-rh 0.29
superiortemporal-rh-superiorfrontal-lh 0.29
superiorfrontal-rh-precuneus-rh 0.29
superiorparietal-lh-cuneus-lh 0.28
PUT-lh-parahippocampal-lh 0.28
CAU-lh-precuneus-rh 0.28
aTHA-rh-posteriorcingulate-lh 0.28
CAU-rh-superiortemporal-lh 0.27
CAU-lh-superiorfrontal-rh 0.27
paracentral-rh-caudalmiddlefrontal-lh 0.26
CAU-rh-supramarginal-lh 0.26
middletemporal-lh-insula-lh 0.26
superiortemporal-rh-caudalanteriorcingulate-lh 0.26
parsorbitalis-rh-fusiform-rh 0.26
NAc-rh-lingual-lh 0.25
aTHA-rh-precuneus-lh 0.25
supramarginal-lh-inferiortemporal-lh 0.25
HIP-lh-caudalmiddlefrontal-rh 0.25
HIP-lh-caudalmiddlefrontal-lh 0.25
PUT-lh-paracentral-lh 0.25
parsopercularis-rh-lingual-lh 0.25
CAU-rh-rostralmiddlefrontal-rh 0.25
superiortemporal-lh-caudalmiddlefrontal-rh 0.25
CAU-lh-precentral-rh 0.25
middletemporal-lh-insula-rh 0.25
posteriorcingulate-rh-bankssts-lh 0.24
NAc-rh-lateralorbitofrontal-rh 0.24
parsopercularis-rh-lateralorbitofrontal-lh 0.24
NAc-lh-temporalpole-lh 0.24
rostralanteriorcingulate-lh-isthmuscingulate-lh 0.24
parsopercularis-rh-paracentral-rh 0.24
superiorfrontal-rh-rostralmiddlefrontal-rh 0.24
aTHA-rh-superiorparietal-lh 0.24
posteriorcingulate-lh-frontalpole-lh 0.24
parsorbitalis-rh-frontalpole-lh 0.24
CAU-rh-temporalpole-rh 0.23
paracentral-rh-bankssts-rh 0.23
superiorparietal-lh-lateralorbitofrontal-lh 0.23
superiorfrontal-rh-rostralmiddlefrontal-lh 0.23
GP-rh-parsorbitalis-rh 0.22
supramarginal-rh-entorhinal-rh 0.22
aTHA-rh-precuneus-rh 0.22
middletemporal-rh-insula-rh 0.22
NAc-lh-superiorparietal-rh 0.22
CAU-lh-GP-lh 0.22
PUT-lh-precentral-rh 0.22
precuneus-lh-frontalpole-rh 0.22
CAU-rh-transversetemporal-rh 0.22
aTHA-rh-rostralmiddlefrontal-rh 0.22
parstriangularis-rh-lateraloccipital-lh 0.22
superiortemporal-rh-posteriorcingulate-rh 0.22
superiortemporal-lh-caudalanteriorcingulate-lh 0.22
superiortemporal-rh-bankssts-rh 0.22
CAU-lh-temporalpole-lh 0.22
aTHA-lh-posteriorcingulate-lh 0.22
middletemporal-rh-inferiorparietal-rh 0.22
posteriorcingulate-lh-fusiform-lh 0.22
middletemporal-lh-lateraloccipital-lh 0.21
AMY-lh-precentral-lh 0.21
superiorparietal-lh-insula-rh 0.21
posteriorcingulate-lh-inferiorparietal-rh 0.21
pericalcarine-lh-paracentral-rh 0.21
superiorfrontal-rh-postcentral-rh 0.21
posteriorcingulate-rh-lateralorbitofrontal-rh 0.21
GP-rh-lateralorbitofrontal-lh 0.21
GP-lh-caudalmiddlefrontal-rh 0.21
middletemporal-rh-caudalmiddlefrontal-rh 0.21
NAc-lh-transversetemporal-rh 0.21
aTHA-rh-frontalpole-lh 0.21
posteriorcingulate-rh-isthmuscingulate-rh 0.21
precuneus-rh-pericalcarine-lh 0.21
superiortemporal-lh-cuneus-lh 0.20
HIP-lh-caudalanteriorcingulate-rh 0.20
posteriorcingulate-lh-cuneus-lh 0.20
GP-rh-inferiortemporal-lh 0.20
AMY-lh-middletemporal-lh 0.20
precentral-lh-fusiform-rh 0.20
precuneus-lh-fusiform-lh 0.20
postcentral-rh-postcentral-rh 0.20
CAU-lh-PUT-lh 0.20
GP-rh-inferiortemporal-rh 0.20
parsopercularis-rh-caudalmiddlefrontal-lh -0.20
aTHA-lh-lateralorbitofrontal-rh -0.20
parsopercularis-rh-frontalpole-rh -0.21
CAU-rh-precentral-lh -0.21
GP-lh-fusiform-lh -0.21
CAU-lh-GP-rh -0.22
parsopercularis-rh-entorhinal-lh -0.22
GP-rh-caudalanteriorcingulate-lh -0.22
PUT-lh-HIP-rh -0.22
parahippocampal-rh-insula-rh -0.23
PUT-lh-superiorfrontal-lh -0.24
NAc-rh-NAc-rh -0.24
parahippocampal-rh-parahippocampal-lh -0.24
NAc-rh-superiorparietal-rh -0.26
NAc-rh-postcentral-lh -0.26
PUT-lh-AMY-rh -0.26
parsopercularis-rh-caudalanteriorcingulate-rh -0.28
PUT-lh-pTHA-rh -0.29
−0.3 −0.2 −0.1 0.0 0.1 0.2 0.3
Pearson r

**Fig. S3.** Feature importance plot for rsMRI neuroimaging phenotype with the highest predictive performance for cognition. Bars display the direction and strength of Pearson correlations ($r \geq 0.15$) between the predicted *g*-factor and functional connectivity across 55 neuronally driven independent components (IC). Pearson *r* was computed in outer-fold test sets pooled across five folds.

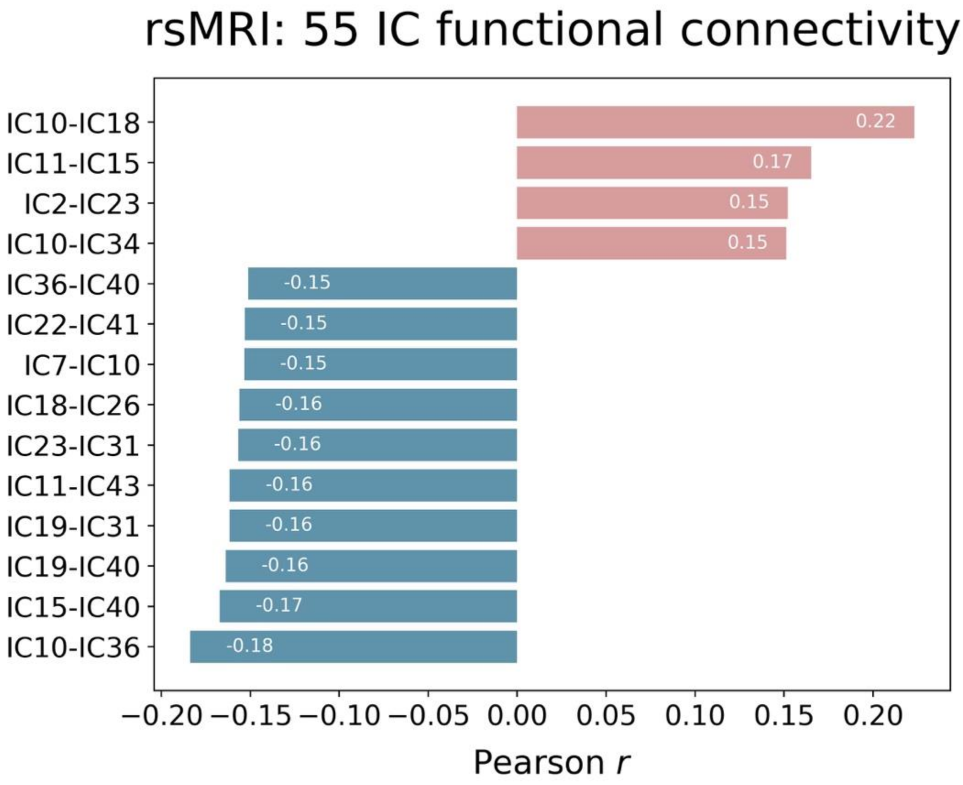

**Fig. S4.** Functional connectivity maps for the 55 neuronally driven independent components (IC) grouped into seven resting-state networks based on Yeo 2011 parcellation [111]. Note IC number in parenthesis corresponds to the original numbers of "good" nodes listed in https://www.fmrib.ox.ac.uk/ukbiobank/.

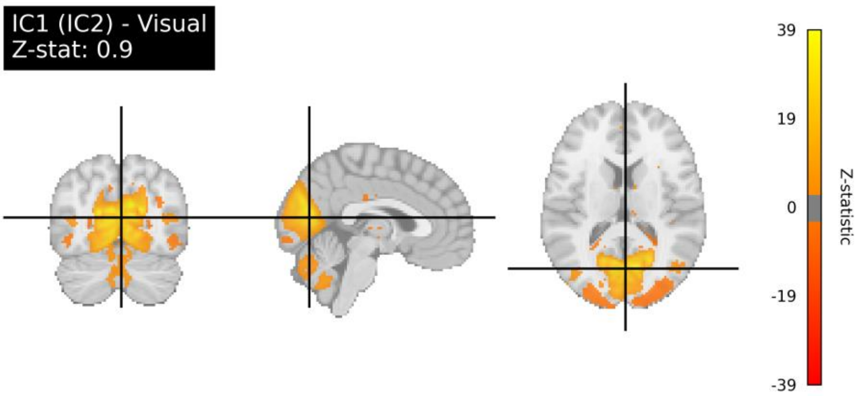

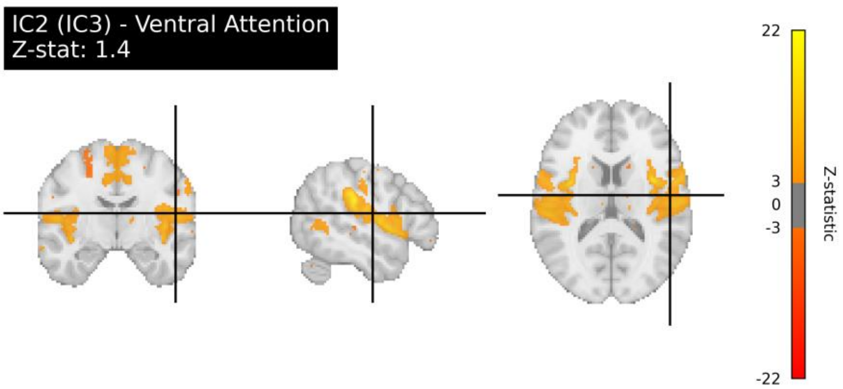
IC2 (IC3) - Ventral Attention
Z-stat: 1.4
22
3
0
-3
-22
Z-statistic

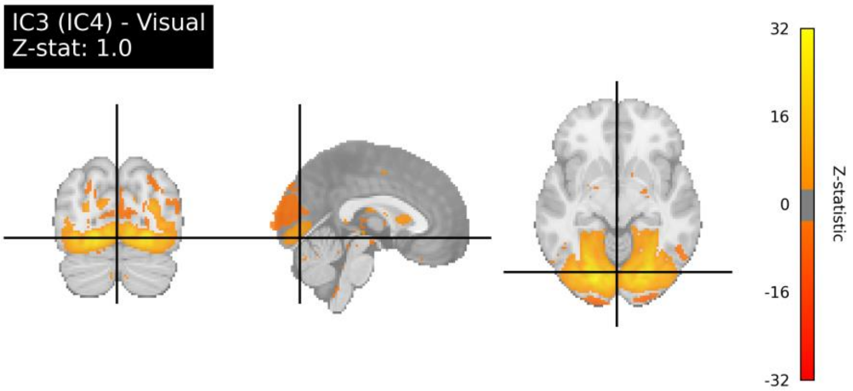
IC3 (IC4) - Visual
Z-stat: 1.0
32
16
0
-16
-32
Z-statistic

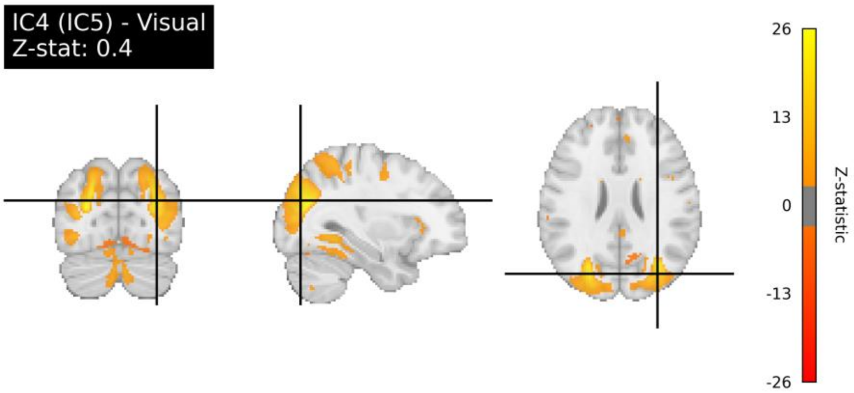
IC4 (IC5) - Visual
Z-stat: 0.4
26
13
0
-13
-26
Z-statistic

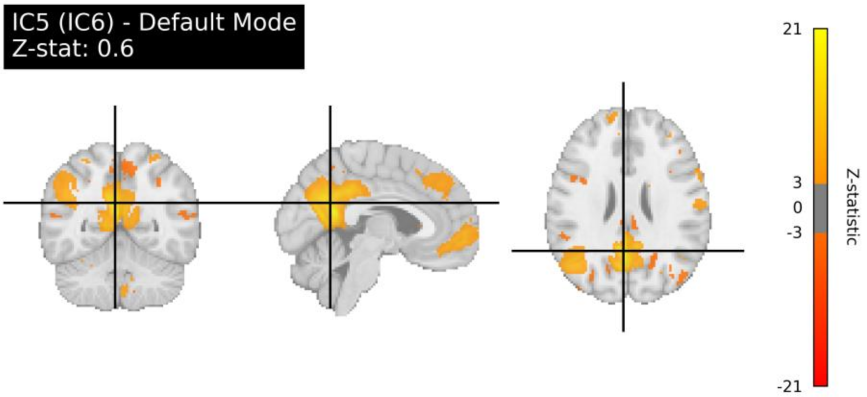
IC5 (IC6) - Default Mode
Z-stat: 0.6
21
3
0
-3
-21
Z-statistic

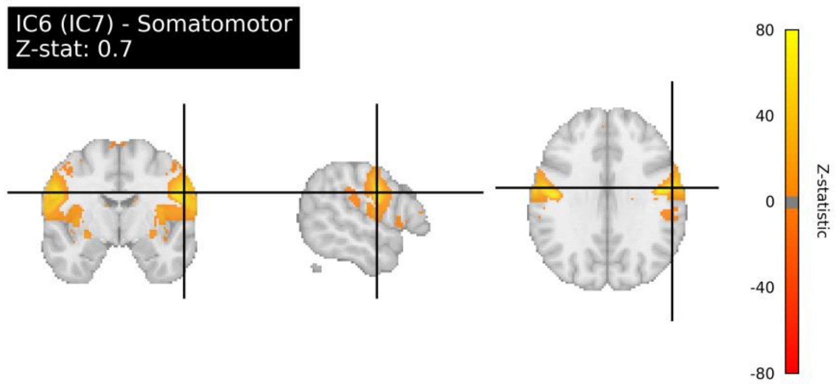
IC6 (IC7) - Somatomotor
Z-stat: 0.7
80
40
0
-40
-80
Z-statistic

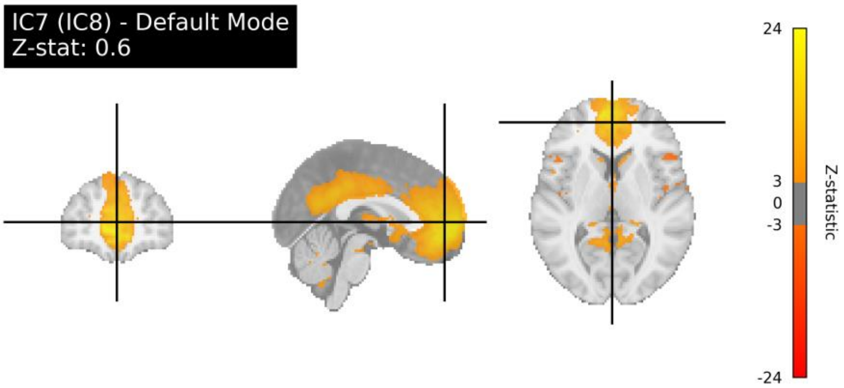
IC7 (IC8) - Default Mode
Z-stat: 0.6
24
3
0
-3
-24
Z-statistic

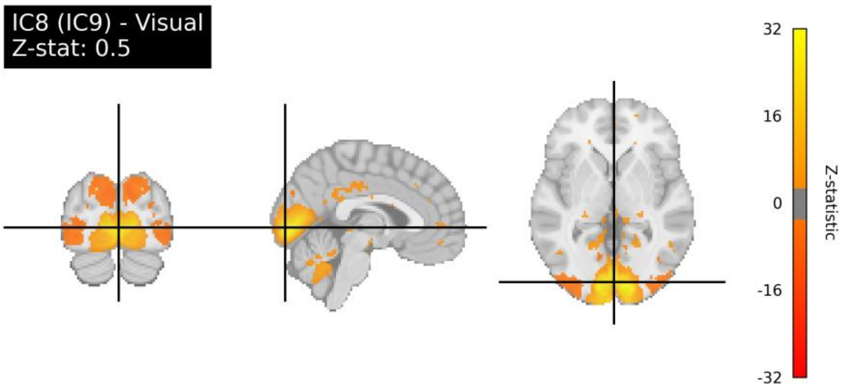
IC8 (IC9) - Visual
Z-stat: 0.5
32
16
0
-16
-32
Z-statistic

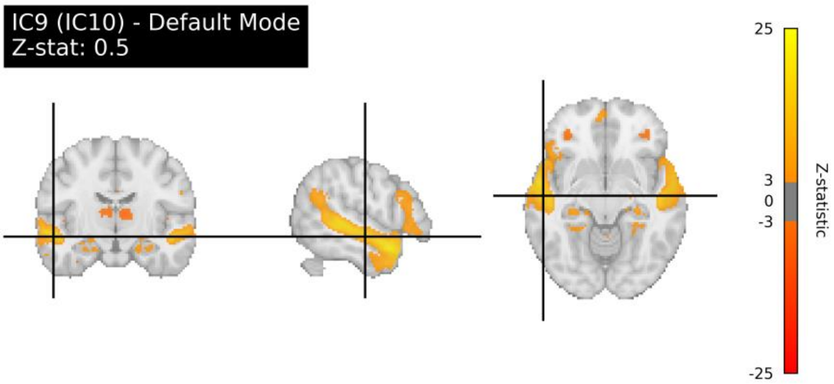
IC9 (IC10) - Default Mode
Z-stat: 0.5
25
3
0
-3
-25
Z-statistic

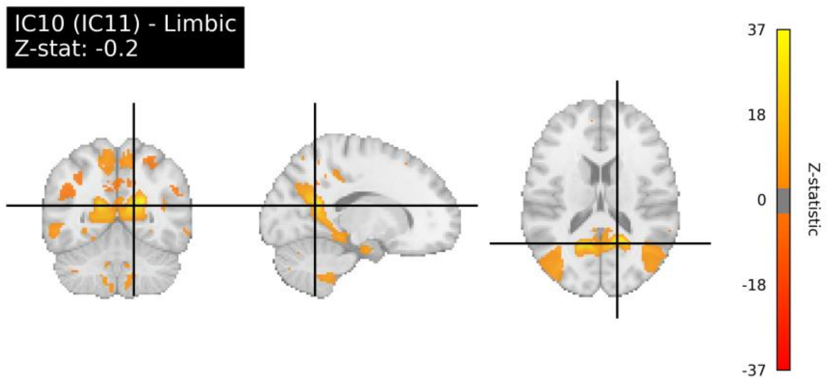
IC10 (IC11) - Limbic
Z-stat: -0.2
37
18
0
-18
-37
Z-statistic

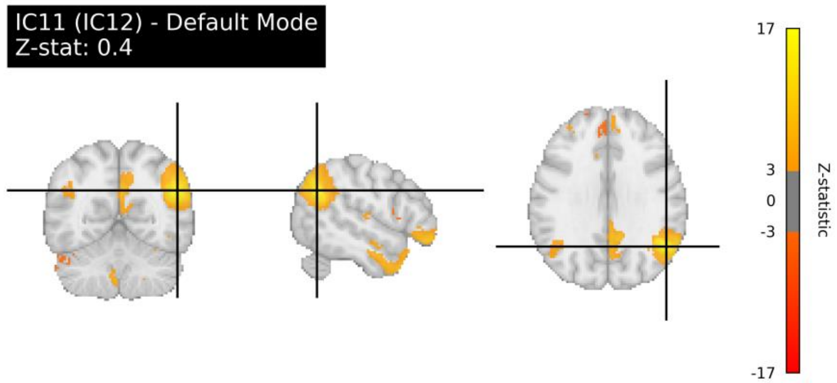
IC11 (IC12) - Default Mode
Z-stat: 0.4
17
3
0
-3
-17
Z-statistic

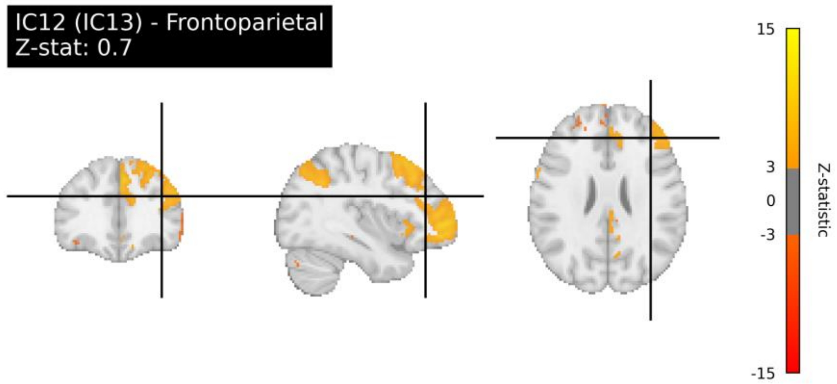
IC12 (IC13) - Frontoparietal
Z-stat: 0.7
15
3
0
-3
-15
Z-statistic

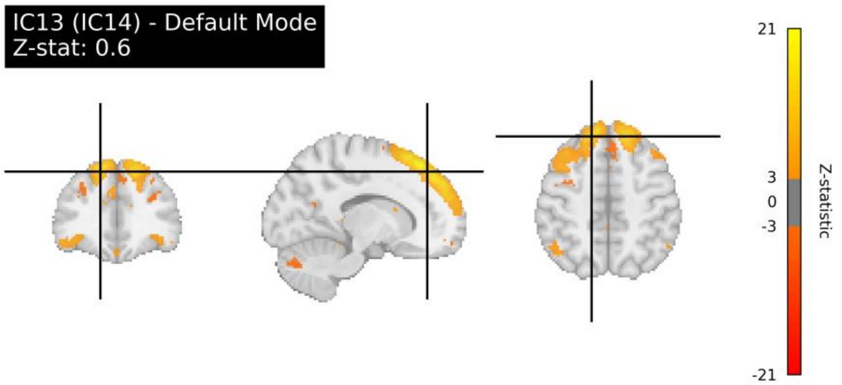
IC13 (IC14) - Default Mode
Z-stat: 0.6
21
3
0
-3
-21
Z-statistic

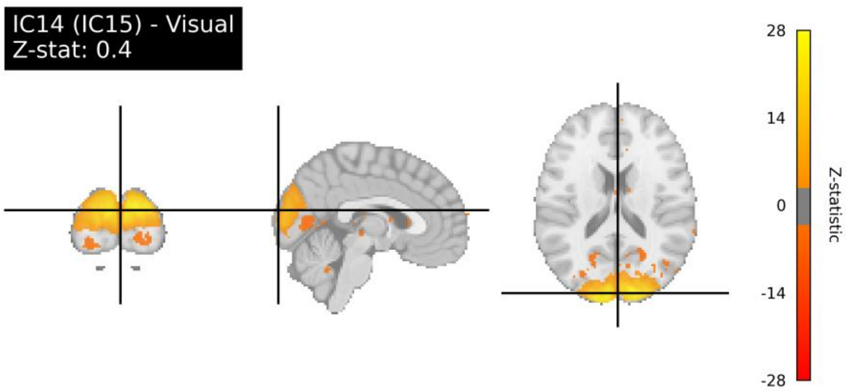
IC14 (IC15) - Visual
Z-stat: 0.4
28
14
0
-14
-28
Z-statistic

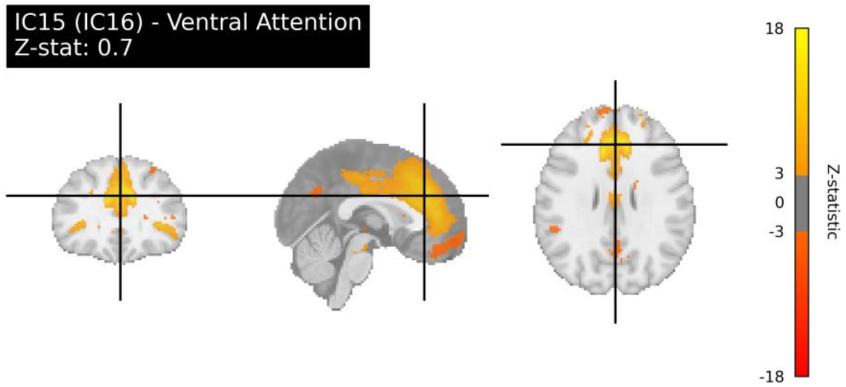
IC15 (IC16) - Ventral Attention
Z-stat: 0.7
18
3
0
-3
-18
Z-statistic

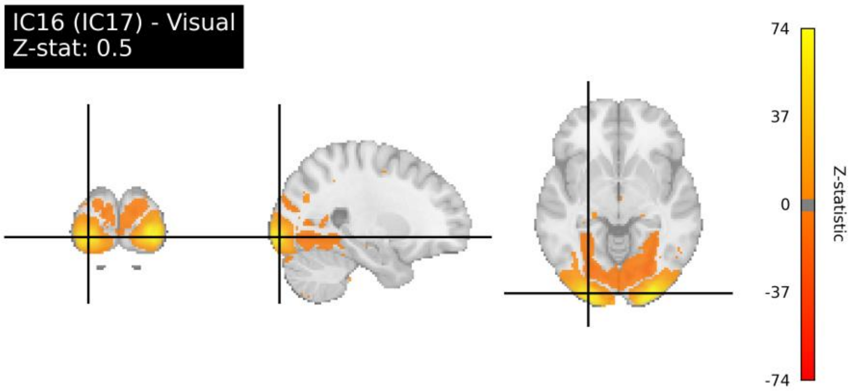
IC16 (IC17) - Visual
Z-stat: 0.5
74
37
0
-37
-74
Z-statistic

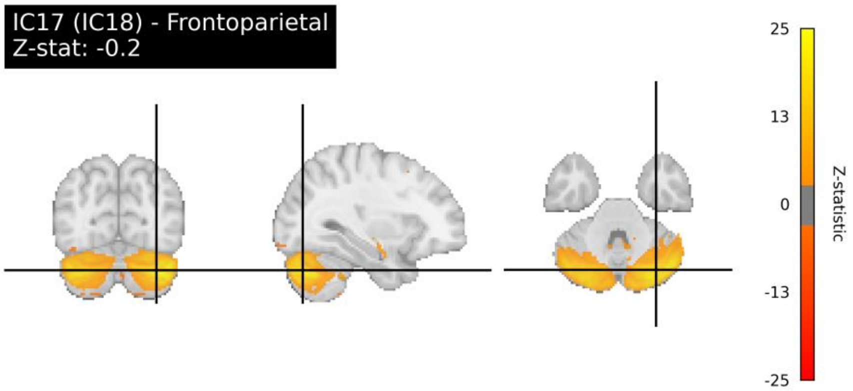
IC17 (IC18) - Frontoparietal
Z-stat: -0.2
25
13
0
-13
-25
Z-statistic

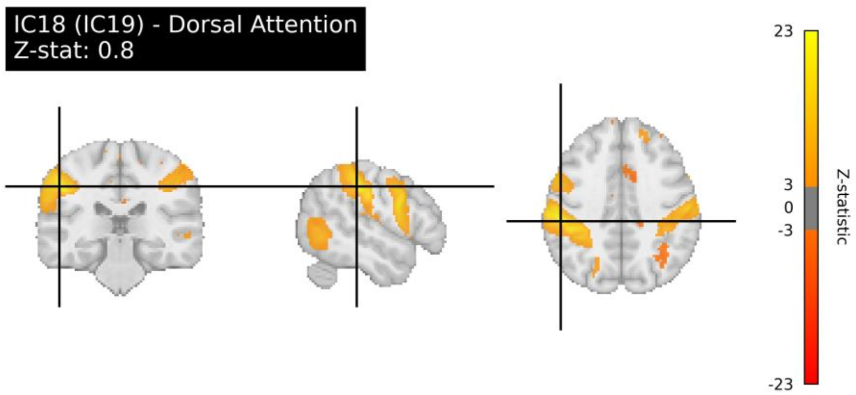
IC18 (IC19) - Dorsal Attention
Z-stat: 0.8
23
3
0
-3
-23
Z-statistic

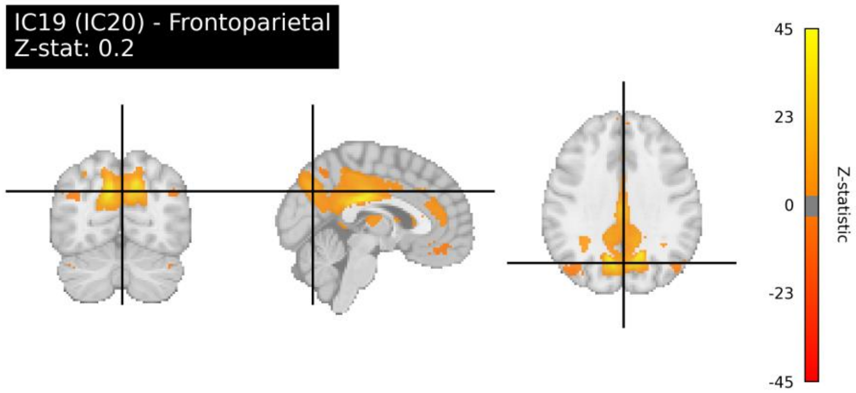
IC19 (IC20) - Frontoparietal
Z-stat: 0.2
45
23
0
-23
-45
Z-statistic

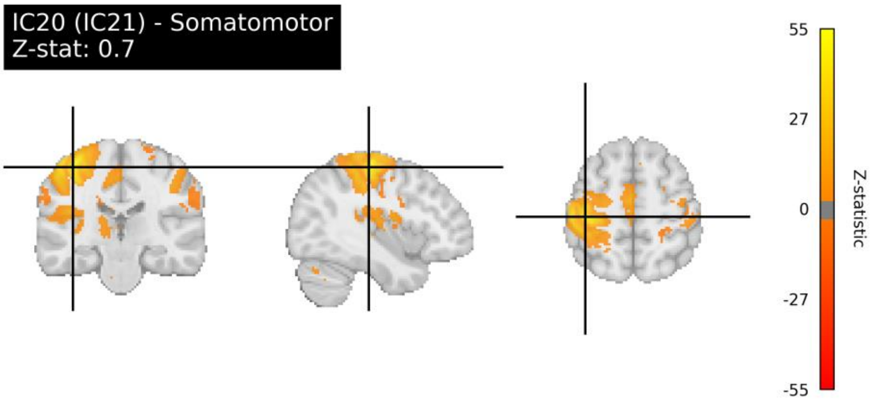
IC20 (IC21) - Somatomotor
Z-stat: 0.7
55
27
0
-27
-55
Z-statistic

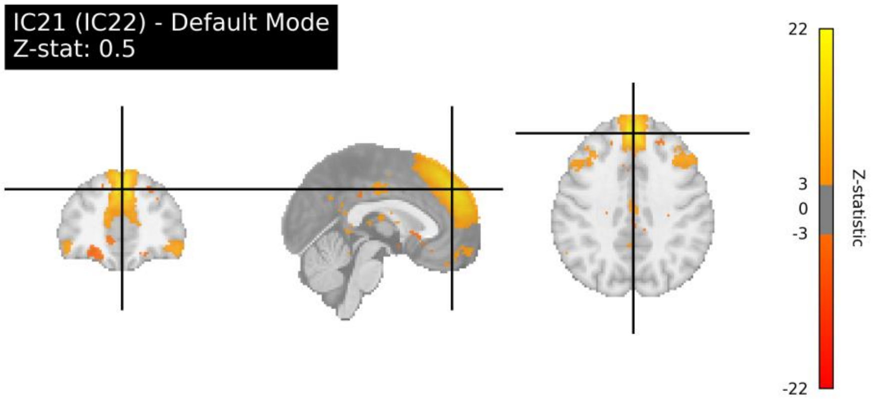
IC21 (IC22) - Default Mode
Z-stat: 0.5
22
3
0
-3
-22
Z-statistic

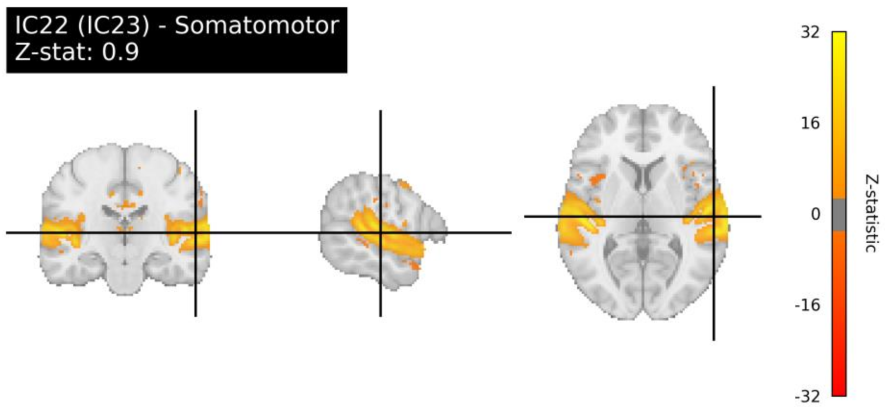
IC22 (IC23) - Somatomotor
Z-stat: 0.9
32
16
0
-16
-32
Z-statistic

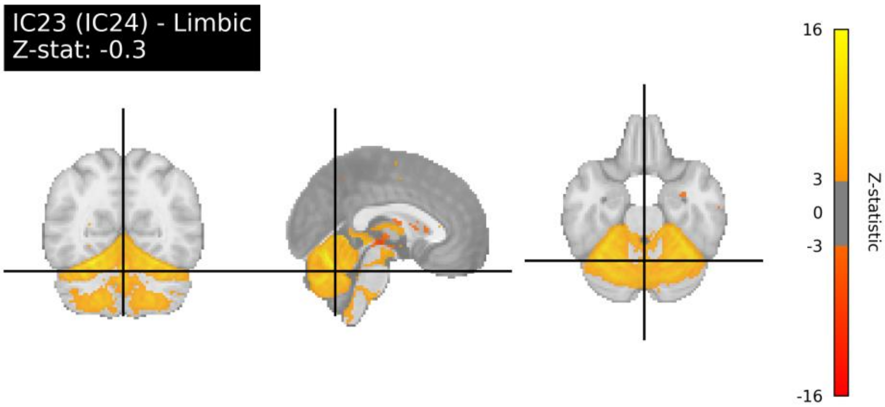
IC23 (IC24) - Limbic
Z-stat: -0.3
16
3
0
-3
-16
Z-statistic

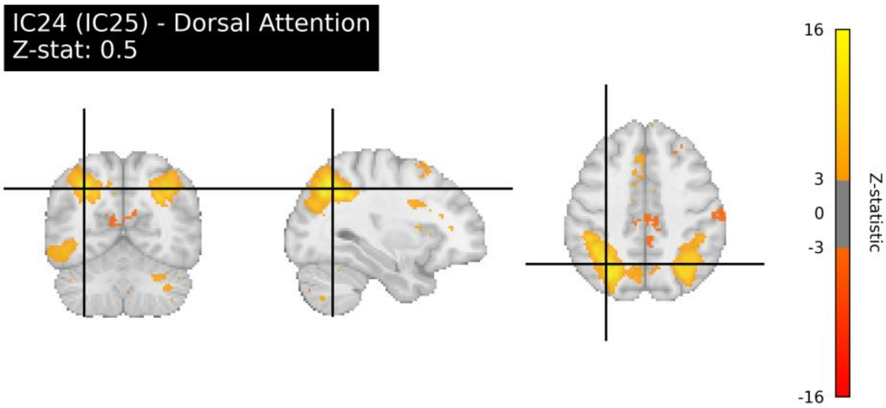
IC24 (IC25) - Dorsal Attention
Z-stat: 0.5
16
3
0
-3
-16
Z-statistic

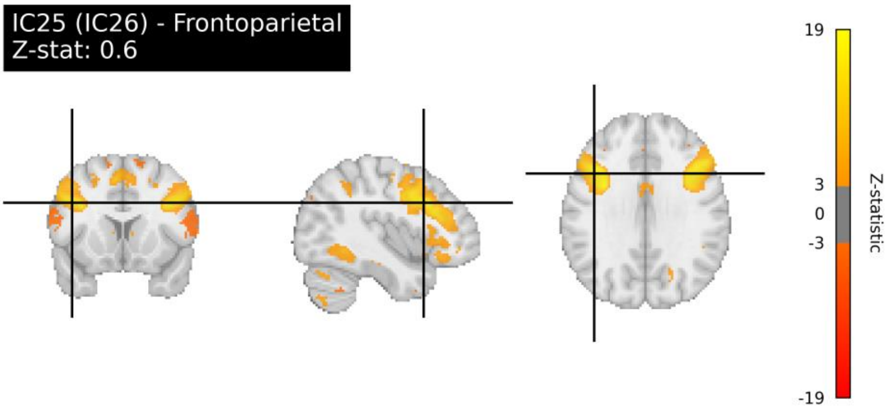
IC25 (IC26) - Frontoparietal
Z-stat: 0.6
19
3
0
-3
-19
Z-statistic

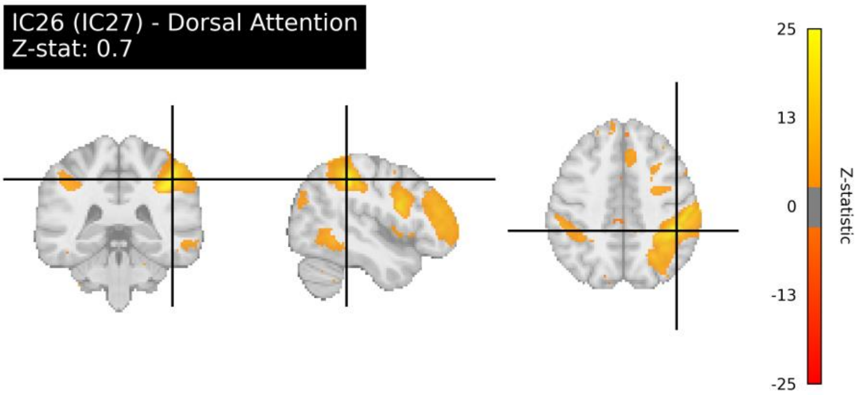
IC26 (IC27) - Dorsal Attention
Z-stat: 0.7
25
13
0
-13
-25
Z-statistic

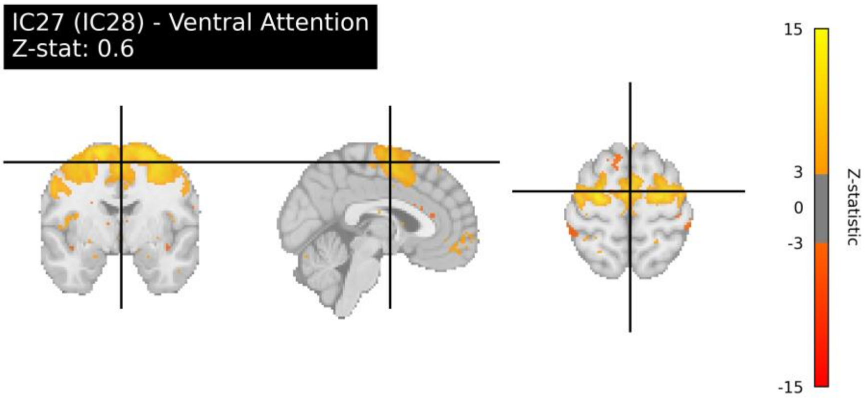
IC27 (IC28) - Ventral Attention
Z-stat: 0.6
15
3
0
-3
-15
Z-statistic

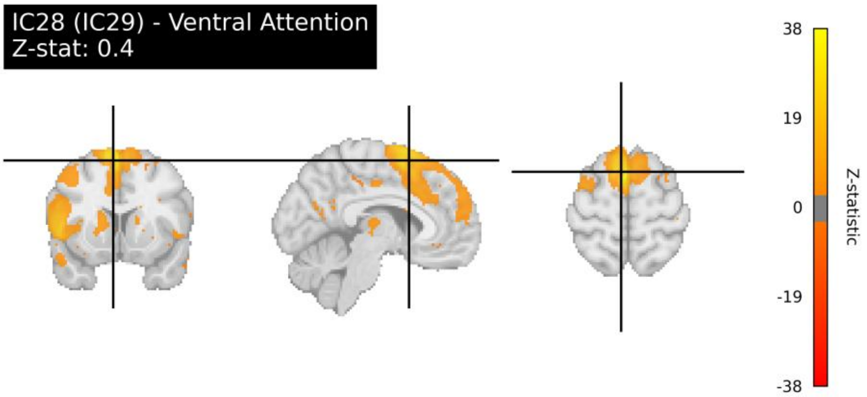
IC28 (IC29) - Ventral Attention
Z-stat: 0.4
38
19
0
-19
-38
Z-statistic

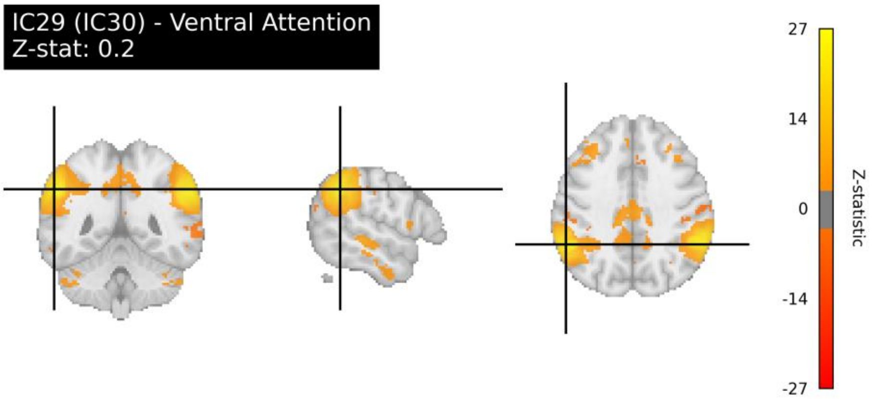
IC29 (IC30) - Ventral Attention
Z-stat: 0.2
27
14
0
-14
-27
Z-statistic

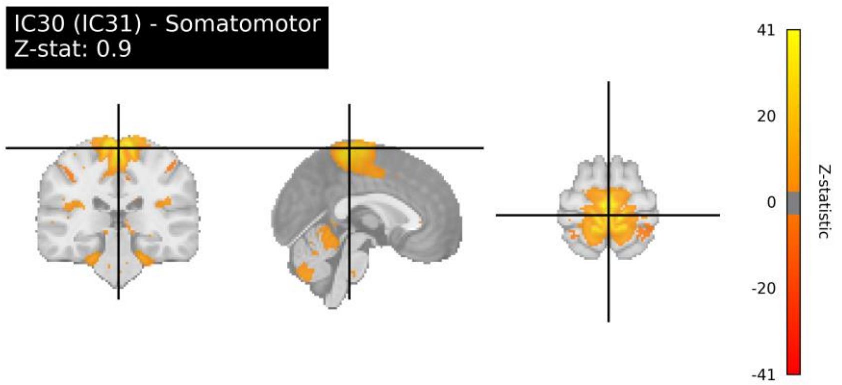
IC30 (IC31) - Somatomotor
Z-stat: 0.9
41
20
0
-20
-41
Z-statistic

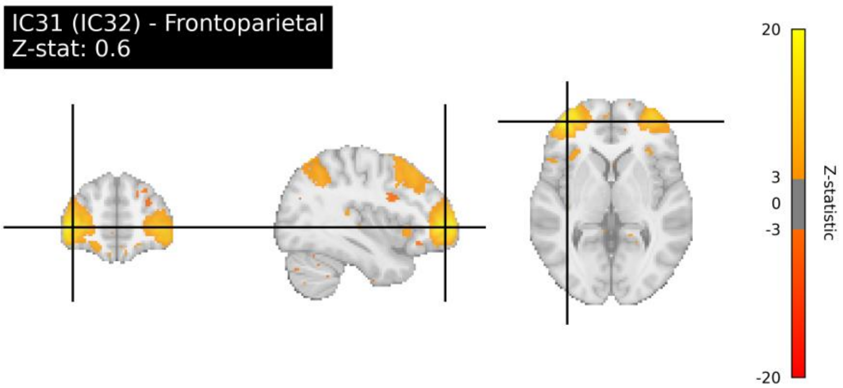
IC31 (IC32) - Frontoparietal
Z-stat: 0.6
20
3
0
-3
-20
Z-statistic

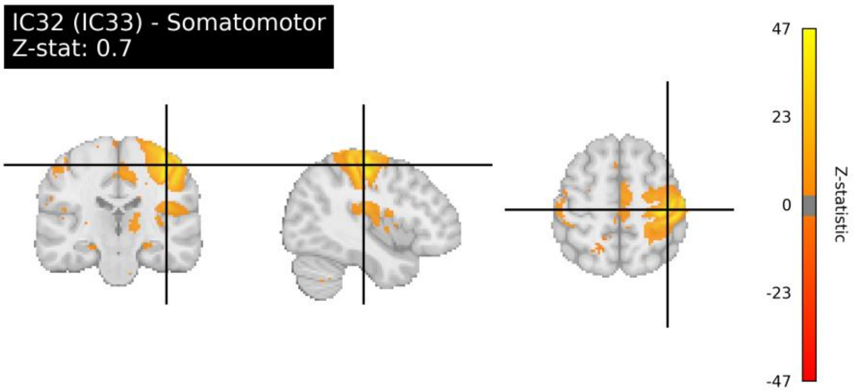
IC32 (IC33) - Somatomotor
Z-stat: 0.7
47
23
0
-23
-47
Z-statistic

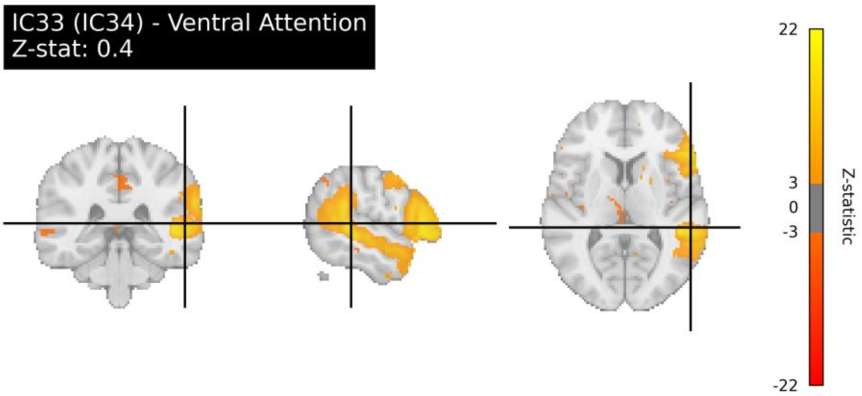
IC33 (IC34) - Ventral Attention
Z-stat: 0.4
22
3
0
-3
-22
Z-statistic

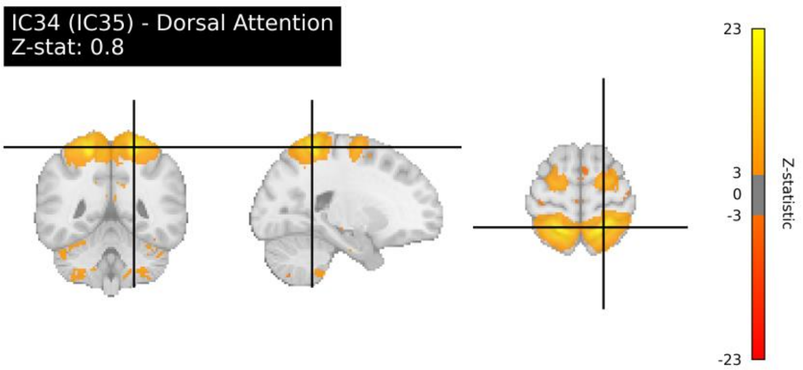
IC34 (IC35) - Dorsal Attention
Z-stat: 0.8
23
3
0
-3
-23
Z-statistic

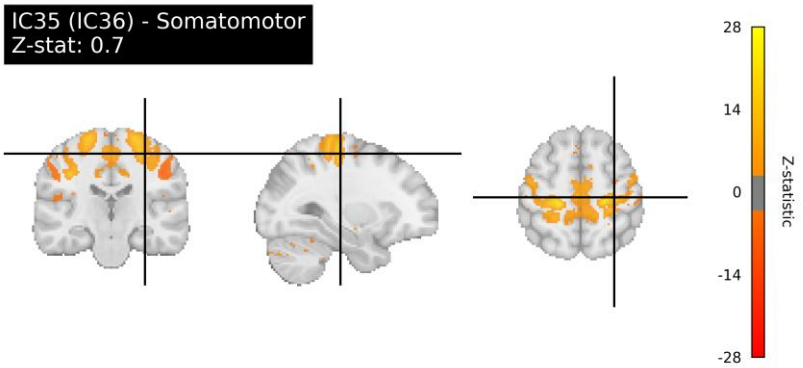
IC35 (IC36) - Somatomotor
Z-stat: 0.7
28
14
0
-14
-28
Z-statistic

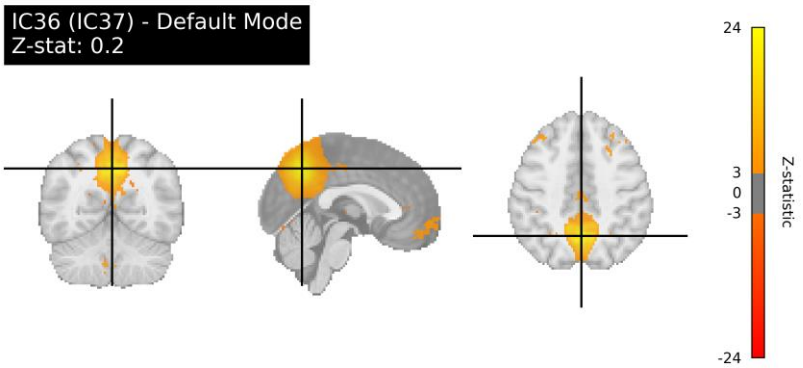
IC36 (IC37) - Default Mode
Z-stat: 0.2
24
3
0
-3
-24
Z-statistic

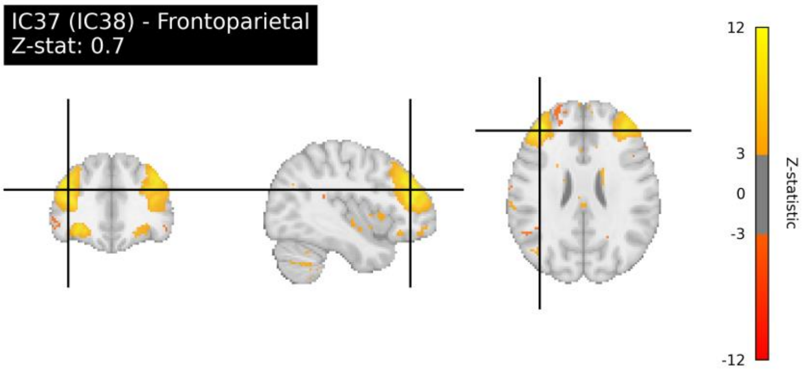
IC37 (IC38) - Frontoparietal
Z-stat: 0.7
12
3
0
-3
-12
Z-statistic

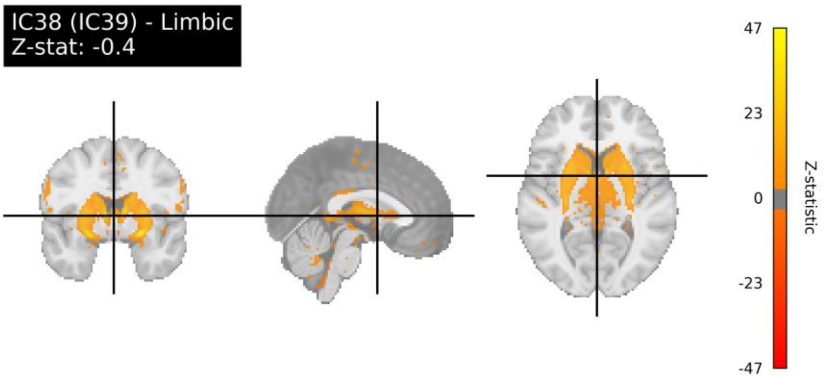
IC38 (IC39) - Limbic
Z-stat: -0.4
47
23
0
-23
-47
Z-statistic

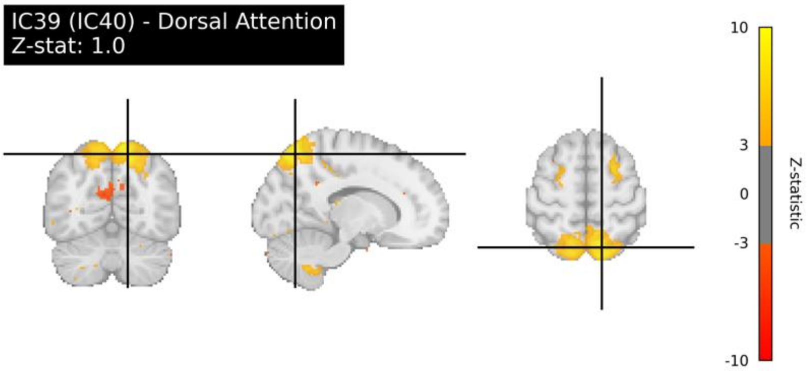
IC39 (IC40) - Dorsal Attention
Z-stat: 1.0
10
3
0
-3
-10
Z-statistic

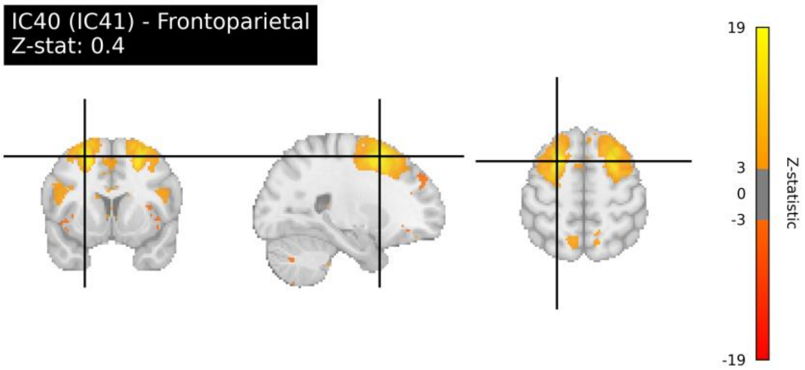
IC40 (IC41) - Frontoparietal
Z-stat: 0.4
19
3
0
-3
-19
Z-statistic

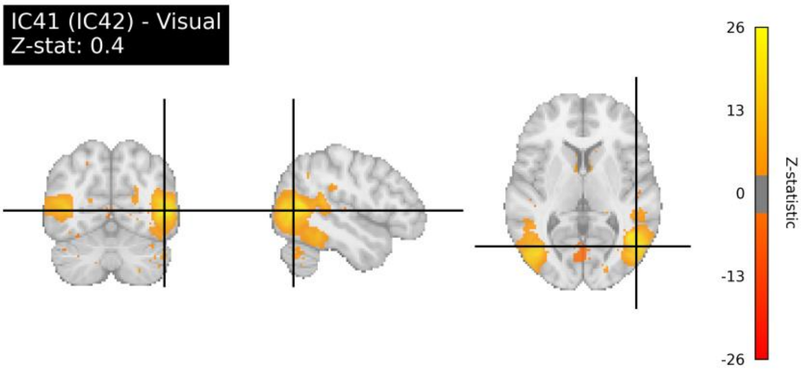
IC41 (IC42) - Visual
Z-stat: 0.4
26
13
0
-13
-26
Z-statistic

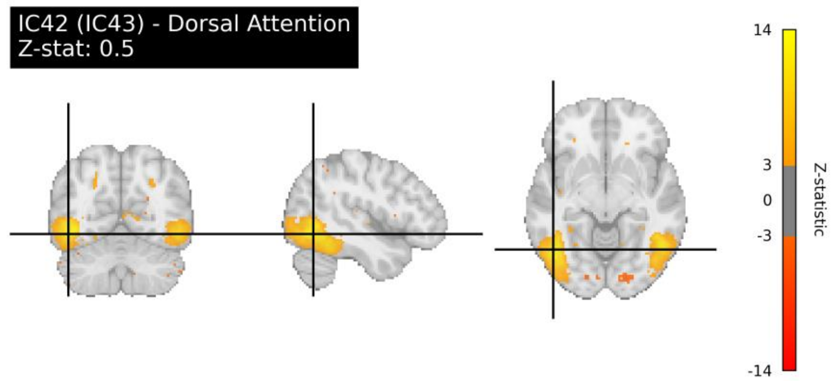
IC42 (IC43) - Dorsal Attention
Z-stat: 0.5
14
3
0
-3
-14
Z-statistic

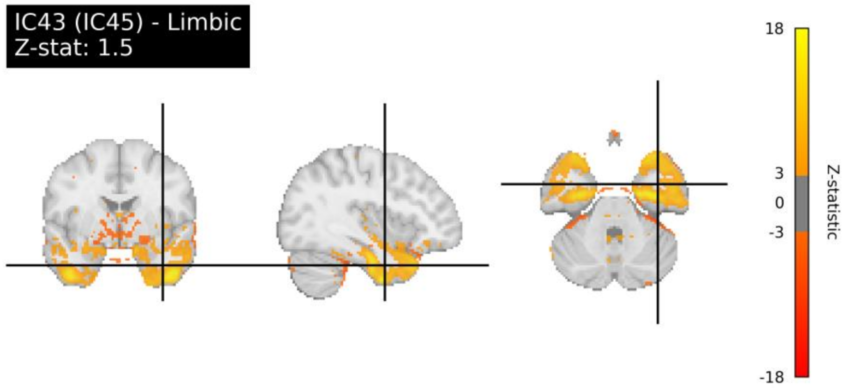
IC43 (IC45) - Limbic
Z-stat: 1.5
18
3
0
-3
-18
Z-statistic

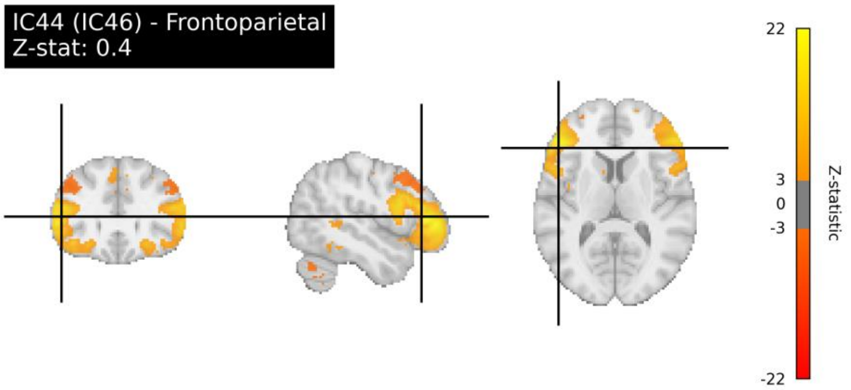
IC44 (IC46) - Frontoparietal
Z-stat: 0.4
22
3
0
-3
-22
Z-statistic

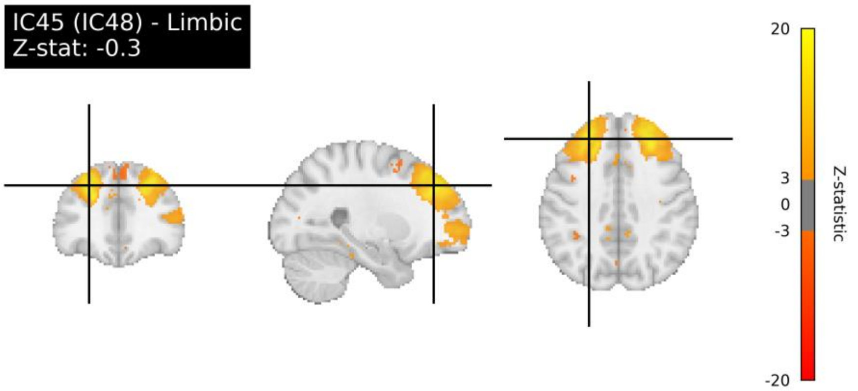
IC45 (IC48) - Limbic
Z-stat: -0.3
20
3
0
-3
-20
Z-statistic

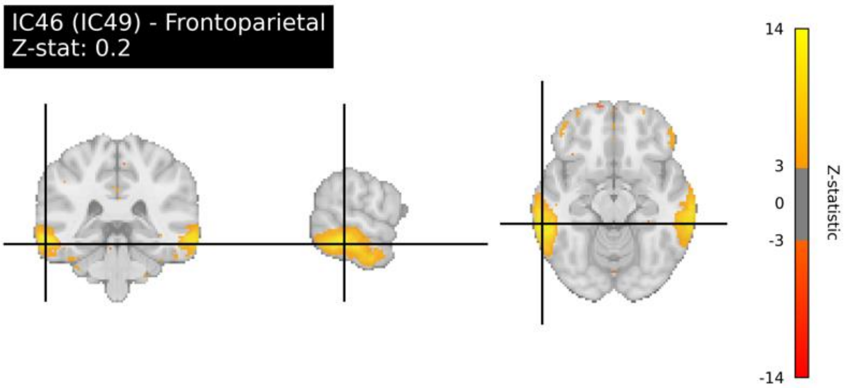
IC46 (IC49) - Frontoparietal
Z-stat: 0.2
14
3
0
-3
-14
Z-statistic

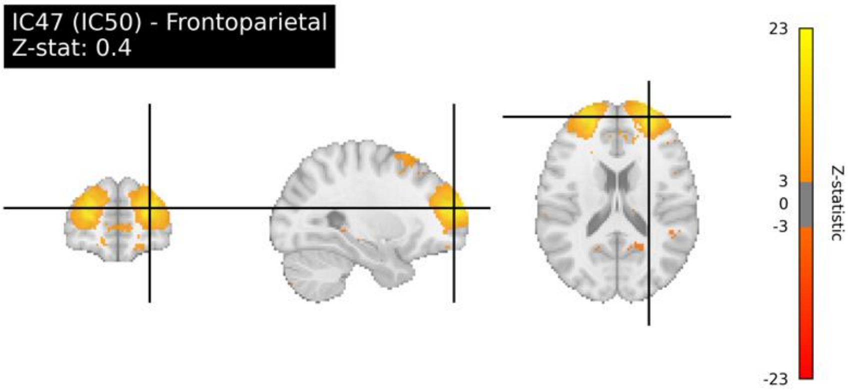
IC47 (IC50) - Frontoparietal
Z-stat: 0.4
23
3
0
-3
-23
Z-statistic

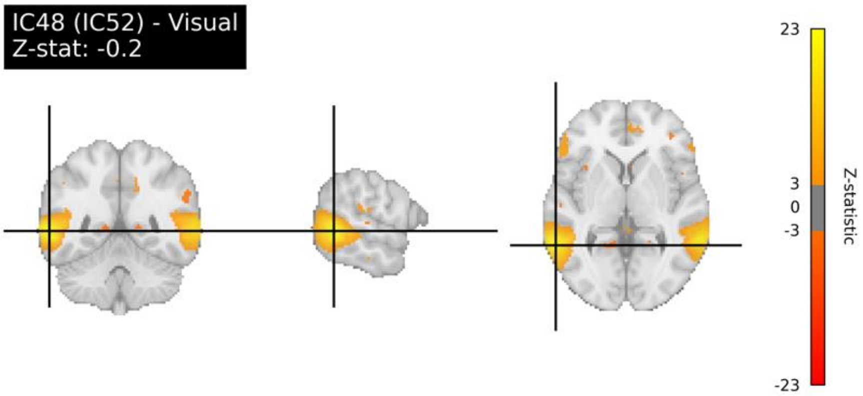
IC48 (IC52) - Visual
Z-stat: -0.2
23
3
0
-3
-23
Z-statistic

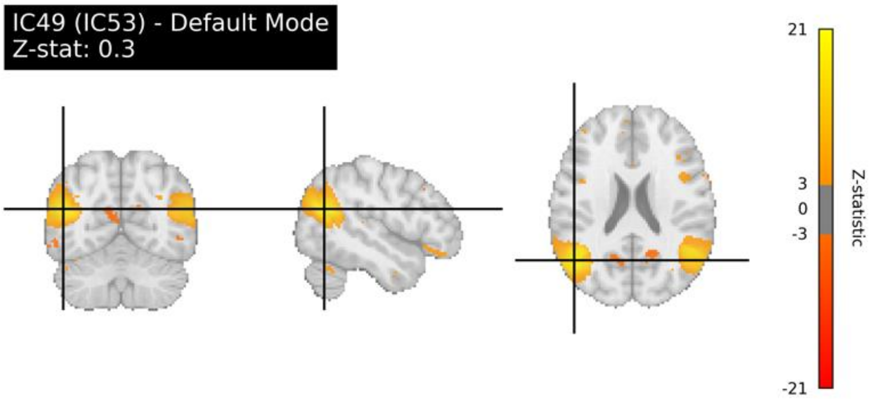
IC49 (IC53) - Default Mode
Z-stat: 0.3
21
3
0
-3
-21
Z-statistic

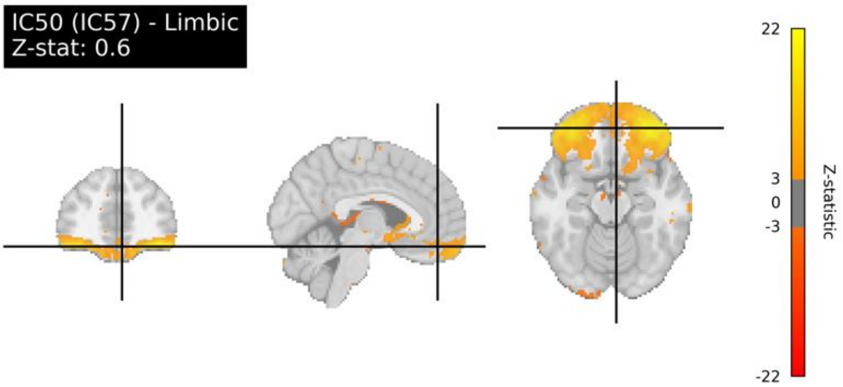

IC50 (IC57) - Limbic
Z-stat: 0.6
22
3
0
-3
-22
Z-statistic

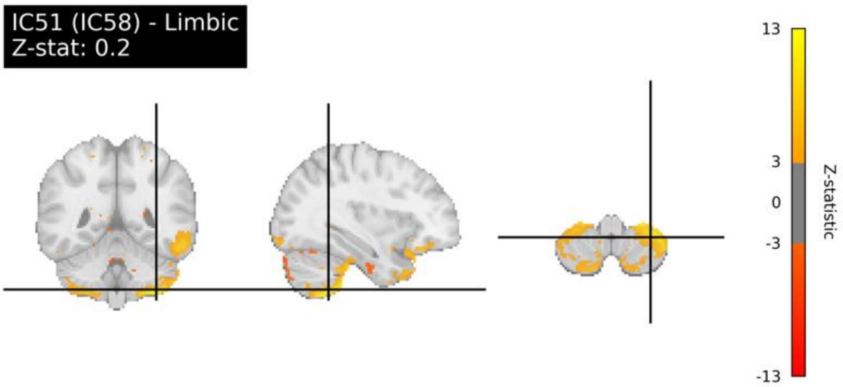

IC51 (IC58) - Limbic
Z-stat: 0.2
13
3
0
-3
-13
Z-statistic

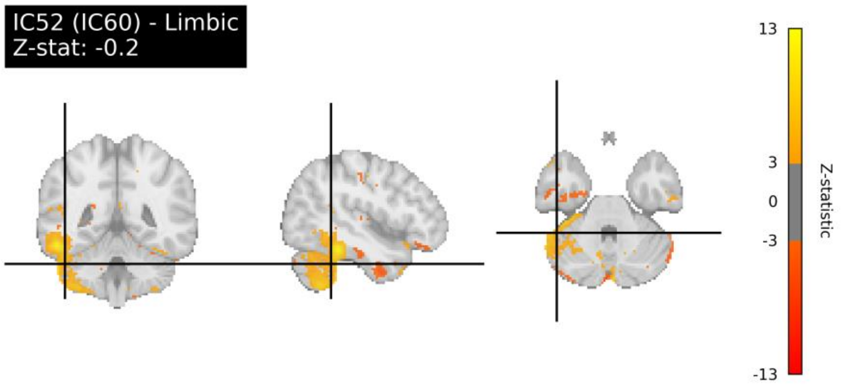

IC52 (IC60) - Limbic
Z-stat: -0.2
13
3
0
-3
-13
Z-statistic

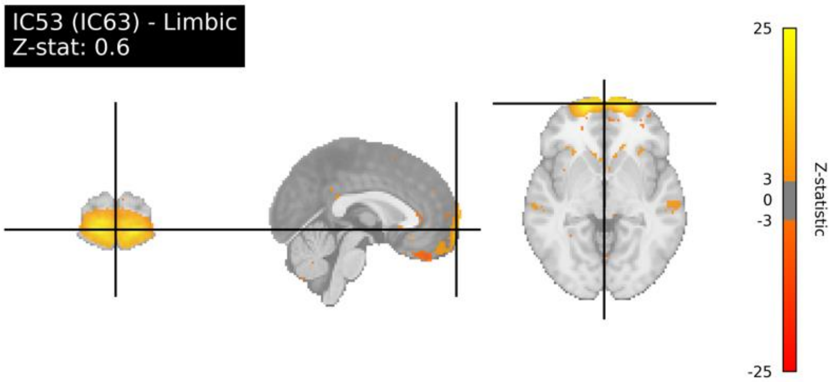

IC53 (IC63) - Limbic
Z-stat: 0.6
25
3
0
-3
-25
Z-statistic

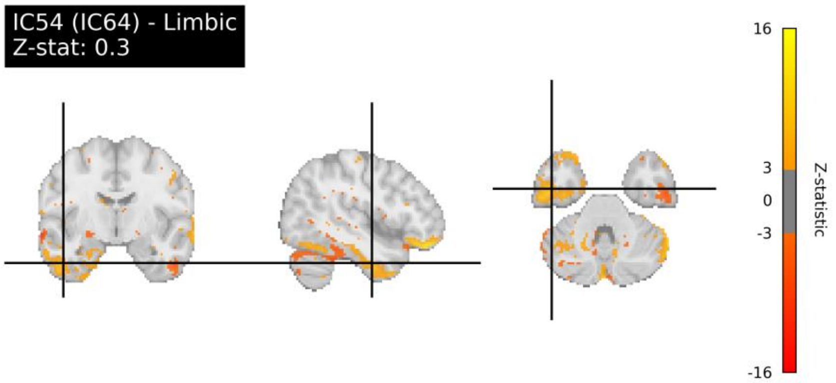

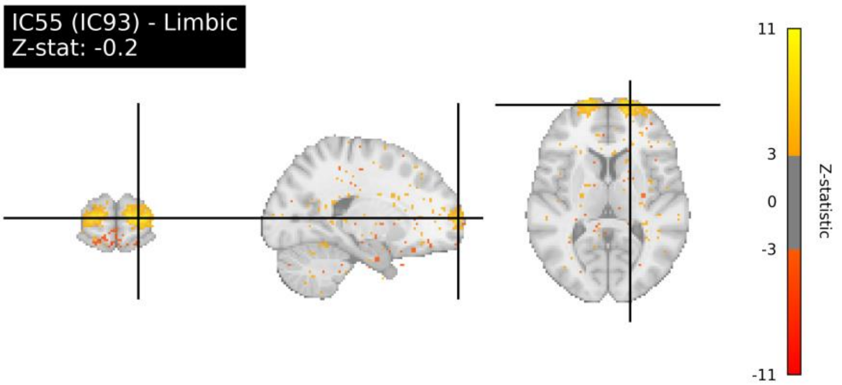

**Fig. S5.** Feature importance plot for sMRI neuroimaging phenotype with the highest predictive performance for cognition. Bars show the direction and magnitude of Pearson correlations ($r \geq 0.2$) between the predicted *g*-factor and volumes of subcortical structures derived from FreeSurfer subcortical volumetric subsegmentation. Pearson *r* was computed in outer-fold test sets pooled across five folds.

763

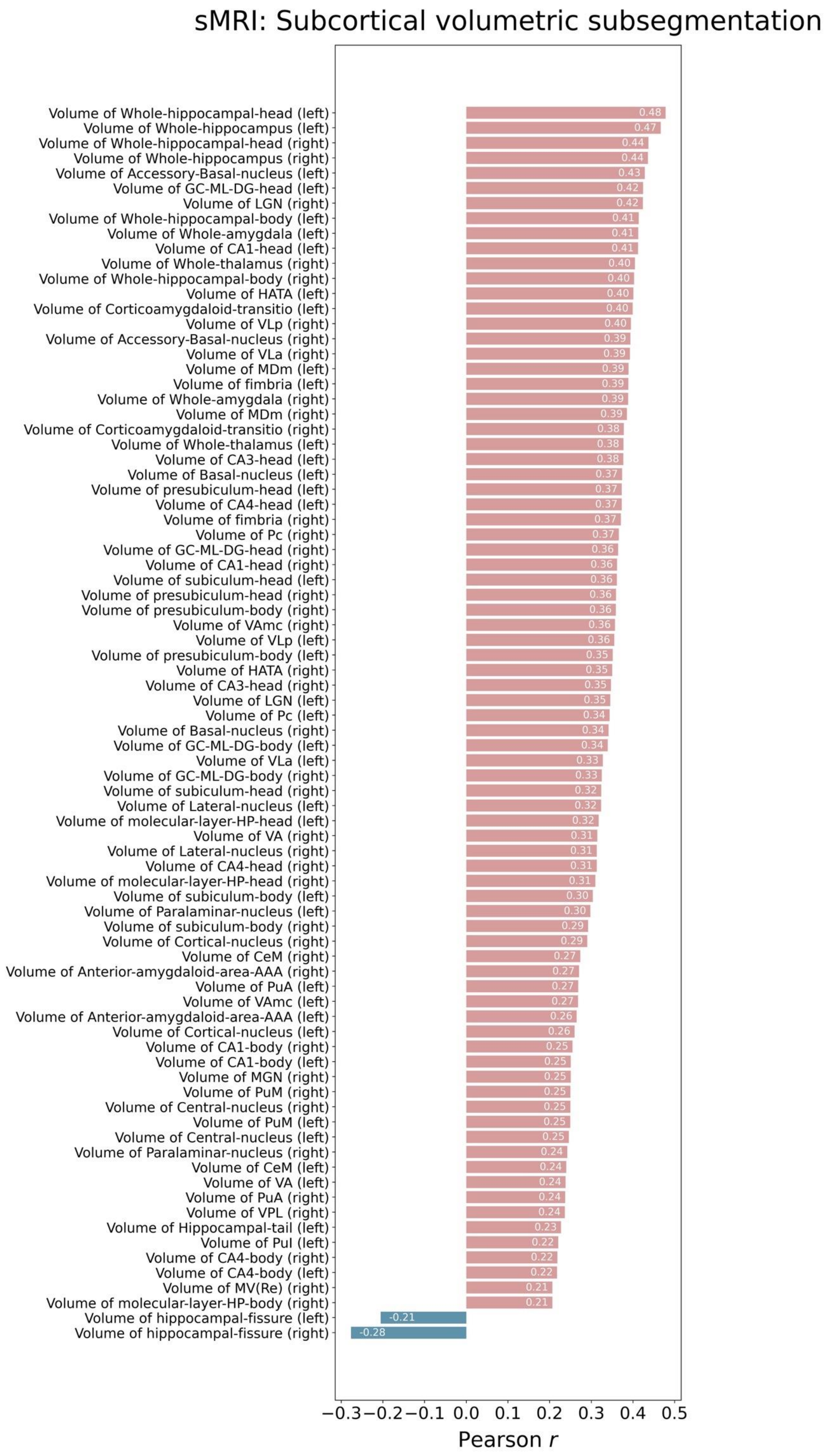

sMRI: Subcortical volumetric subsegmentation
Volume of Whole-hippocampal-head (left) 0.48
Volume of Whole-hippocampus (left) 0.47
Volume of Whole-hippocampal-head (right) 0.44
Volume of Whole-hippocampus (right) 0.44
Volume of Accessory-Basal-nucleus (left) 0.43
Volume of GC-ML-DG-head (left) 0.42
Volume of LGN (right) 0.42
Volume of Whole-hippocampal-body (left) 0.41
Volume of Whole-amygdala (left) 0.41
Volume of CA1-head (left) 0.41
Volume of Whole-thalamus (right) 0.40
Volume of Whole-hippocampal-body (right) 0.40
Volume of HATA (left) 0.40
Volume of Corticoamygdaloid-transitio (left) 0.40
Volume of VLp (right) 0.40
Volume of Accessory-Basal-nucleus (right) 0.39
Volume of VLa (right) 0.39
Volume of MDm (left) 0.39
Volume of fimbria (left) 0.39
Volume of Whole-amygdala (right) 0.39
Volume of MDm (right) 0.39
Volume of Corticoamygdaloid-transitio (right) 0.38
Volume of Whole-thalamus (left) 0.38
Volume of CA3-head (left) 0.38
Volume of Basal-nucleus (left) 0.37
Volume of presubiculum-head (left) 0.37
Volume of CA4-head (left) 0.37
Volume of fimbria (right) 0.37
Volume of Pc (right) 0.37
Volume of GC-ML-DG-head (right) 0.36
Volume of CA1-head (right) 0.36
Volume of subiculum-head (left) 0.36
Volume of presubiculum-head (right) 0.36
Volume of presubiculum-body (right) 0.36
Volume of VAmc (right) 0.36
Volume of VLp (left) 0.36
Volume of presubiculum-body (left) 0.35
Volume of HATA (right) 0.35
Volume of CA3-head (right) 0.35
Volume of LGN (left) 0.35
Volume of Pc (left) 0.34
Volume of Basal-nucleus (right) 0.34
Volume of GC-ML-DG-body (left) 0.34
Volume of VLa (left) 0.33
Volume of GC-ML-DG-body (right) 0.33
Volume of subiculum-head (right) 0.32
Volume of Lateral-nucleus (left) 0.32
Volume of molecular-layer-HP-head (left) 0.32
Volume of VA (right) 0.31
Volume of Lateral-nucleus (right) 0.31
Volume of CA4-head (right) 0.31
Volume of molecular-layer-HP-head (right) 0.31
Volume of subiculum-body (left) 0.30
Volume of Paralaminar-nucleus (left) 0.30
Volume of subiculum-body (right) 0.29
Volume of Cortical-nucleus (right) 0.29
Volume of CeM (right) 0.27
Volume of Anterior-amygdaloid-area-AAA (right) 0.27
Volume of PuA (left) 0.27
Volume of VAmc (left) 0.27
Volume of Anterior-amygdaloid-area-AAA (left) 0.26
Volume of Cortical-nucleus (left) 0.26
Volume of CA1-body (right) 0.25
Volume of CA1-body (left) 0.25
Volume of MGN (right) 0.25
Volume of PuM (right) 0.25
Volume of Central-nucleus (right) 0.25
Volume of PuM (left) 0.25
Volume of Central-nucleus (left) 0.25
Volume of Paralaminar-nucleus (right) 0.24
Volume of CeM (left) 0.24
Volume of VA (left) 0.24
Volume of PuA (right) 0.24
Volume of VPL (right) 0.24
Volume of Hippocampal-tail (left) 0.23
Volume of Pul (left) 0.22
Volume of CA4-body (right) 0.22
Volume of CA4-body (left) 0.22
Volume of MV(Re) (right) 0.21
Volume of molecular-layer-HP-body (right) 0.21
Volume of hippocampal-fissure (left) -0.21
Volume of hippocampal-fissure (right) -0.28
−0.3 −0.2 −0.1 0.0 0.1 0.2 0.3 0.4 0.5
Pearson r

#### Stacking

**Table S16.** Mean (averaged across five folds) out-of-sample predictive performance of MRI modalities stacked using four machine learning algorithms

| | Algorithm | $R^2$ | Pearson *r* | MSE | MAE |
|---|---|---|---|---|---|
| **dwMRI** | ElasticNet | 0.027 | 0.227 | 0.97 | 0.782 |
| | **Random Forest** | **0.073** | **0.265** | **0.924** | **0.764** |
| | Support Vector Regression | 0.036 | 0.247 | 0.961 | 0.777 |
| | XGBoost | 0.061 | 0.26 | 0.936 | 0.768 |
| **rsMRI** | ElasticNet | 0.100 | 0.325 | 0.897 | 0.752 |
| | **Random Forest** | **0.105** | **0.325** | **0.891** | **0.75** |
| | Support Vector Regression | 0.101 | 0.327 | 0.896 | 0.751 |
| | XGBoost | 0.102 | 0.326 | 0.895 | 0.751 |
| **sMRI** | ElasticNet | 0.094 | 0.294 | 0.903 | 0.755 |
| | Random Forest | 0.093 | 0.293 | 0.904 | 0.755 |
| | **Support Vector Regression** | **0.095** | **0.298** | **0.902** | **0.753** |
| | XGBoost | 0.095 | 0.296 | 0.902 | 0.754 |
| **All MRI modalities** | ElasticNet | 0.131 | 0.374 | 0.866 | 0.738 |
| | Random Forest | 0.152 | 0.383 | 0.845 | 0.729 |
| | Support Vector Regression | 0.139 | 0.383 | 0.859 | 0.734 |
| | **XGBoost** | **0.159** | **0.398** | **0.838** | **0.726** |

$R^2$ coefficient of determination, *MSE* mean squared error, *MAE* mean absolute error, *dwMRI* diffusion-weighted MRI, *rsMRI* resting-state MR, *sMRI* T1-weighted and T2-weighted structural MRI. The algorithms that yielded the highest $R^2$ are highlighted in bold.

### S11. Commonality Analysis

**Table S17.** Variance in the *g*-factor captured by mental health and dwMRI neuroimaging phenotypes in the commonality analysis

| | Unique: MH | Unique: dwMRI | Common: MH, dwMRI | Total Variance | % Var. Exp. dwMRI |
|---|---|---|---|---|---|
| **aparc a2009s MSA I Connectome Streamline Count** | 7.29 | 3.33 | 1.74 | 12.36 | 19.27 |
| **Schaefer7n200p MSA I Connectome SIFT2** | 7.43 | 3.30 | 1.60 | 12.33 | 17.72 |
| **Schaefer7n200p MSA I Connectome Streamline Count** | 7.48 | 3.29 | 1.55 | 12.32 | 17.17 |
| **aparc MSA I Connectome Streamline Count** | 7.38 | 3.28 | 1.65 | 12.31 | 18.27 |
| **aparc MSA I Connectome Mean Length** | 7.46 | 3.17 | 1.57 | 12.2 | 17.39 |
| **aparc MSA I Connectome SIFT2** | 7.36 | 3.14 | 1.67 | 12.17 | 18.49 |
| **Schaefer7n200p MSA I Connectome FA** | 7.5 | 3.08 | 1.53 | 12.11 | 16.94 |
| **aparc a2009s MSA I Connectome SIFT2** | 7.46 | 3.02 | 1.57 | 12.05 | 17.39 |
| **aparc a2009s MSA I Connectome FA** | 7.43 | 2.97 | 1.60 | 12 | 17.72 |
| **aparc MSA I Connectome FA** | 7.48 | 2.96 | 1.55 | 11.99 | 17.17 |
| **Schaefer7n500p MSA IV Connectome SIFT2** | 7.57 | 2.93 | 1.46 | 11.96 | 16.17 |
| **Glasser MSA IV Connectome SIFT2** | 7.57 | 2.90 | 1.46 | 11.93 | 16.17 |
| **Schaefer7n500p MSA IV Connectome Streamline Count** | 7.66 | 2.90 | 1.37 | 11.93 | 15.17 |
| **L2 TBSS** | 7.63 | 2.89 | 1.40 | 11.92 | 15.5 |

| | | | | | |
|---|---|---|---|---|---|
| **Glasser MSA IV Connectome Streamline Count** | 7.63 | 2.88 | 1.40 | 11.91 | 15.5 |
| **aparc a2009s MSA I Connectome Mean Length** | 7.54 | 2.87 | 1.49 | 11.9 | 16.5 |
| **Glasser MSA IV Connectome FA** | 7.74 | 2.87 | 1.29 | 11.9 | 14.29 |
| **Schaefer7n500p MSA IV Connectome FA** | 7.81 | 2.75 | 1.22 | 11.78 | 13.51 |
| **Glasser MSA I Connectome FA** | 7.81 | 2.74 | 1.22 | 11.77 | 13.51 |
| **FA TBSS** | 7.73 | 2.71 | 1.30 | 11.74 | 14.4 |
| **L3 TBSS** | 7.82 | 2.70 | 1.21 | 11.73 | 13.4 |
| **L1 TBSS** | 7.8 | 2.69 | 1.23 | 11.72 | 13.62 |
| **Schaefer7n200p MSA I Connectome Mean Length** | 7.87 | 2.65 | 1.16 | 11.68 | 12.85 |
| **MD TBSS** | 7.66 | 2.65 | 1.37 | 11.68 | 15.17 |
| **Glasser MSA I Connectome SIFT2** | 7.72 | 2.60 | 1.31 | 11.63 | 14.51 |
| **Glasser MSA I Connectome Streamline Count** | 7.76 | 2.58 | 1.27 | 11.61 | 14.06 |
| **ICVF TBSS** | 7.77 | 2.53 | 1.26 | 11.56 | 13.95 |
| **MD Probabilistic** | 7.85 | 2.51 | 1.18 | 11.54 | 13.07 |
| **Glasser MSA IV Connectome Mean Length** | 7.89 | 2.42 | 1.14 | 11.45 | 12.62 |
| **Glasser MSA I Connectome Mean Length** | 7.93 | 2.36 | 1.10 | 11.39 | 12.18 |
| **L3 Probabilistic** | 7.92 | 2.34 | 1.11 | 11.37 | 12.29 |
| **L2 Probabilistic** | 7.94 | 2.23 | 1.09 | 11.26 | 12.07 |
| **ISOVF TBSS** | 8.11 | 2.13 | 0.92 | 11.16 | 10.19 |
| **L1 Probabilistic** | 7.96 | 2.07 | 1.07 | 11.1 | 11.85 |
| **ICVF Probabilistic** | 8.12 | 1.87 | 0.91 | 10.9 | 10.08 |
| **OD TBSS** | 8.12 | 1.70 | 0.91 | 10.73 | 10.08 |
| **MO TBSS** | 8.17 | 1.69 | 0.86 | 10.72 | 9.52 |
| **Schaefer7n500p** | 8.33 | 1.52 | 0.70 | 10.55 | 7.75 |

| | | | | | |
|---|---|---|---|---|---|
| **MSA IV Connectome Mean Length** | | | | | |
| **ISOVF Probabilistic** | 8.28 | 1.52 | 0.75 | 10.55 | 8.31 |
| **FA Probabilistic** | 8.39 | 1.45 | 0.64 | 10.48 | 7.09 |
| **OD Probabilistic** | 8.7 | 0.53 | 0.33 | 9.56 | 3.65 |
| **MO Probabilistic** | 8.84 | 0.38 | 0.19 | 9.41 | 2.1 |

*Unique* unique variance, i.e., the proportion of variance in the *g*-factor explained by unique contribution of the predictor (%), *Common* common variance, i.e., the proportion of variance in the *g*-factor explained by common effects of predictor variables (%), *Total Variance* proportion of variance in the *g*-factor explained by all predictor variables (%), *dwMRI* diffusion-weighted MRI, *% Var. Exp. dwMRI* proportion of the relationship between mental health and cognition captured by dwMRI (common effect of dwMRI and mental health ÷ total effect of mental health × 100), *MH* mental health, *FA* fractional anisotropy, *MD* mean diffusivity, *MO* diffusion tensor mode, *L1, L2, L3* eigenvalues of the diffusion tensor, *OD* orientation dispersion index, *ICVF* intracellular volume fraction, *ISOVF* isotropic volume fraction, *TBSS* Tract-Based Spatial Statistics, *aparc* Desikan-Killiany cortical atlas, *aparc a2009s* Destrieux cortical atlas, *Schaefer7n200p* Schaefer Atlas for 7 networks and 200 parcels, *Schaefer7n500p* Schaefer Atlas for 7 networks and 500 parcels, *MSA* Melbourne Subcortical Atlas, *SIFT2* Spherical-Deconvolution Informed Filtering of Tractograms 2.

**Table S18.** Variance in the *g*-factor captured by mental health and rsMRI neuroimaging phenotypes in the commonality analysis

| | **Unique: MH** | **Unique: rsMRI** | **Common: MH, rsMRI** | **Total Variance** | **% Var. Exp. rsMRI** |
|---|---|---|---|---|---|
| **55 IC Functional connectivity (Tangent)** | 6.64 | 5.83 | 2.31 | 14.77 | 25.81 |
| **Schaefer7n200p MSA I Functional connectivity (Full correlation)** | 6.97 | 4.79 | 1.98 | 13.73 | 22.12 |
| **Schaefer7n500p MSA IV** | 7.35 | 4.36 | 1.60 | 13.3 | 17.88 |

| | | | | | |
|---|---|---|---|---|---|
| **Functional connectivity (Full correlation)** | | | | | |
| **Glasser MSA I Functional connectivity (Full correlation)** | 7.41 | 4.14 | 1.54 | 13.08 | 17.21 |
| **Glasser MSA IV Functional connectivity (Full correlation)** | 7.43 | 3.89 | 1.52 | 12.83 | 16.98 |
| **aparc a2009s MSA I Functional connectivity (Full correlation)** | 7.53 | 3.52 | 1.42 | 12.46 | 15.87 |
| **21 IC Functional connectivity** | 7.45 | 3.51 | 1.50 | 12.45 | 16.76 |
| **aparc MSA I Functional connectivity (Full correlation)** | 7.43 | 3.34 | 1.52 | 12.28 | 16.98 |
| **55 IC Amplitudes** | 8.32 | 1.35 | 0.63 | 10.29 | 6.94 |
| **21 IC Amplitudes** | 8.58 | 0.77 | 0.37 | 9.71 | 4.03 |

*Unique* unique variance, i.e., the proportion of variance in the *g*-factor explained by unique contribution of the predictor (%), *Common* common variance, i.e., the proportion of variance in the *g*-factor explained by common effects of predictor variables (%), *Total Variance* proportion of variance in the *g*-factor explained by all predictor variables (%), *rsMRI* resting-state MRI, *% Var. Exp. rsMRI* proportion of the relationship between mental health and cognition captured by rsMRI (common effect of rsMRI and mental health ÷ total effect of mental health × 100), *MH* mental health, *IC* independent components, *aparc* Desikan-Killiany cortical atlas, *aparc a2009s* Destrieux cortical atlas, *Schaefer7n200p* Schaefer Atlas for 7 networks and 200 parcels, *Schaefer7n500p* Schaefer Atlas for 7 networks and 500 parcels, *MSA* Melbourne Subcortical Atlas.

**Table S19.** Variance in the *g*-factor captured by mental health and sMRI neuroimaging phenotypes in the commonality analysis

| | Unique: MH | Unique: sMRI | Common: MH, sMRI | Total Variance | % Var. Exp. sMRI |
|---|---|---|---|---|---|
| **Subcortical Volumetric Subsegmentation** | 6.89 | 4.18 | 1.92 | 12.98 | 21.79 |
| **ASEG Volume** | 6.98 | 3.98 | 1.83 | 12.78 | 20.68 |
| **FSL FAST** | 7.29 | 3.41 | 1.52 | 12.21 | 17.25 |
| **Whole-brain T1/T2** | 7.47 | 3.16 | 1.34 | 11.96 | 15.21 |
| **Desikan GM/WM Intensity** | 7.56 | 2.61 | 1.25 | 11.41 | 14.19 |
| **ASEG Mean Thickness** | 7.44 | 2.60 | 1.37 | 11.4 | 15.45 |
| **aparc.a2009s Volume** | 7.81 | 2.20 | 1.00 | 11 | 11.35 |
| **aparc.a2009s Area** | 7.98 | 1.62 | 0.83 | 10.42 | 9.32 |
| **aparc.a2009s Mean Thickness** | 7.96 | 1.62 | 0.85 | 10.42 | 9.55 |
| **FSL FIRST** | 7.95 | 1.61 | 0.86 | 10.41 | 9.76 |
| **DKT Volume** | 8.03 | 1.56 | 0.78 | 10.36 | 8.85 |
| **Desikan WM Volume** | 8.06 | 1.50 | 0.75 | 10.3 | 8.41 |
| **BA *ex-vivo* Volume** | 8.1 | 1.47 | 0.71 | 10.27 | 7.95 |
| **Desikan WM Mean Thickness** | 8.05 | 1.44 | 0.76 | 10.24 | 8.63 |
| **DKT Mean Thickness** | 8.08 | 1.44 | 0.73 | 10.24 | 8.18 |
| **Desikan Pial** | 8.14 | 1.38 | 0.67 | 10.18 | 7.5 |
| **DKT Area** | 8.18 | 1.12 | 0.63 | 9.92 | 7.05 |
| **Desikan WM Area** | 8.13 | 1.08 | 0.68 | 9.88 | 7.72 |
| **BA *ex-vivo* Mean Thickness** | 8.3 | 0.99 | 0.51 | 9.79 | 5.68 |
| **BA *ex-vivo* Area** | 8.39 | 0.73 | 0.42 | 9.53 | 4.77 |

*Unique* unique variance, i.e., proportion of variance in the *g*-factor explained by unique contribution of the predictor (%), *Common* common variance, i.e., proportion of variance in the *g*-factor explained by common effects of predictor variables (%), *Total Variance* proportion of variance in the *g*-factor explained by all predictor variables (%), *sMRI* T1-weighted (T1w) and T2-weighted (T2w) structural MRI, *% Var. Exp. sMRI* proportion of the relationship between mental health and cognition captured by sMRI (common effect of sMRI and mental health ÷ total effect of mental health × 100), *MH* mental health, *ASEG* FreeSurfer automated subcortical volumetric segmentation, *FSL FAST* FSL FMRIB's Automated Segmentation Tool, *a2009s* FreeSurfer Destrieux, *FSL FIRST* FMRIB's Integrated Registration and Segmentation Tool, *BA* FreeSurfer *ex-vivo* Brodmann Area Maps, *DKT* Desikan-Killiany-Tourville.

**Table S20.** Variance in the *g*-factor captured by mental health and stacked MRI modalities in the commonality analysis

| | Unique: MH | Unique: MRI | Common: MH, MRI | Total Variance | % Var. Exp. MRI |
|---|---|---|---|---|---|
| **dwMRI** | 6.73 | 4.78 | 2.3 | 13.81 | 25.47 |
| **rsMRI** | 6.28 | 6.95 | 2.67 | 15.9 | 29.83 |
| **sMRI** | 6.05 | 6.28 | 2.8 | 15.13 | 31.64 |
| **All MRI modalities** | 4.55 | 10.88 | 4.21 | 19.64 | 48.06 |

*Unique* unique variance, i.e., the proportion of variance in the *g*-factor explained by the unique contribution of the predictor (%), *Common* common variance, i.e., the proportion of variance in the *g*-factor explained by common effects of predictor variables (%), *Total Variance* proportion of variance in the *g*-factor explained by all predictor variables (%), *% Var. Exp. MRI* proportion of the relationship between mental health and cognition captured by MRI data (a common effect of MRI data and mental health ÷ total effect of mental health × 100), *MH* mental health, *dwMRI* diffusion-weighted MRI, *rsMRI* resting-state MRI, *sMRI* T1-weighted and T2-weighted structural MRI.

**Table S21.** Variance in the *g*-factor captured by mental health and three MRI modalities in the commonality analysis

| | Unique: MH | Unique: MRI | Unique: Age&Sex | Common: MH, MRI | Common: MH, Age&Sex | Common: MRI, Age&Sex | Common: MH, MRI, Age&Sex | Total Variance |
|---|---|---|---|---|---|---|---|---|
| **dwMRI** | 4.78 | 0.79 | 4.78 | 0.29 | 1.95 | 2.01 | 3.99 | 18.59 |
| **rsMRI** | 3.7 | 5.01 | 6.77 | 1.34 | 2.58 | 1.33 | 1.94 | 22.67 |
| **sMRI** | 4.74 | 0.86 | 3.16 | 0.24 | 1.31 | 2.51 | 5.42 | 18.24 |

| | | | | | | | | |
|---|---|---|---|---|---|---|---|---|
| **All MRI modalities** | 3.52 | 5.29 | 2.76 | 1.45 | 1.03 | 2.76 | 5.59 | 22.4 |

*Unique* unique variance, i.e., proportion of variance in the *g*-factor explained by unique contribution of the predictor (%), *Common* common variance, i.e., proportion of variance in the *g*-factor explained by common effects of predictor variables (%), *Total Variance* proportion of variance in the *g*-factor explained by all predictor variables (%), *MH* mental health, *dwMRI* diffusion-weighted MRI, *rsMRI* resting-state MRI, *sMRI* T1-weighted and T2-weighted structural MRI. *Age&Sex* is defined as a combination of age, sex, $age^2$, age×sex, and $age^2$×sex.
